# Dynamics of Gut Metabolome and Microbiome Maturation during Early Life

**DOI:** 10.1101/2023.05.29.23290441

**Authors:** Anna-Katariina Aatsinki, Santosh Lamichhane, Heidi Isokääntä, Partho Sen, Matilda Kråkström, Marina Amaral Alves, Anniina Keskitalo, Eveliina Munukka, Hasse Karlsson, Laura Perasto, Minna Lukkarinen, Matej Oresic, Henna-Maria Kailanto, Linnea Karlsson, Leo Lahti, Alex M Dickens

**Affiliations:** Centre for Population Health Research, University of Turku and Turku University Hospital, Turku, Finland; FinnBrain Birth Cohort Study, Turku Brain and Mind Center, Department of Clinical Medicine, University of Turku, Turku, Finland; Turku Bioscience Centre, University of Turku and Åbo Akademi University, 20520 Turku, Finland; Walter Mors Institute of Research on Natural Products, Federal University of Rio de Janeiro, Rio de Janeiro, RJ 21941-902, Brazil; Research Center for Infections and Immunity, Institute of Biomedicine, University of Turku, Turku, Finland; Department of Clinical Microbiology, Turku University Hospital, 20520 Turku, Finland; Faculty of Medicine, Microbiome Biobank, University of Turku and Turku University Hospital, Turku, Finland; Department of Psychiatry, University of Turku and Turku University Hospital, Turku, Finland; Department of Pediatrics and Adolescent Medicine, Turku University Hospital and University of Turku, Turku, Finland; School of Medical Sciences, Örebro University, 702 81 Örebro, Sweden; Department of Computing, University of Turku, 20014 Turku, Finland; Department of Chemistry, University of Turku, 20520 Turku, Finland

## Abstract

Early-life gut microbiome-metabolome crosstalk has a pivotal role in the maintenance of host physiology. However, our understanding on early-life gut microbiome-metabolome maturation trajectories in humans remains limited. This study aims to explore the longitudinal patterns of gut metabolites during early life, and how they are related to gut microbiota composition in birth cohort samples of n = 670 children collected at 2.5 (n=272), 6 (n=232), 14 (n=289), and 30 months (n=157) of age.

Factor analysis showed that breastfeeding has an effect on several metabolites including secondary bile acids. We found that the prevalent gut microbial abundances were associated with metabolite levels, especially in the 2.5 months-olds. We also demonstrated that the prevalent early colonizers *Bacteroides, Escherichia* and *Bifidobacterium* abundances associated with microbial metabolites bile acids especially in the breastfed infants.

Taken together, our results suggests that as the microbiome matures during the early-life there is an association with the metabolome composition in an analogous fashion to how the genome information mature during early life.

## Introduction

The human gut harbors an estimated 500–1000 species of microbes on average in the adult population. The gut microbiome, which includes the by-products of microbes, as amino acids, vitamins, and organic acids, and the host interaction, is considered to be an “essential organ” within human beings^1,2^. The process of gut microbiome colonization after birth has been intensively studied during the last decade^3-5^. Recent studies have demonstrated that gut microbiome plays a crucial role in modulating human health and disease, including many common health complications such as: inflammatory bowel disease^6^, obesity^7^, various neurological and psychiatric disorders^8,9^. Growing evidence suggests that the process of gut microbiota maturation in early life relates to the trajectories of human disease and their resultant outcomes later in life^10,11^. These studies have demonstrated the importance of understanding the early-life gut microbiome composition and its function in order to understand long-term human health outcomes.

Crosstalk between the host and gut microbiome is vital for maintaining human metabolic capacity^12^. Many complex interactions between the host and gut microbiome occur via enterohepatic circulation from the liver to the intestine and back, and the metabolite production both in the host and the microbiome begins already prenatally^13,14^. As such, profiling fecal metabolites can provide an indirect functional readout of the gut microbiome composition. The metabolites can act as an intermediate phenotype mediating host-microbiome interactions^15^. In fact, bidirectional interactions exist between the gut microbiome and metabolome^16^. For example, microbial biotransformation of bile acids (BAs) can regulate human physiology and in turn the overall host BA pool can control the microbial diversity^17^. Intriguingly, a recent rodent study suggests that gut metabolome drives gut microbiota development and maturation^18^. However, our understanding on early-life gut microbiome-metabolome maturation trajectories in humans is limited^4,19-21^.

Here, we study how early-life gut metabolome is associated with the maturation of gut microbiota. More specifically we aimed to identify trajectories of fecal metabolome which may drive the maturation of early gut microbiome.

## Results

The study subjects are described in the Supplementary Table 1. We analyzed longitudinal metabolome, using both mass spectrometry based targeted and untargeted techniques, and microbiome in stool samples collected at 2.5 (n=444 for microbiome, n=272 for metabolome), 6 (n=256 for microbiome, n=232 for metabolome), 14 (n=302 for microbiome and n=289 metabolome), and 30 (n=207 for microbiome and n=157 for metabolome) months (mo) of age. The metabolomics dataset used for the analysis included identified metabolites from the following classes: short chain fatty acids (SCFA), BAs (taurine and glycine conjugated), amino acids, carboxylic acids (mainly free fatty acids and other organic acids), hydroxy acids, phenolic compounds, alcohols, and sugar derivatives. There was no complete overlap between the timepoints: 37 children had microbiome data from all the timepoints, whereas two or three samples were available from 208 or 110 children, respectively. Descriptive findings from the 16S rRNA analysis are shown in the Supplementary figures 1-3.

**Figure 1.**
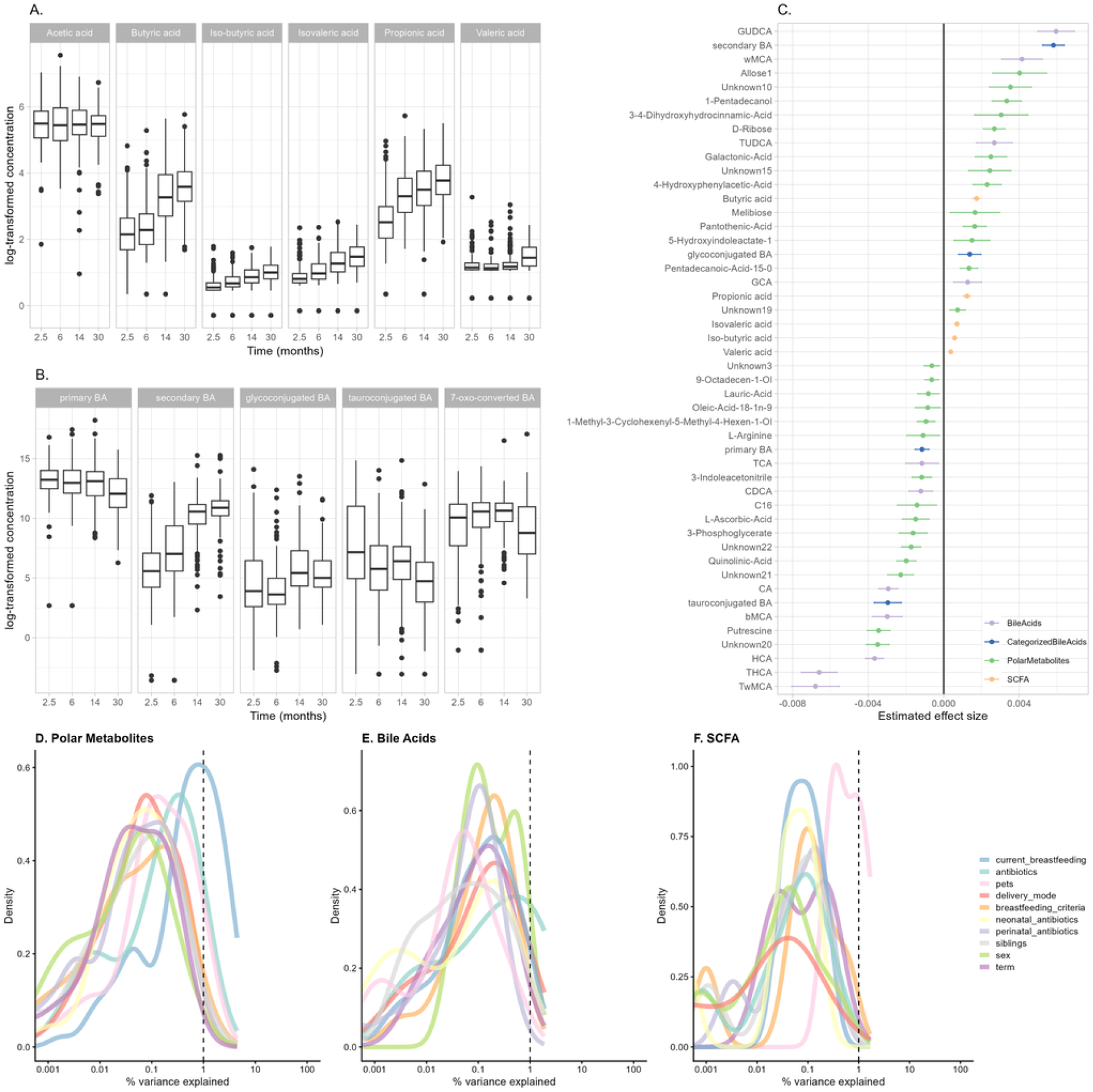
A, B. The average changes in SCFAs and BAs concentrations observed across different age groups. C. Effect-size (age coefficients) for individual metabolites as estimated from the linear mixed models. Lighter colours indicate lower concentration. Density plots showing % of variances explained by total D. polar metabolites, E. BAs and F. SCFAs associated with the clinical and demographic factors.

### Fecal Metabolome age-trends

First, we explored how the gut metabolome changes by age. As expected, age-related variation displayed the major effect on the gut metabolome. Most of the SCFAs, except for acetic acid, increased with age (Fig. 1A.). Individual BAs and polar metabolites showed no clear age-related patterns (Fig 1.). Secondary BAs were positively, whilst, primary and secondary tauroconjugated BAs remained negatively associated with age (Fig. 1B., G.) Glycoconjugated BAs were positively associated with age, however, this association attenuated when adjusting for breastfeeding (Supplementary Figure 8). Some of the metabolites including 5-Hydroxyindoleacetate, 4-Hydroxyphenylacetic acid and multiple unidentified polar metabolites that had a significant age trend were attenuated when adjusting for breastfeeding associated with breastfeeding (Supplementary Figure 8).

### Metabolome associations with factors known to associate with microbiome colonization

In order to understand the overall contributions of various factors to gut metabolome, we performed variance analysis using variables previously shown to associate with gut microbiota maturation, *i*.*e*. breastfeeding, delivery mode, antibiotics intake, prenatal birth, biological sex assigned at birth, pet ownership and having siblings. In general, demographic exposures explained on average <1 % of variance in polar metabolites, SCFAs, and BA concentrations (Fig.1D.,E.,F., Supplementary Table 3).

Next, to study in more detail how gut metabolites related to demographic exposures, we built linear mixed-effect models. We found that breastfed infants had lower concentration of secondary, 7-oxo-converted and individual tauro- and glycoconjugated BAs especially at an early age (Fig. 2A,B, Supplementary Figure 10). Vaginal delivery was related to lower concentration of hydroxyindoleacetate (Fig. 1C), and exposure to intravenous antibiotics in the neonatal period was associated with higher butyric acid concentration (Fig.1D). In the cross-sectional group comparison (Supplementary Figures 9-51), vaginally born infants had lower concentration of 7-oxo-converted BA at 14 mo. The primary BAs at 14 mo, and tauroconjugated BAs at 2.5 mo were also lower (Supplementary Figures 48, 50). Likewise, breastfed infants had lower concentration of 7-oxo-converted and primary BAs at 2.5 months (Supplementary Figure 48). Having pets was positively associated with tauroconjugated BAs concentration at 14 mo, whereas having siblings was positively associated with secondary BAs concentration at 6 and 14 months (Supplementary Figures 49, 50). It seems that factors related to optimal microbiome development, especially breastfeeding, associated with fecal metabolite concentrations.

**Figure 2.**
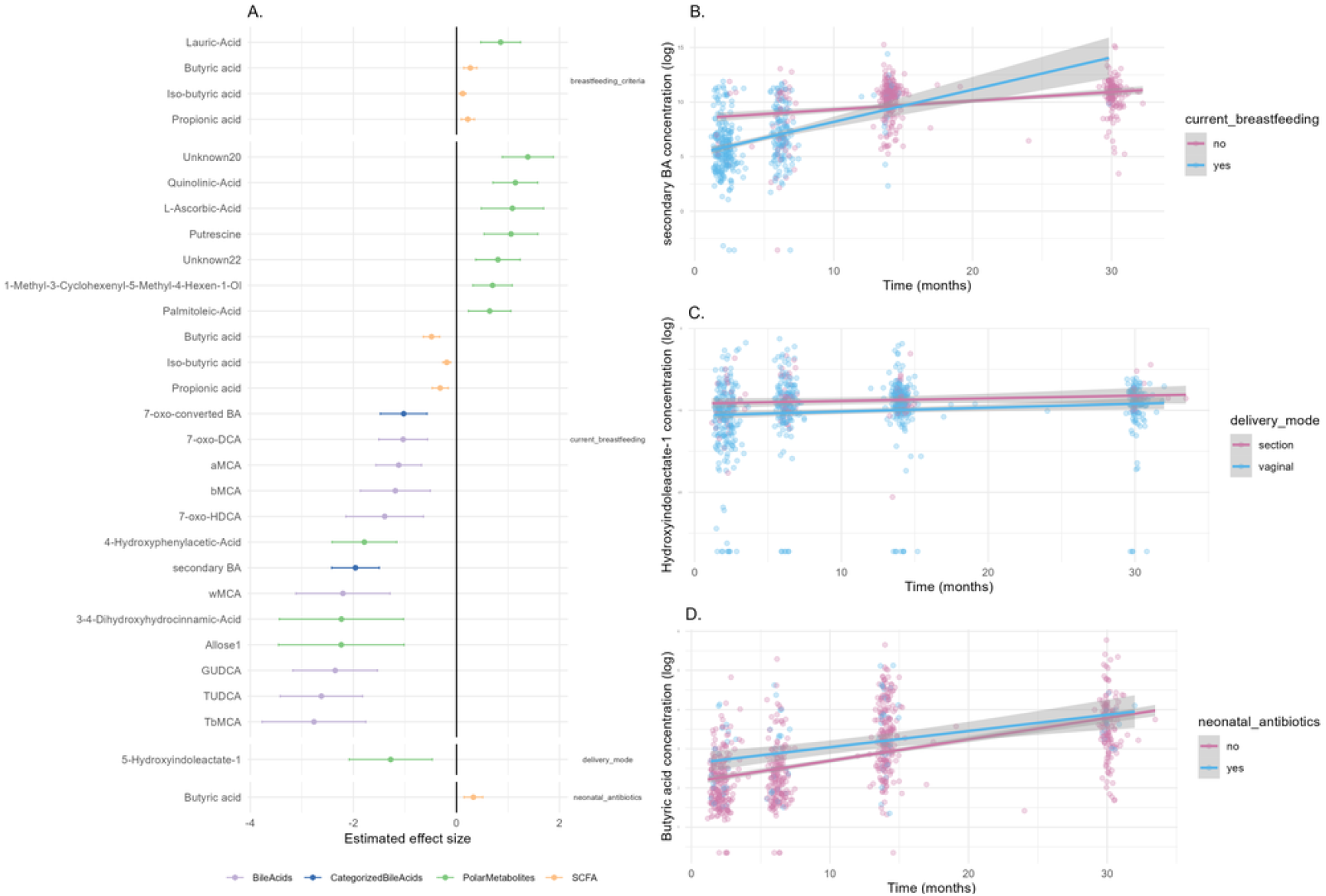
A. Estimates for each demographic variable from age-adjusted mixed model. B. Secondary BA concentrations were lower among breastfed infants in the 2.5-6 month timepoints. C. Vaginally born infants had consistently lower concentration of hydroxyindoleacetate-1 across all timepoints. D. Concentration of butyric acid was higher in infants who received antibiotic treatment in the neonatal period.

### Microbiome age-trends and association with demographic exposures

Previous literature has suggested a successional development of infant gut microbiota taxonomic composition, and we wanted to confirm these patterns in our data. To examine the patterns of gut microbiome succession during early-life we performed Dirichlet Multinomial Mixture (DMM) clustering. We identified 7 community types according to Laplace criteria when jointly analysing the samples from all time points. The first timepoint was dominated by three community types, that were driven by the abundances of *Bacteroides* and *Bifidobacterium* (C1), *Escherichia* (C2), *Veillonella*, and an unidentified genus in *Enterobacriaceae* (C3), (Figure 2 and Supplementary Figure 52). The majority of the later timepoints were dominated by a single community type that were driven by *Bacteroides, Clostridium* or *Veillonella* with differing proportions (C4-7, Figure 3, Supplementary figure 52).

**Figure 3.**
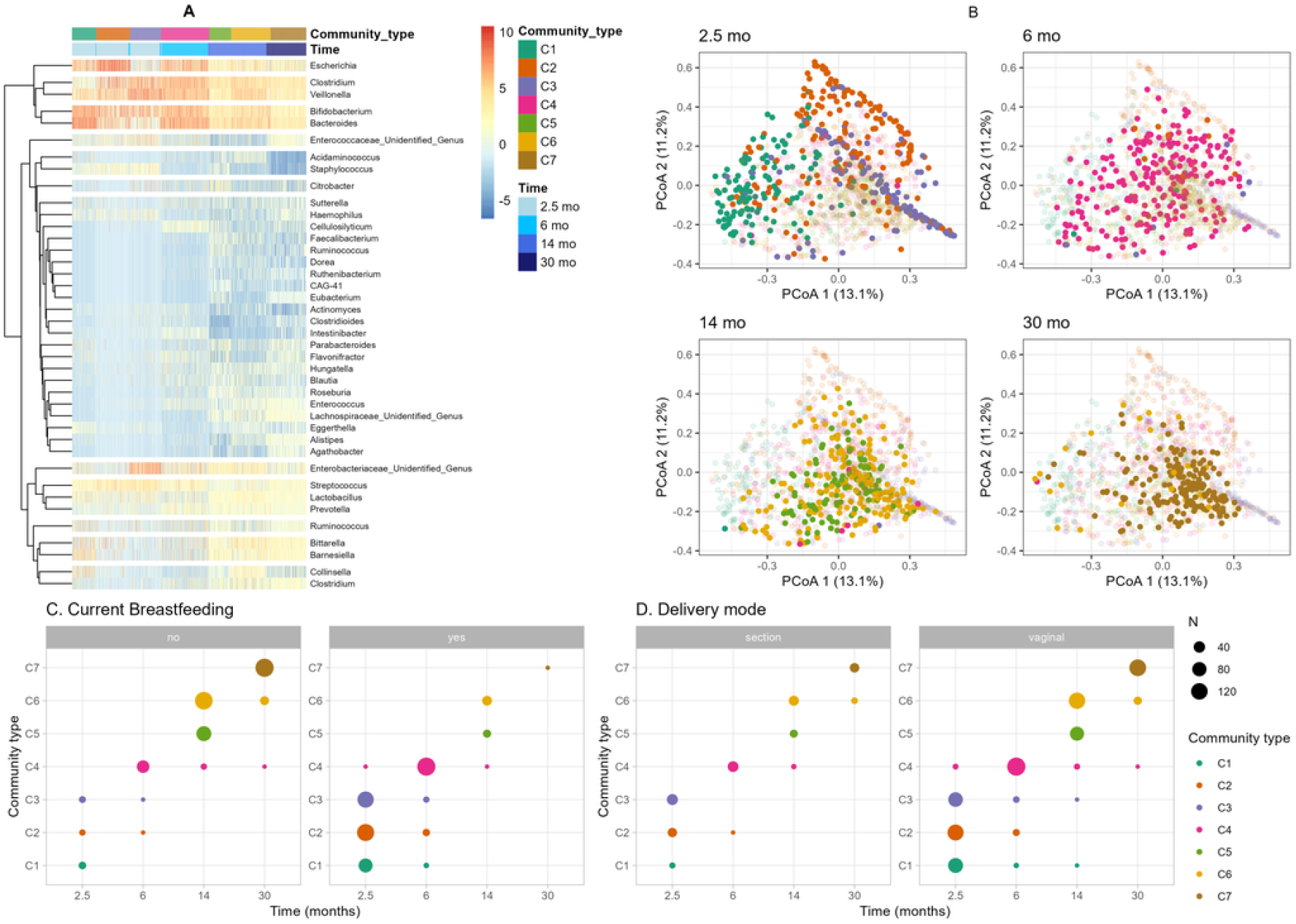
A. We identified seven community types. These abundances visualized in PCoA plot shows that although there is aggregation of clusters, there is no clear separation between those. However, it seems that C7 was the most homogenous as indicated by DMM theta (Supplementary table 4). In a mixed model, C. current breastfeeding and D. delivery mode explained transition between clusters.

Consistently with previous reports, the gut microbe community differed according to the background factors, including delivery mode and breastfeeding (Figure 3). Some additional trends were consistent with earlier reports but did not reach statistical significance. On the other hand, infant sex, having pets, and intravenous neonatal or recent antibiotic intake was not associated with gut microbe community membership. When stratified by timepoint, delivery mode at 2.5 months (C1 3.3%, C2 15.1%, C3 29.4% of C-section born infants, Χ^2^ q < 0.005, Supplementary figure 53) and preterm delivery at 6 months (C1 50%, C2 89%, C3 67%, C4 98%, Supplementary figure 53) were enriched in a specific community type(Supplementary tables 5-7), whereas perinatal (2.5 mo, 30 mo) and recent antibiotic treatments (6 mo, 30 mo), siblings (2.5 mo, 14 mo) were not significant (Supplementary Table 5). In a mixed model, the vaginal delivery and current breastfeeding were negatively related to community type progression (Fig 2C,D).

### Associations between metabolite concentrations and microbiome

Next, we sought to determine whether the microbiome composition associated with metabolome profiles. We found microbiota alpha diversity was correlated with multiple metabolite classes (Fig 5), in particular, SCFA concentration showed consistently positive associations with alpha diversity. The observed richness was also associated positively with SCFA concentration (Supplementary figure 57). Linear mixed model showed that THCA, TωMCA and several polar metabolites (Arachidonic acid, 2-Methylpentadecanoic acid, Putrescine) were negatively associated with Shannon Index (q < 0.015, Fig. 4), whereas butyric, propionic, isovaleric and iso-butyric acid, ωMCA, αMCA, UDCA, and other polar metabolite concentrations were positively associated with Shannon index when adjusted for age (p<0.039, Fig. 4). In addition, we found Shannon index was positively associated with 7-oxo-converted BA concentrations (estimate = 0.3, 95%-CI 0.3-0.56, q = 0.047). *Clostridium* and *Bifidobacterium* showed associations with butyric acid and conjugated BAs in opposing directions (Supplementary figure 55).

**Figure 4.**
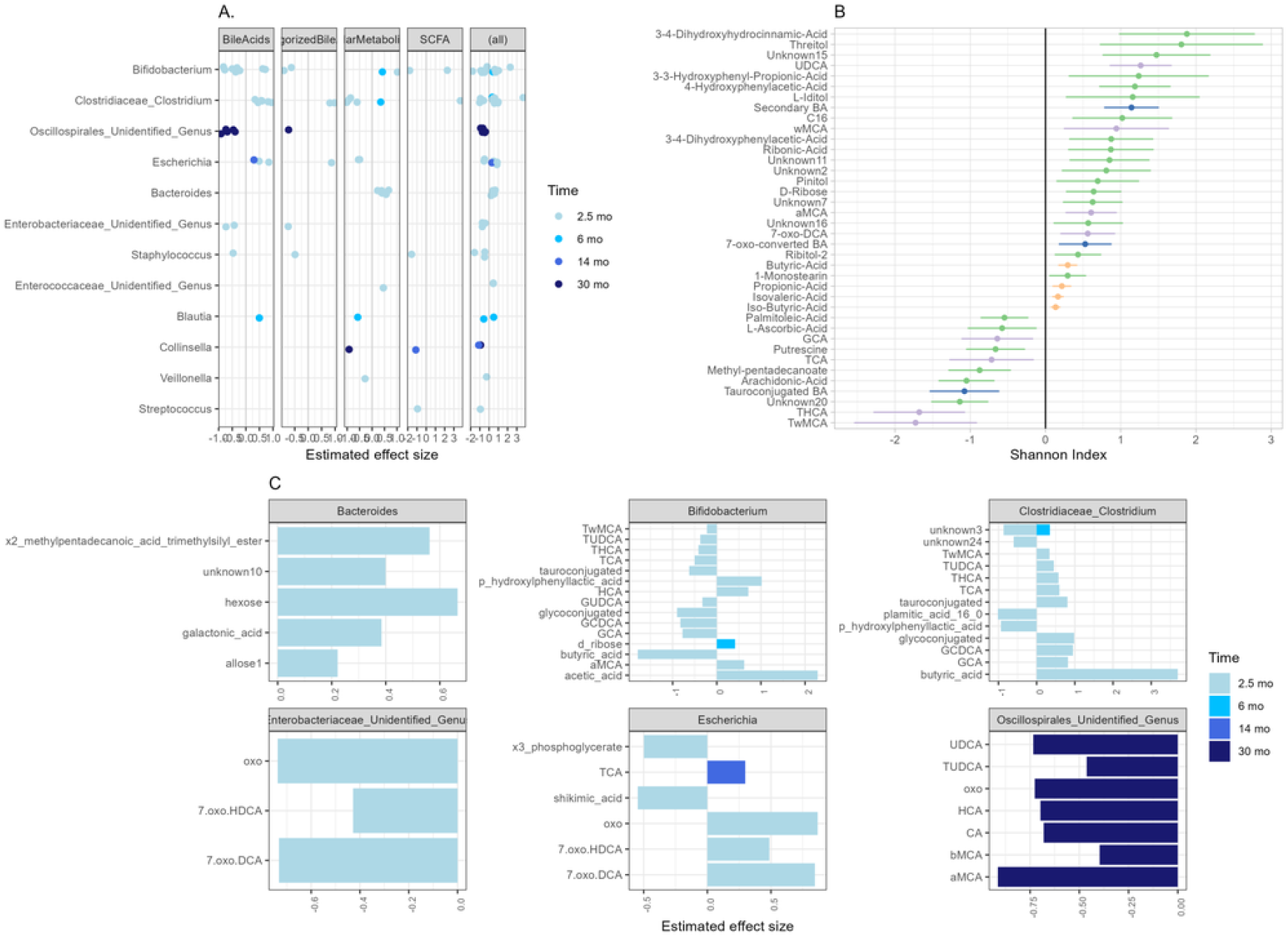
A. Differential abundance analysis showed multiple associations between genus abundances and metabolites (with the ALDEx2 method). Only significant associations (q < 0.05) are visualized. Across all timepoints, *Bifidobacterium* (n=22), *Clostridium* (n=18), unidentified genus in Oscillaspirales (n=11), *Bacteroides* (n=9), *Escherichia* (n=9) had most significant associations. B. SCFA tended to positively correlate with alpha diversity, whereas individual polar metabolites and BAs correlated both negatively and positively. C. Most significant associations between genera and metabolites were at 2.5 mo timepoint. *Bifidobacterium* at 2.5 mo associated negatively and *Clostridium* at 2.5 mo associated positively with conjugated BAs and butyric acid. *Streptococcus* was associated negatively with propionic acid. Unidentified genus in Oscillospiroles at 30 months associated negatively with multiple BAs, especially 7-oxo-converted and tauroconjugated BAs.

**Figure 5.**
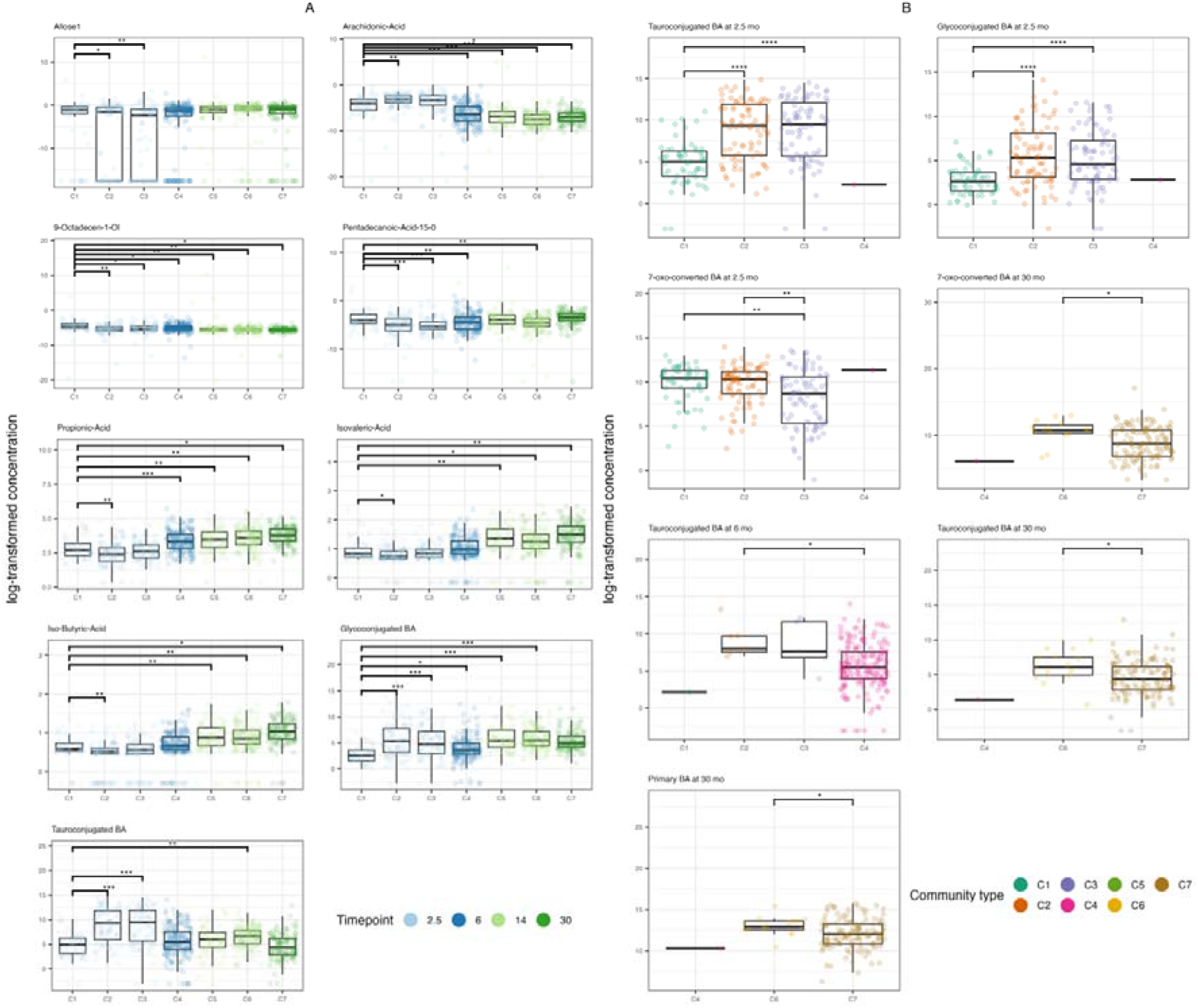
Cluster showed different levels of metabolites. A. Several associations remained after adjusting for breastfeeding in the mixed model, color indicating the timepoint. B. Clusters at 2.5, 6 and 30 months had different levels of BA based on cross-sectional group comparison and post hoc testing. * q < 0.05 & q > 0.01, ** q <= 0.01 & q > 0.001, ***q <= 0.001.

**Figure 6.**
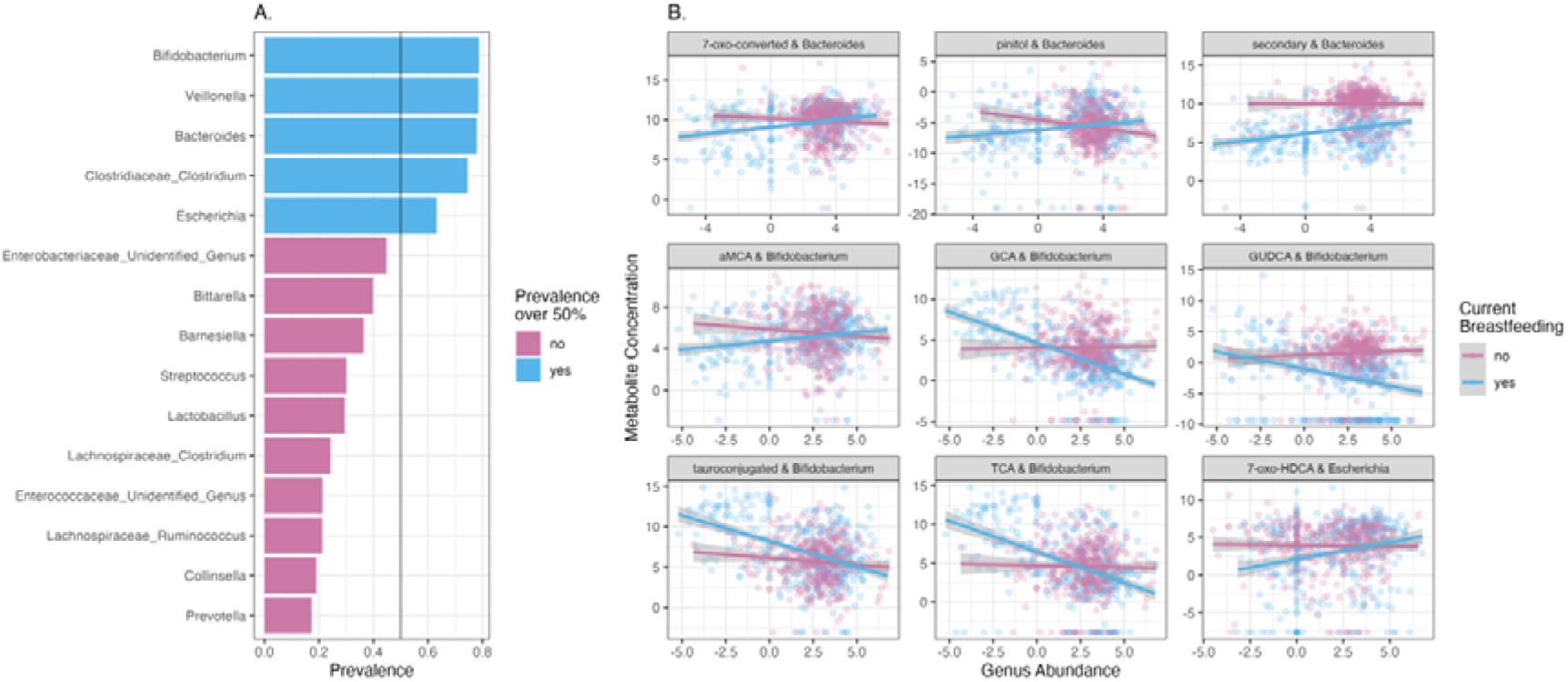
A. Only top 5 most prevalent taxa were observed in >50 % of the study subjects, and those selected for the interaction analyses. B. *Escherichia, Bifidobacterium* and *Bacteroides* showed significant interaction with breastfeeding. Scatterplots for significant interaction models.

#### Microbiota clusters associate with different levels of metabolites

Furthermore, we utilized the microbiota clusters to study whether metabolite concentrations were different between community types. Clusters showed different levels of fecal metabolites per timepoint, and largest effect sizes were for TwMCA, TCA, THCA, GCA as well as succinic acid and an unknown polar metabolite at 2.5 months (Supplementary Table 8). For all the above mentioned BAs, C1 had a lower concentration compared with C2 and/or C3 (Supplementary Table 8). Additionally, both glucoconjugated and tauroconjugated BA concentrations were lower in C1 at 2.5 months (Supplementary Figure 65, 66). At 14 months, butyric acid concentration was higher in C6 compared with C5. Additionally, at 30 months, C7 had higher concentrations of valeric acid, βMCA, succinic acid and βMCA with moderate effect size (Supplementary figures 62, 63).

The BAs TωMCA, THCA, TCA, GCA and arachidonic acid showed positive association with cluster membership. Whereas multiple polar metabolites, UDCA, propionic acid, and branched SCFA showed negative associations with cluster membership (Fig. 5). Likewise, glycoconjugated and tauroconjugated were both positively associated with clusters C2-C6 and C2, C3 and C6, respectively (FDR < 0.05, C1 as reference, Fig. 5).

#### The association between levels of metabolites and their interaction with breastfeeding, as well as with the microorganisms capable of metabolizing them

Breastfeeding drives the microbiome maturation. We observed that breastfeeding showed the strongest associations with metabolite levels. Thus, we wanted to further explore the interactions between gut microbes abundances, metabolite levels and breastfeeding. As the prevalent genera were driving the clusters and they showed the most associations metabolite levels, we studied how the interaction between prevalent genera and breastfeeding status associated with metabolite levels (Supplementary table 9.).

We observed that *Bifidobacterium* abundances were associated negatively with tauroconjugated BA concentration only in breastfed infants. On the other hand, *Bacteroides* was positively associated with 7-oxo-converted and secondary BA in the breastfed infants. Moreover, the less there is *Escherichia* in breastfed infants gut microbiota, the less there is 7-oxo-HDCA. Of the polar metabolites, *Bacteroides* abundances were positively associated with pinitol concentrations in the breastfed infants.

## Discussion

Gut microbiota undergoes successional development in early life^22^, which is affected by factors such as breastfeeding and delivery mode^4^. However, less is known about development of fecal metabolites, which are important mediators of physiological effects of the gut microbiome. Here, we showed in our population-based cohort that the fecal metabolome develops alongside the gut microbiome, and individual variation in microbiome is associated with the metabolome composition. Additionally, our observations suggest that breastfeeding, an important microbiome-modulating factor, is related to metabolite concentration depending on gut microbiota composition. This not only shows that metabolome is related to microbiome development, but that common exposures may have individualized effects based on microbiome composition.

SCFAs, except acetic acid, were systematically increased by age and this might be explained by more complex microbiota and increased intake of indigestible fiber by age. This is in agreement with earlier studies, which suggest an increasing stool SCFA trend after birth^23^ with exception for acetic acid. We observed no significant age-related increase for acetic acid, which might relate to the lack of very early sampling in our study. On the other hand, developmental patterns of BAs were more nuanced. Secondary BAs increased by age, whereas primary and tauroconjugated BAs decreased by age, which is partially in line with our previous findings^24^. The decrease in BAs could be related to increased bile salt hydrolase (BSH) activity, potentially driven by increasing abundances of *Clostridium* and *Bacteroides*^*25*^. Interestingly, glycoconjugated BAs were not increased by age when adjusting for breastfeeding. This could be explained by the observation that most of the infants in the first time point were breastfed, and thus harbored more bifidobacteria, which often have BSH enzymes with preference for glycine as a substrate over taurine^26^. Thus, it may be that the *Bifidobacterium*-dominated microbiome is already capable of deconjugating glycine in earlier phases, which is further supported by our observation that *Bifidobacterium* was negatively associated with glycoconjugated BA concentration.

We observed that breastfeeding was related to lower abundances of butyric, iso-butyric and propionic acid, which is in contrast to Brink et al. report^26^. However, we noted a negative association between *Bifidobacterium* and butyric acid, which corroborates a finding by Nguyen et al.. As noted by them, certain *Bifidobacterium* strains can compete for the same substrates as butyrate producers^19,27^, and thus the strain-level variation between the studies may underline the discrepancies in the reports. Our data would suggest that if a breastfed infants has a lower *Bifidobacterium* or higher *Bacteroides* abundance, there is a concomitant higher concentration of microbially modified BAs. Both *Bacteroides* and *Bifidobacterium* are hallmark genera of breastfed infants gut ecosystem, and those harbour differential capacity for BA metabolism. We acknowledge that strain level information is missing in our study. Notwithstanding that, we corroborate that secondary BA concentration was lower in breastfed infants^28^, which might reflect slower acquisition of microbiome with BA metabolizing capacity.

In line with previous reports, we observed clustering in the gut microbiome that mostly aligned with age reflecting typical colonization patterns in early-life. Notable exception was the 2.5 months’ timepoint when most infants were breastfed, where three community types with either *Bifidobacterium* and *Bacteroides, Veillonella* and *Enterobacteriaceae* or *Escherichia* dominance were observed. *Bifidobacterium* and *Bacteroides* dominated community type was related to lower rate of C-section, which aligns with existing literature^4,29^. Moreover, in addition to vaginal delivery, breastfeeding was related to a slower community type progression, which may indicate slower maturation of gut microbiota^4^. Thus, our data would support the observation that cessation of breastfeeding would result in faster maturation of gut microbiota.

The differences in metabolite concentrations between community types in the 2.5-month-olds further elucidates the interaction between microbiomes, factors affecting colonization and metabolites. The *Bifidobacterium* and *Bacteroides* dominated community type, which had a higher proportion of vaginally born infants, was associated with lower concentration of conjugated BAs than the two other major clusters in the first time point, most likely reflecting differences in BSH enzymatic activity. On the other hand, *Bifidobacterium* and *Bacteroides* dominated community type had higher concentration of propionic acid and branched SCFA iso-butyric and iso-valeric acids than the *Escherichia*-dominated community type, which may indicate increased availability of protein for microbial fermentation. The difference may relate to variation in human milk composition^30^, as no difference in breastfeeding was observed between community typedominating the first timepoint.

It is evident that breastfeeding is an essential factor in determining the microbiome. However, there is variation in the individual colonization patterns also in the breastfed infants, and we wanted to explore how the interaction between breastfeeding and prevalent taxa is associated with metabolite concentrations. Not surprisingly, conjugated BA concentrations were lower in the breastfed infants the more they had *Bifidobacteria*. On the other hand, breastfed infants with high *Bacteroides* abundances had higher concentrations of secondary BAs. This indicates a complex interaction between early nutrition, early life microbiome and microbially metabolized products. Thus, future focus on human milk components that potentially relate to the colonization patterns in microbiome when serving as substrates for microbial fermentation is warranted.

BAs participate to regulation of inflammatory and metabolic processes via farnesoid X receptor and other bile acid-responsive receptors. For instance, secondary BAs, more abundant in breastfed infants with high *Bacteroides*-levels, may inhibit pro-inflammatory processes in microglia^31^, and they are also required to activate vitamin D receptor to support optimal growth and development of adaptive immunity^32,33^. Early-life microbiome-bile acid crosstalk may then participate in programming of growth and later brain health. However, it is uncertain how exactly the complex feedback systems affect the physiological outcomes, since gut metabolites shape the postnatal gut microbiota composition^18^, and for instance tauroconjugated BAs metabolized by gut bacteria may in feedback inhibit BA synthesis via FXR antagonism^34^.

Although our study benefits from a large sample of children and a representative variation in breastfeeding and delivery mode, our sample collection time points do not extend to the neonatal time nor was the sampling dense. This may have limited us to detect more nuanced patterns in the colonization and metabolome development. The utilized 16s rRNA sequencing data provided important information on the overall microbiome profiles, but the results call for future studies focusing on gene-level differences in gut microbiome. Leveraging metagenomic sequencing in future studies will help to disentangle the role of BA metabolizing capacity in the developing gut microbiome. Moreover, more detailed data on early diet, such as analysis of human milk composition, may also help to describe the differences in microbiome composition and the functional output especially in breastfed infants. Future integration of the reported exploratory findings to mechanistic models will help to elucidate the clinical potential related to inflammation^32,35^ and metabolic programming^25^.

## Conclusion

First, we showed that SCFA concentrations, except acetic acid, increase within the first 30 months. Second, breastfeeding, among the background factors known to influence gut microbiota maturation, associated with multiple metabolites. Interestingly, the secondary BA concentrations were lower in the breastfed infants. Third, we corroborated that gut microbiota shows successional maturation during the first 30 months of life. Fourth, we showed that prevalent gut microbe abundances are associated with metabolite levels, especially in the 2.5-month-olds. Finally, we demonstrate that the prevalent early colonizers *Bacteroides, Escherichia* and *Bifidobacterium* abundances associate with the microbial metabolites BAs especially in the breastfed infants. Alterations in early-life bile acid-microbiota crosstalk may in future studies prove important mechanism in developmental programming of health. Breastfeeding and human milk composition are likely to be important moderators in the process.

## Methods

### Cohort description and data collection

The study subjects are children from the FinnBrain Cohort Study^36^ that is a general population birth cohort study located in the southwestern Finland. The FinnBrain Birth Cohort Study recruited families with sufficient fluency in Finnish or Swedish, and normal 1st trimester ultrasound examination. A subset of the cohort participated in the study visits, and there were no exclusion criteria for the collection of fecal samples. The initial recruitment took place between December 2011 and April 2015, and fecal samples were collected from May 2013 to May 2018. The fecal samples were collected from the children by the parents according to written and oral instructions at 2.5, 6, 14 and 30 months postpartum. The samples were collected in plastic tubes, and parents were instructed to store the sample in a refrigerator, and bring the sample to the laboratory within 24 h. The sample collection time was reported.

Clinical data used in the study were collected with parental reports during and after pregnancy at 14, 24, 34 gestational weeks, 3, 6, 12, and 24 months postpartum and during study visits (2.5, 6, 14, and 30 months). Likewise, the data on maternal pre-pregnancy body mass index (BMI; kg/m2), duration of gestation as well as mode of delivery (caesarian section vs. vaginal) were collected from National Birth Registry provided by the National Institute for Health and Welfare of Finland (www.thl.fi). The information on maternal perinatal and infant neonatal intravenous antibiotic intake was collected from the hospital records. Breastfeeding was categorized in two ways: 1) any current breastfeeding (yes vs. no); 2) exclusive breastfeeding at least 4 months and partial breastfeeding for at least 6 months (breastfeeding_criteria, yes vs. no).

The Ethical Committee of Southwestern Finland approved the study. Parents provided informed consent on behalf of their children. STORMS guideline was used for reporting the methods and materials.

### Metabolome analysis

The BAs were measured in fecal samples as described previously^16^. Only samples frozen within 24 h of sample collection were included in the metabolome analyses. The order of the samples was randomized before sample preparation. Two aliquots (50 mg) of each fecal sample were weighed. An aliquot was freeze-dried prior to extraction to determine the dry weight.. The second aliquot was homogenized by adding homogenizer beads and 20 μL of water for each mg of dry weight in the fecal sample, followed by samples freezing to at least -70 °C and homogenizing them for five minutes using a bead beater. The BAs analysed were Litocholic acid (LCA), 12-oxo-litocholic acid(12-oxo-LCA), Chenodeoxycholic acid (CDCA), Deoxycholic acid (DCA), Hyodeoxycholic acid (HDCA), Ursodeoxycholic acid (UDCA), Dihydroxycholestanoic acid (DHCA), 7-oxo-deoxycholic acid (7-oxo-DCA), 7-oxo-hyocholic acid (7-oxo-HCA), Hyocholic acid(HCA), β-Muricholic acid (b-MCA), Cholic acid (CA), Ω/α-Muricholic acid (w/a-MCA), Glycolitocholic acid (GLCA), Glycochenodeoxycholic acid (GCDCA), Glycodeoxycholic acid (GDCA), Glycohyodeoxycholic acid (GHDCA), Glycoursodeoxycholic acid (GUDCA), Glycodehydrocholic acid (GDHCA), Glycocholic acid (GCA), Glycohyocholic acid (GHCA), Taurolitocholic acid (TLCA), Taurochenodeoxycholic acid (TCDCA), Taurodeoxycholic acid (TDCA), Taurohyodeoxycholic acid (THDCA), Tauroursodeoxycholic acid (TUDCA), Taurodehydrocholic acid (TDHCA), Tauro-α-muricholic acid (TaMCA), Tauro-β-muricholic acid (TbMCA), Taurocholic acid (TCA), Trihydroxycholestanoic acid (THCA) and Tauro-Ω-muricholic acid (TwMCA). BAs were extracted by adding 40 μL fecal homogenate to 400 μL crash solvent (methanol containing 62,5 ppb each of the internal standards LCA-d4, TCA-d4, GUDCA-d4, GCA-d4, CA-d4, UDCA-d4, GCDCA-d4, CDCA-d4, DCA-d4 and GLCA-d4) and filtering them using a Supelco protein precipitation filter plate. The samples were dried under a gentle flow of nitrogen and resuspended using 20 μL resuspenstion solution (Methanol:water (40:60) with 5 ppb Perfluoro-n-[13C9]nonanoic acid as in injection standard). Quality control (QC) samples were prepared by combining an aliquot of every sample into a tube, vortexing it and preparing QC samples in the same way as the other samples. Blank samples were prepared by pipetting 400 μL crash solvent into a 96-well plate, then drying and resuspending them the same way as the other samples. Calibration curves were prepared by pipetting 40 μL of standard dilution into vials, adding 400 μL crash solution and drying and resuspending them in the same way as the other samples. The concentrations of the standard dilutions were between 0.0025 and 600 ppb.

The LC separation was performed on a Sciex Exion AD 30 (AB Sciex Inc., Framingham, MA) LC system consisting of a binary pump, an autosampler set to 15 °C and a column oven set to 35 °C. A waters Aquity UPLC HSS T3 (1.8μm, 2.1×100mm) column with a precolumn with the same material was used. Eluent A was 0.1 % formic acid in water and eluent B was 0.1 % formic acid in methanol. The gradient started from 15 % B and increased to 30 % B over 1 minute. The gradient further increased to 70 % B over 15 minutes. The gradient was further increased to 100 % over 2 minutes. The gradient was held at 100 % B for 4 minutes then decreased to 15 % B over 0.1 minutes and re-equilibrated for 7.5 minutes. The flow rate was 0.5 mL/min and the injection volume was 5 μL.

The mass spectrometer used for this method was a Sciex 5500 QTrap mass spectrometer operating in scheduled multiple reaction monitoring mode in negative mode. The ion source gas1 and 2 were both 40 psi. The curtain gas was 25 psi, the CAD gas was 12 and the temperature was 650 °C. The spray voltage was 4500 V. Data processing was performed on Sciex MultiQuant.

#### Quantification of SCFA

We adapted and modified the targeted SCFA analysis from previous work^37^. Fecal samples were homogenized by adding water (10 μL per mg of dry weight as determined for the BA analysis) to wet feces, the samples were homogenized using a bead beater. Analysis of SCFA was performed on fecal homogenate (50 μL) crashed with 500 μL methanol containing internal standard (propionic acid-d6 and hexanoic acid-d3 at 10 ppm). Samples were vortexed for 1 min, followed by filtration using 96-Well protein precipitation filter plate (Sigma-Aldrich, 55263-U). Retention index (RI, 8 ppm C10-C30 alkanes and 4 ppm 4,4-Dibromooctafluorobiphenyl in hexane) was added to the samples. Gas chromatography (GC) separation was performed on an agilent 5890B GC system equipped with a Phenomenex Zebron ZB-WAXplus (30 m × 250 μm × 0.25 μm) column a short blank pre-column (2 m) of the same dimensions was also added. A sample volume of 1 μL was injected into a split/splitless inlet at 285°C using split mode at 2:1 split ratio using a PAL LSI 85 sampler. Septum purge flow and split flow were set to 13 mL/min and 3.2 mL/min, respectively. Helium was used as carrier gas, at a constant flow rate of 1.6 mL/min. The GC oven program was as follows: initial temperature 50°C, equilibration time 1 min, heat up to 150°C at the rate of 10°C/min, then heat at the rate of 40°C/min until 230°C and hold for 2 min. Mass spectrometry was performed on an Agilent 5977A MSD. Mass spectra were recorded in Selected Ion Monitoring (SIM) mode. The detector was switched off during the 1 min of solvent delay time. The transfer line, ion source and quadrupole temperatures were set to 230, 230 and 150°C, respectively. Dilution series of SCFA standards of acetic, propionic, butyric, valeric, hexanoic acid, isobutyric, and iso-valeric acid were prepared in concentrations of 0.1, 0.5, 1, 2, 5, 10, 20, 40, and 100 ppm for the construction of standard curves for quantification.

#### Analysis of polar metabolites

Polar metabolites were extracted in methanol. The method was adapted from the method used by Lamichhane et al.^24^. Fecal homogenate (60 μL) were diluted with 600 μL methanol crash solvent containing internal standards (heptadecanoic acid (5 ppm) valine-d8 (1 ppm) and glutamic acid-d5 (1 ppm)). After precipitation the samples were filtered using Supelco protein precipitation filter plates. One aliquot (50 μL) was transferred to a shallow 96-well plate to create a QC sample. The rest of the sample volume was dried under a gentle stream of nitrogen and stored in -80 °C until analysis. After thawing the samples were again dried to remove any traces of water. Derivatization was carried out on a Gerstel MPS MultiPurpoe Sampler using the following protocol: 25 μL methoxamine (20 mg/mL) was added to the sample followed by incubation on a shaker heated to 45 °C for 60 minutes. N-Methyl-N-(trimethylsilyl) trifluoroacetamide (25 μL) was added followed by incubation (60 min). After that, 25 μL retention index was added, the sample was allowed to mix for one min followed by injection. The automatic derivatization was carried out using the Gerstel maestro 1 software (version 1.4).

Gas chromatographic (GC) separation was carried out on an Agilent 7890B GC system equipped with an Agilent DB-5MS (20 m x 0,18 mm (0,18 μm)) column. A sample volume of 1 μl was injected into a split/splitless inlet at 250°C using splitless mode. The system was guarded by a retention gap column of deactivated silica (internal dimensions 1.7 m, 0.18 mm, PreColumn FS, Ultimate Plus Deact; Agilent Technologies, CA, USA). Helium was used as carrier gas at a flow rate of 1.2 ml/min for 16 min followed by 2 mL/min for 5.75 min. The temperature programme started at 50°C (5 min), then a gradient of 20°C/min up to 270°C was applied and then finally a gradient of 40°/min to 300°C, where it was held stable for 7 min. The mass spectrometry was carried out on a LECO Pegasus BT system (LECO). The acquisition delay was 420 sec. The acquisition rate was 16 spectra/sec. The mass range was 50 – 500 m/z and the extraction frequency was 30 kHz. The ion source was held at 250 °C and the transferline heater temperature was 230 °C. ChromaTOF software (version 5.51) was used for data aquisition. The samples were run in 9 batches, each consisting of 100 samples and a calibration curve. In order to monitor the run a blank, a QC and a standard sample with a known concentration run between every 10 samples. Between every batch the septum and liner on the GC were replaced, the precolumn was cut if necessary and the instrument was tuned.

The retention index was determined with ChromaTOF using the reference method function. For every batch a reference file was created. The reference file contained the spectras and approximate retention times of the alkanes from C10 to C30 as determined manually). A reference method was implemented for every sample in order to determine the exact retention time of the alkanes. Text files with the names and retention times of the alkanes were then exported and converted to the correct format for MSDIAL using an in-house R script. The samples were exported from ChromaTOF using the netCDF format. After this they were converted to abf files using the abfConverter software (Reifycs). Untargeted data processing was carried out using MSDIAL (version 4.7). The minimum peak height was set to an amplitude of 1000, the sigma window value was 0.7 and the EI spectra cut off was 10. The identification was carried out using retention index with the help of the GCMS DB-Public-kovatsRI-VS3 library provided on the MSDIAL webpage. A separate RI file was used for each sample. The RI tolerance was 20 and the m/z tolerance was 0.5 Da. the EI similarly cut off was 70 %. The identification score cut off was 70 % and retention information was used for scoring. Alignment was carried out using the RI with an RI tolerance of 10. The EI similarity tolerance was 60 %. The RI factor was 0.7 and the EI similarity factor was 0.5. The results were exported as peak areas and further processed with excel. In excel the results were normalized using heptadecanoic acid as internal standard and the features with a coefficient of variance of less than 30 % in QC samples were selected. Further filtering was carried out to remove alkanes and duplicate features. The IDs of the features which passed the CV check were further checked using the Golm Metabolome Database.

### Microbiome analysis

#### DNA extraction and sample processing

The samples were divided into cryotubes and freezed in -80C within 2 days after arriving at the laboratory. Samples were kept at +4C before freezing. Only samples that were freezed within 48 h of sample collection were sequenced. Sample volume for DNA extraction was approximately 100 mg. Lysis buffer was added 1 ml and the samples were homogenized with glass beads 1000 rpm / 3 min. The samples were centrifuged at high speed (> 13000 rpm) for 5 min. The lysate (800μL) was then transferred to tubes and the extraction proceeded according to the manufacturer’s protocol. DNA was extracted using a semi-automatic extraction instrument Genoxtract with DNA stool kit (HAIN life science, Germany).

DNA yields were measured with Qubit fluorometer using Qubit dsDNA High Sensitivity Assay kit (Thermo Fisher Scientific, USA). The DNA extraction and sequencing was performed in the University of Turku.

#### 16S ribosomal RNA (rRNA) amplicon sequencing

Bacterial community composition was determined by sequencing the V4 region of 16S rRNA gene using Illumina MiSeq platform (Illumina, USA). The sequence library was constructed with an in-house developed protocol where amplicon PCR and index PCR were combined..

The DNA samples were diluted in PCR grade water to 10 ng/μL concentration prior to library PCR. PCR was performed with KAPA HiFi High Fidelity PCR kit with dNTPs (Roche, USA). Reverse and forward primers included in-house modifications verified by Rintala et al.^38^. The forward and reverse primer sequences were 5’-AATGAT-ACGGCGACCACCGAGATCTACAC -i5-TATGGTAATT -GT-GTGCCAGCMGCCGCGGTAA-3’ and 5’-CAAGCAGAAGACGGCATACGAGAT -i7-AGTCAGTCAG-GC-GGACTACHVGGGTWTCTAAT-3’, respectively, where i5 and i7 indicate the sample specific indexes. After PCR, 5μl of the product was analyzed with 1,5% TBE agarose gel (100V, 1h15min). PCR products were purified with AMPure XP magnetic beads (Becman Coulter, USA). The DNA concentration of the purified samples were measured with Qubit fluorometer using Qubit dsDNA High Sensitivity Assay kit (Thermo Fisher Scientific, USA), after which the samples were mixed in equimolar concentration into a 4 nM library pool. The library pool was denatured, diluted to a concentration of 4 pM and a denaturized PhiX control (Illumina, USA) was added. The sequencing was performed with Illumina MiSeq Reagent kit v3 (600 cycles) on MiSeq system with 2x 250 base pair (bp) paired ends following the manufacturer’s instructions. Positive control (DNA 7-mock standard) and negative control (PCR grade water) were included in library preparation and sequencing runs (Supplementary figures 5-8).

DADA2-pipeline (version 1.14) was used to preprocess the 16S rRNA gene sequencing data to infer exact amplicon sequence variants (ASVs)^39^. The reads were truncated to length 225 and reads with more than two expected errors were discarded (maxEE = 2). SILVA taxonomy database (version 138)^40,41^ and RDP Naive Bayesian Classifier algorithm^42^ were used for the taxonomic assignments of the ASVs.

### Statistical analyses

The data analyses were performed with R version 4.2.0 with packages including *phyloseq, mia, vegan, DirichletMultinomial* and *lme*. Heatmaps were created with the *pheatmap* R package. Shannon Index and Inverse Simpson were used as alpha diversity indices and those were calculated with *mia* package from the untransformed ASV-table. Metabolite concentrations were log-transformed with a pseudocount (minimum value / 2). Dirichlet Multinomial Mixtures model with the rarefied, genus-level count data were used for clustering the microbiome data. The number of community types was justified by the Laplace criteria.

Factor analysis, the relative contribution of a clinical/demographic factor towards the total variance of the metabolite classes were estimated by fitting a linear regression model. The total metabolite concentrations of a particular class was regressed to a clinical/demographic factor of interest, and median marginal coefficient of determination (R2) and % of explained variance were estimated. Factor analysis was performed using the ‘Scater’ package deployed in R.

Wilcoxon test, Chi-square test, and Kruskal-Wallis test with Dunn’s posthoc test were used in the analyses. Linear mixed models with child ID as random effect and sampling age as fixed effect was used to study i. metabolite age-trends, ii. association between metabolite concentrations and demographic factors, iii., association between *microbiome* community typemembership and demographic factors, iv. associations between metabolite concentrations and *microbiome* community type membership, and v. association between metabolite concentrations and the interaction with breastfeeding and rclr-transformed prevalent genera abundances as breastfeeding has been shown to drive the microbiome maturation^4^. Package lme4 was used to check for model singularity, and nlme was used for running the mixed model. The clr-module from the ALDEx2 package was used for the differential abundance analysis^43^. Variance explained in the metabolome assays by demographic factors was calculated with the package *scater*^*44*^. p-Values were adjusted for multiple testing with Benjamini-Hochberg procedure.

The analysis scripts and full results can be found in the supplementary files.

## Data Availability

Due to Finnish national legislation, the research data cannot be made available online, but data can potentially be shared with Material Transfer Agreement as part of research collaboration. Requests for collaboration can be sent to the Board of the FinnBrain Birth Cohort Study; please contact Hasse Karlsson and Linnea Karlsson. Source code for the data analysis is provided as a supplementary file.

## Ethics declarations

Ethical issues have been considered and there is a research permit for the project. FinnBrain has a permit from VSSHP ethical committee (ETMK: 57/180/2011), which has approved Cohort profile and research protocol (Karlsson et al. 2018). FinnBrain parents have signed a consent form about their children’s participation in research and given permission to use their samples for scientific purposes. Samples went through the laboratory process anonymously with research code to protect participants’ privacy.

## Funding

Finnbrain Birth cohort Study (HK) has been funded by Academy of Finland (grant numbers 253270, 134950), Jane and Aatos Erkko Foundation, as well as Signe and Ane Gyllenberg Foundation. LK was funded by the Academy of Finland (grant number 308176 and 325292), Yrjö Jahnsson Foundation (6847, 6976), Signe and Ane Gyllenberg Foundation, Finnish State Grants for Clinical Research (P3654), Jalmari and Rauha Ahokas Foundation, and Waterloo Foundation (2110-3601). AKA was supported by Yrjö Jahnsson Foundation, Psychiatry Research Foundation, Emil Aaltonen Foundation, Brain Foundation, Instrumentarium Science Foundation, Signe and Ane Gyllenberg Foundation, Duodecim Finnish Medical Society, Juho Vainio Foundation and Academy of Finland (grant number 347640). LL was supported by Academy of Finland (grant number 295741). EM was supported by the government research grant awarded to Turku University Hospital. AD has been funded by the Waterloo foundation and Finnish Academy (347924). “Inflammation in human early life: targeting impacts on life-course health” (INITIALISE) consortium funded by the Horizon Europe Program of the European Union under Grant Agreement 101094099 (to MO HK AD)

## Acknowledgements

We want to thank all the participating families and the FinnBrain staff and assisting personnel. Turku Metabolomics Centre and Biocenter Finland is acknowledged for the collaboration regarding fecal sample metabolomics.

## Conflicts of interest

EM is the medical advisor in Biocodex Finland. Other authors report no conflicts of interest.

## Author Contributions

Conception or design of the work: AKA, SL, AD, LL, ..

Acquisition, analysis, or interpretation of data: LK, HK, EM, HMK, HI, AK, LP, ML, MO, AD, AKA, SL, LL, MAA ..

Drafting or substantial revision of the work: AKA, SL, HI, AD, ..

All authors have approved the submitted revision.

## Notes

### Competing Interest Statement

Eveliina Munukka is the medical advisor in Biocodex Finland.
All other authors present no conflict of interest

### Author Declarations

Ethical issues have been considered and there is a research permit for the project. FinnBrain has a permit from VSSHP ethical committee (ETMK: 57/180/2011) which has approved Cohort profile and research protocol (Karlsson et al. 2018). FinnBrain parents have signed a consent form about their childrens participation in research and given permission to use their samples for scientific purposes. Samples went through the laboratory process anonymously with research code to protect participants privacy.

## References

1 Turnbaugh, P. J. et al. The human microbiome project. Nature 449, 804–810, doi:10.1038/nature06244 (2007).

2 O’Hara, A. M. & Shanahan, F. The gut flora as a forgotten organ. EMBO Rep 7, 688–693, doi:10.1038/sj.embor.7400731 (2006).

3 Backhed, F. et al. Dynamics and Stabilization of the Human Gut Microbiome during the First Year of Life. Cell Host Microbe 17, 852, doi:10.1016/j.chom.2015.05.012 (2015).

4 Stewart, C. J. et al. Temporal development of the gut microbiome in early childhood from the TEDDY study. Nature 562, 583–588, doi:10.1038/s41586-018-0617-x (2018).

5 Korpela, K. & de Vos, W. M. Early life colonization of the human gut: microbes matter everywhere. Curr Opin Microbiol 44, 70–78, doi:10.1016/j.mib.2018.06.003 (2018).

6 Franzosa, E. A. et al. Gut microbiome structure and metabolic activity in inflammatory bowel disease. Nat Microbiol 4, 293–305, doi:10.1038/s41564-018-0306-4 (2019).

7 Muscogiuri, G. et al. Gut microbiota: a new path to treat obesity. Int J Obes Suppl 9, 10–19, doi:10.1038/s41367-019-0011-7 (2019).

8 Cryan, J. F. et al. The Microbiota-Gut-Brain Axis. Physiol Rev 99, 1877–2013, doi:10.1152/physrev.00018.2018 (2019).

9 Andrioaie, I. M. et al. The Role of the Gut Microbiome in Psychiatric Disorders. Microorganisms 10, pdoi:10.3390/microorganisms10122436 (2022).

10 Robertson, R. C., Manges, A. R., Finlay, B. B. & Prendergast, A. J. The Human Microbiome and Child Growth - First 1000 Days and Beyond. Trends Microbiol 27, 131–147, doi:10.1016/j.tim.2018.09.008 (2019).

11 Tamburini, S., Shen, N., Wu, H. C. & Clemente, J. C. The microbiome in early life: implications for health outcomes. Nat Med 22, 713–722, doi:10.1038/nm.4142 (2016).

12 Rooks, M. G. & Garrett, W. S. Gut microbiota, metabolites and host immunity. Nat Rev Immunol 16, 341–352, doi:10.1038/nri.2016.42 (2016).

13 Vuong, H. E. et al. The maternal microbiome modulates fetal neurodevelopment in mice. Nature 586, 281–286, doi:10.1038/s41586-020-2745-3 (2020).

14 Sprockett, D., Fukami, T. & Relman, D. A. Role of priority effects in the early-life assembly of the gut microbiota. Nat Rev Gastroenterol Hepatol 15, 197–205, doi:10.1038/nrgastro.2017.173 (2018).

15 Zierer, J. et al. The fecal metabolome as a functional readout of the gut microbiome. Nat Genet 50, 790–795, doi:10.1038/s41588-018-0135-7 (2018).

16 Lamichhane, S., Sen, P., Dickens, A. M., Oresic, M. & Bertram, H. C. Gut metabolome meets microbiome: A methodological perspective to understand the relationship between host and microbe. Methods 149, 3–12, doi:10.1016/j.ymeth.2018.04.029 (2018).

17 Guzior, D. V. & Quinn, R. A. Review: microbial transformations of human bile acids. Microbiome 9, 140, doi:10.1186/s40168-021-01101-1 (2021).

18 van Best, N. et al. Bile acids drive the newborn’s gut microbiota maturation. Nat Commun 11, 3692, doi:10.1038/s41467-020-17183-8 (2020).

19 Nguyen, Q. P. et al. Associations between the gut microbiome and metabolome in early life. BMC Microbiol 21, 238, doi:10.1186/s12866-021-02282-3 (2021).

20 Jian, C. et al. Early-life gut microbiota and its connection to metabolic health in children: Perspective on ecological drivers and need for quantitative approach. EBioMedicine 69, 103475, doi:10.1016/j.ebiom.2021.103475 (2021).

21 Matharu, D. et al. Bacteroides abundance drives birth mode dependent infant gut microbiota developmental trajectories. Front Microbiol 13, 953475, doi:10.3389/fmicb.2022.953475 (2022).

22 Beller, L. et al. Successional Stages in Infant Gut Microbiota Maturation. mBio 12, e0185721, doi:10.1128/mBio.01857-21 (2021).

23 Xiong, J. et al. Development of gut microbiota along with its metabolites of preschool children. BMC Pediatr 22, 25, doi:10.1186/s12887-021-03099-9 (2022).

24 Lamichhane, S. et al. Dysregulation of secondary bile acid metabolism precedes islet autoimmunity and type 1 diabetes. Cell Rep Med 3, 100762, doi:10.1016/j.xcrm.2022.100762 (2022).

25 Wahlstrom, A., Sayin, S. I., Marschall, H. U. & Backhed, F. Intestinal Crosstalk between Bile Acids and Microbiota and Its Impact on Host Metabolism. Cell Metab 24, 41–50, doi:10.1016/j.cmet.2016.05.005 (2016).

26 Brink, L. R. et al. Neonatal diet alters fecal microbiota and metabolome profiles at different ages in infants fed breast milk or formula. Am J Clin Nutr 111, 1190–1202, doi:10.1093/ajcn/nqaa076 (2020).

27 Moens, F., Weckx, S. & De Vuyst, L. Bifidobacterial inulin-type fructan degradation capacity determines cross-feeding interactions between bifidobacteria and Faecalibacterium prausnitzii. Int J Food Microbiol 231, 76–85, doi:10.1016/j.ijfoodmicro.2016.05.015 (2016).

28 Khine, W. W. T. et al. Indonesian children fecal microbiome from birth until weaning was different from microbiomes of their mothers. Gut Microbes 12, 1761240, doi:10.1080/19490976.2020.1761240 (2020).

29 Reyman, M. et al. Impact of delivery mode-associated gut microbiota dynamics on health in the first year of life. Nat Commun 10, 4997, doi:10.1038/s41467-019-13014-7 (2019).

30 Borewicz, K. et al. The association between breastmilk oligosaccharides and faecal microbiota in healthy breastfed infants at two, six, and twelve weeks of age. Sci Rep 10, 4270, doi:10.1038/s41598-020-61024-z (2020).

31 Joo, S. S., Kang, H. C., Won, T. J. & Lee, D. I. Ursodeoxycholic acid inhibits pro-inflammatory repertoires, IL-1 beta and nitric oxide in rat microglia. Arch Pharm Res 26, 1067–1073, doi:10.1007/BF02994760 (2003).

32 Song, X. et al. Microbial bile acid metabolites modulate gut RORgamma(+) regulatory T cell homeostasis. Nature 577, 410–415, doi:10.1038/s41586-019-1865-0 (2020).

33 Ahmad, O., Nogueira, J., Heubi, J. E., Setchell, K. D. R. & Ashraf, A. P. Bile Acid Synthesis Disorder Masquerading as Intractable Vitamin D-Deficiency Rickets. J Endocr Soc 3, 397–402, doi:10.1210/js.2018-00314 (2019).

34 Sayin, S. I. et al. Gut microbiota regulates bile acid metabolism by reducing the levels of tauro-beta-muricholic acid, a naturally occurring FXR antagonist. Cell Metab 17, 225–235, doi:10.1016/j.cmet.2013.01.003 (2013).

35 Devkota, S. et al. Dietary-fat-induced taurocholic acid promotes pathobiont expansion and colitis in Il10-/-mice. Nature 487, 104–108, doi:10.1038/nature11225 (2012).

36 Karlsson, L. et al. Cohort Profile: The FinnBrain Birth Cohort Study (FinnBrain). Int J Epidemiol 47, 15–16j, doi:10.1093/ije/dyx173 (2018).

37 Trimigno, A. et al. Identification of weak and gender specific effects in a short 3 weeks intervention study using barley and oat mixed linkage beta-glucan dietary supplements: a human fecal metabolome study by GC-MS. Metabolomics 13, 108, doi:10.1007/s11306-017-1247-2 (2017).

38 Rintala, A. et al. Gut Microbiota Analysis Results Are Highly Dependent on the 16S rRNA Gene Target Region, Whereas the Impact of DNA Extraction Is Minor. J Biomol Tech 28, 19–30, doi:10.7171/jbt.17-2801-003 (2017).

39 Callahan, B. J. et al. DADA2: High-resolution sample inference from Illumina amplicon data. Nat Methods 13, 581–583, doi:10.1038/nmeth.3869 (2016).

40 Quast, C. et al. The SILVA ribosomal RNA gene database project: improved data processing and web-based tools. Nucleic Acids Res 41, D590–596, doi:10.1093/nar/gks1219 (2013).

41 Yilmaz, P. et al. The SILVA and “All-species Living Tree Project (LTP)” taxonomic frameworks. Nucleic Acids Res 42, D643–648, doi:10.1093/nar/gkt1209 (2014).

42 Wang, Q., Garrity, G. M., Tiedje, J. M. & Cole, J. R. Naive Bayesian classifier for rapid assignment of rRNA sequences into the new bacterial taxonomy. Appl Environ Microbiol 73, 5261–5267, doi:10.1128/AEM.00062-07 (2007).

43 Fernandes, A. D., Macklaim, J. M., Linn, T. G., Reid, G. & Gloor, G. B. ANOVA-like differential expression (ALDEx) analysis for mixed population RNA-Seq. PLoS One 8, e67019, doi:10.1371/journal.pone.0067019 (2013).

44 McCarthy, D. J., Campbell, K. R., Lun, A. T. & Wills, Q. F. Scater: pre-processing, quality control, normalization and visualization of single-cell RNA-seq data in R. Bioinformatics 33, 1179–1186, doi:10.1093/bioinformatics/btw777 (2017).

