## Supplement Information for "Dynamics of Gut Metabolome and Microbiome Maturation during Early Life"

Supplementary tables and figures.

Figures:

Figure 1. Richness and diversity in all timepoints

Figure 2. Library sizes in all timepoints and the whole sample.

Figure 3. Relative abundances of the 15 most abundant genera.

Figures 4-7 Read counts and relative abundances in control samples.

Figure 8. Estimates and 95 % confidence intervals for metabolites associated with age when adjusting for current breastfeeding.

Figure 9. Associations between demographic factors and metabolite concentrations per timepoint.

Figure 10. Secondary bile acid concentrations in breastfed and non-breastfed children per timepoint.

Figures 11-47. Metabolite concentrations and demographic factors box plots per timepoint. Only group differences with q < 0.05 are visualized.

Figures 48-51. Categorized bile acid concentrations and demographic factors per timepoint. Only group differences with q < 0.05 are visualized.

Figure 52. Contribution of taxonomic groups to the DMM clusters.

Figure 53. Differences in demographic factors in clusters per timepoint. Findings with p-value <0.05 are visualized.

Figure 54. Estimates for significant (q < 0.05) findings from ALDEx2 across all timepoints per genus.

Figure 55. ALDEx2 indicated multiple associations between bile acids concentrations and Bifidobacterium and Clostridium abundances at 2.5 mo. Interestingly, they were in opposing directions. Bile acid data is log-transformed whereas abundance data is robust centered log-transformed (rclr) for the visualization.

Figure 56. At 30 months unidentified genus in Oscillospirales order associated negative with bile acids, including 7-oxo-converted bile acids. Bile acid data is log-transformed whereas abundance data is robust centered log-transformed (rclr) for the visualization.

Figure 57. Estimates for richness in mixed-model adjusted for age.

Figure 58. Differences in bile acid concentrations between clusters per timepoint. Only results with q < 0.05 and Dunn’s test adjusted p-value < 0.05 are shown.

Figure 59. Differences in SCFA concentrations between clusters per timepoint. Only results with q < 0.05 and Dunn’s test adjusted p-value < 0.05 are shown.

Figure 60. Differences in polar metabolites concentrations between clusters at 2.5 months. Only results with q < 0.05 and Dunn’s test adjusted p-value < 0.05 are shown.

Figure 61. Differences in polar metabolites concentrations between clusters at 6 months. Only results with q < 0.05 and Dunn’s test adjusted p-value < 0.05 are shown.

Figure 62. Differences in polar metabolites concentrations between clusters at 14 months. Only results with q < 0.05 and Dunn’s test adjusted p-value < 0.05 are shown.

Figure 63. Differences in polar metabolites concentrations between clusters at 30 months. Only results with q < 0.05 and Dunn’s test adjusted p-value < 0.05 are shown.

Figure 64. Differences in categorized bile acid concentrations between clusters. Only results with q < 0.05 and Dunn’s test adjusted p-value < 0.05 are shown.

Figure 65. Tauroconjugated bile acids were lower in C1 compared with C2, C3 and C6 in mixed-models.

Figure 66. Glycoonjugated bile acids were lower in C1 compared with C2, C3, C4, C5 and C6 in mixed-models.

Tables:

Table 1. Sample characteristics.

Table 2. Genera prevalences. Genera with prevalence >= 1 % are presented.

Table 3. Variance explained on average by the demographic factors.

Table 4. Cluster pi and theta values.

Table 5. Demographic factors associated with cluster membership per timepoint.

Table 6. Dunn’s posthoc test for significant cluster differences in timepoint-wise group comparison

Table 7. Cross-tabulations for delivery mode at 2.5 months and preterm delivery at 6 months

Table 8. Metabolite concentration differences between clusters with Kruskall-Wallis test and Dunn’s Posthoc test. * Adjusted p-value from Dunn’s posthoc test ** adjusted p-value from Kruskall-Wallis test. Only results with q < 0.05 and Dunn’s test adjusted p-value < 0.05 are shown.

Table 9. Breastfeeding interaction with prevalent taxa rclr-transformed abundances in mixed model.

### Descriptives

Table 1. Sample characteristics.

| **timepoint** | 2.5 (N=444) | 6 (N=256) | 14 (N=302) | 30 (N=207) | Total (N=1209) |
| --- | --- | --- | --- | --- | --- |
| **antibiotics** |  |  |  |  |  |
| N-Miss | 140 | 124 | 169 | 111 | 544 |
| no | 302 (99.3%) | 128 (97.0%) | 111 (83.5%) | 75 (78.1%) | 616 (92.6%) |
| yes | 2 (0.7%) | 4 (3.0%) | 22 (16.5%) | 21 (21.9%) | 49 (7.4%) |
| **neonatal_antibiotics** |  |  |  |  |  |
| no | 395 (89.0%) | 227 (88.7%) | 266 (88.1%) | 182 (87.9%) | 1070 (88.5%) |
| yes | 49 (11.0%) | 29 (11.3%) | 36 (11.9%) | 25 (12.1%) | 139 (11.5%) |
| **sex** |  |  |  |  |  |
| boy | 229 (51.6%) | 143 (55.9%) | 163 (54.0%) | 121 (58.5%) | 656 (54.3%) |
| girl | 215 (48.4%) | 113 (44.1%) | 139 (46.0%) | 86 (41.5%) | 553 (45.7%) |
| **term** |  |  |  |  |  |
| preterm | 15 (3.4%) | 8 (3.1%) | 13 (4.3%) | 10 (4.8%) | 46 (3.8%) |
| term | 429 (96.6%) | 248 (96.9%) | 289 (95.7%) | 197 (95.2%) | 1163 (96.2%) |
| **pets** |  |  |  |  |  |
| N-Miss | 192 | 92 | 106 | 52 | 442 |
| no | 143 (56.7%) | 96 (58.5%) | 120 (61.2%) | 87 (56.1%) | 446 (58.1%) |
| yes | 109 (43.3%) | 68 (41.5%) | 76 (38.8%) | 68 (43.9%) | 321 (41.9%) |
| **delivery_mode** |  |  |  |  |  |
| N-Miss | 7 | 1 | 3 | 1 | 12 |
| section | 74 (16.9%) | 43 (16.9%) | 49 (16.4%) | 33 (16.0%) | 199 (16.6%) |
| vaginal | 363 (83.1%) | 212 (83.1%) | 250 (83.6%) | 173 (84.0%) | 998 (83.4%) |
| **perinatal_antibiotics** |  |  |  |  |  |
| N-Miss | 5 | 0 | 2 | 1 | 8 |
| no | 365 (83.1%) | 216 (84.4%) | 260 (86.7%) | 179 (86.9%) | 1020 (84.9%) |
| yes | 74 (16.9%) | 40 (15.6%) | 40 (13.3%) | 27 (13.1%) | 181 (15.1%) |
| **siblings** |  |  |  |  |  |
| N-Miss | 107 | 51 | 39 | 26 | 223 |
| no | 150 (44.5%) | 95 (46.3%) | 116 (44.1%) | 87 (48.1%) | 448 (45.4%) |
| yes | 187 (55.5%) | 110 (53.7%) | 147 (55.9%) | 94 (51.9%) | 538 (54.6%) |
| **current_breastfeeding** |  |  |  |  |  |
| N-Miss | 100 | 30 | 25 | 31 | 186 |
| no | 19 (5.5%) | 57 (25.2%) | 240 (86.6%) | 175 (99.4%) | 491 (48.0%) |
| yes | 325 (94.5%) | 169 (74.8%) | 37 (13.4%) | 1 (0.6%) | 532 (52.0%) |
| **breastfeeding_criteria** |  |  |  |  |  |
| N-Miss | 27 | 12 | 11 | 5 | 55 |
| adequate_breastfed | 276 (66.2%) | 173 (70.9%) | 215 (73.9%) | 137 (67.8%) | 801 (69.4%) |
| not_adequate | 141 (33.8%) | 71 (29.1%) | 76 (26.1%) | 65 (32.2%) | 353 (30.6%) |
| **sampling_age** |  |  |  |  |  |
| N-Miss | 2 | 3 | 0 | 2 | 7 |
| Mean (SD) | 65.213 (14.580) | 187.482 (20.510) | 428.768 (38.855) | 913.800 (62.809) | 327.017 (303.177) |
| Range | 12.000 - 125.000 | 47.000 - 227.000 | 47.000 - 782.000 | 47.000 - 1018.000 | 12.000 - 1018.000 |
| **mom_age** |  |  |  |  |  |
| Mean (SD) | 30.750 (4.376) | 30.723 (4.496) | 31.152 (4.308) | 31.155 (4.474) | 30.914 (4.401) |
| Range | 19.000 - 45.000 | 19.000 - 42.000 | 19.000 - 45.000 | 21.000 - 45.000 | 19.000 - 45.000 |
| **mom_edu** |  |  |  |  |  |
| N-Miss | 27 | 15 | 16 | 6 | 64 |
| High school | 109 (26.1%) | 64 (26.6%) | 71 (24.8%) | 48 (23.9%) | 292 (25.5%) |
| vocational | 135 (32.4%) | 72 (29.9%) | 90 (31.5%) | 65 (32.3%) | 362 (31.6%) |
| university | 173 (41.5%) | 105 (43.6%) | 125 (43.7%) | 88 (43.8%) | 491 (42.9%) |
| **Scfa data available** |  |  |  |  |  |
| N-Miss | 205 | 61 | 59 | 62 | 387 |
| yes | 239 (100.0%) | 195 (100.0%) | 243 (100.0%) | 145 (100.0%) | 822 (100.0%) |
| **Bile acid data available** |  |  |  |  |  |
| N-Miss | 207 | 62 | 59 | 62 | 390 |
| yes | 237 (100.0%) | 194 (100.0%) | 243 (100.0%) | 145 (100.0%) | 819 (100.0%) |
| **Polar metabolites data available** |  |  |  |  |  |
| N-Miss | 206 | 63 | 60 | 61 | 390 |
| yes | 238 (100.0%) | 193 (100.0%) | 242 (100.0%) | 146 (100.0%) | 819 (100.0%) |

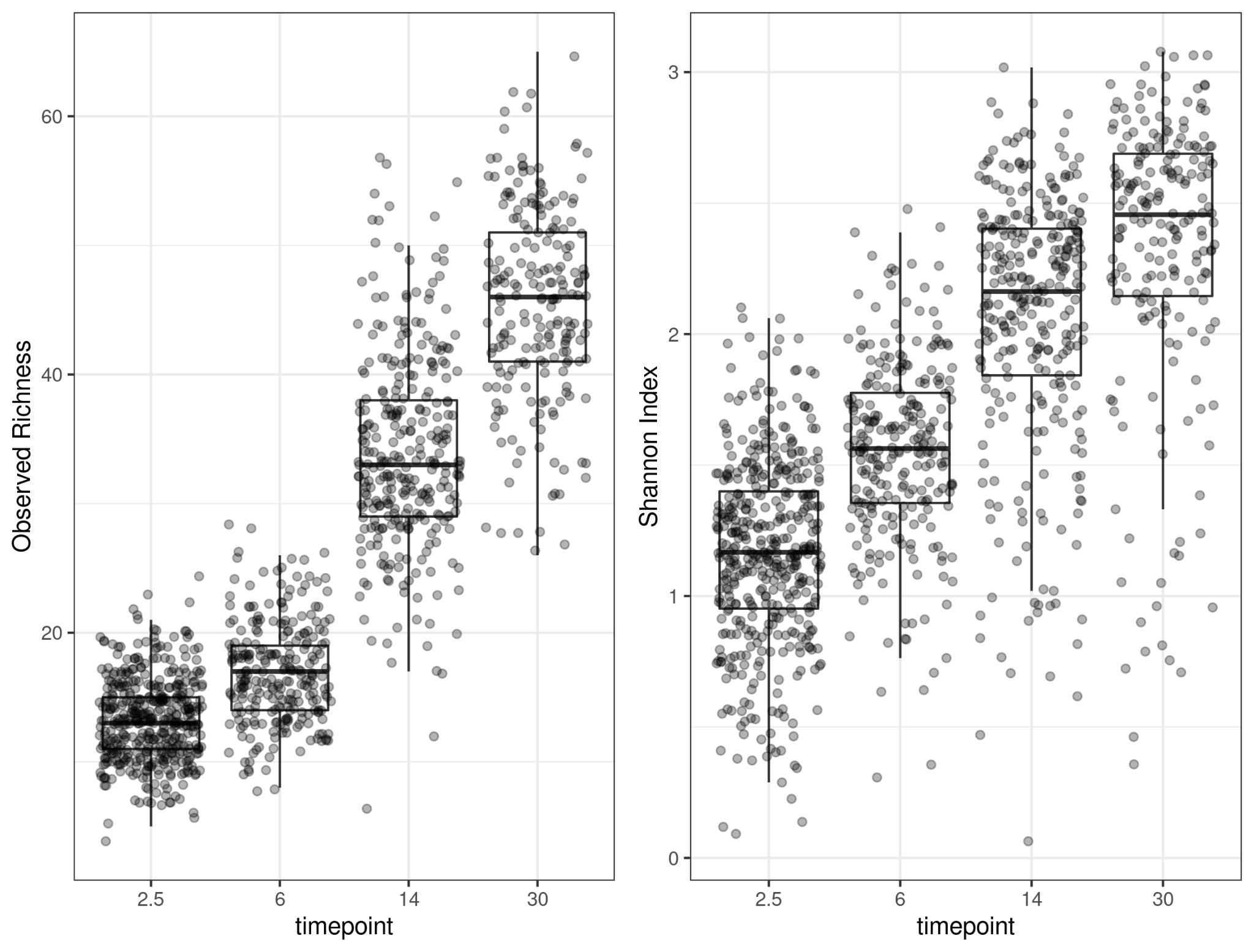

Figure 1. Richness and diversity in all timepoints

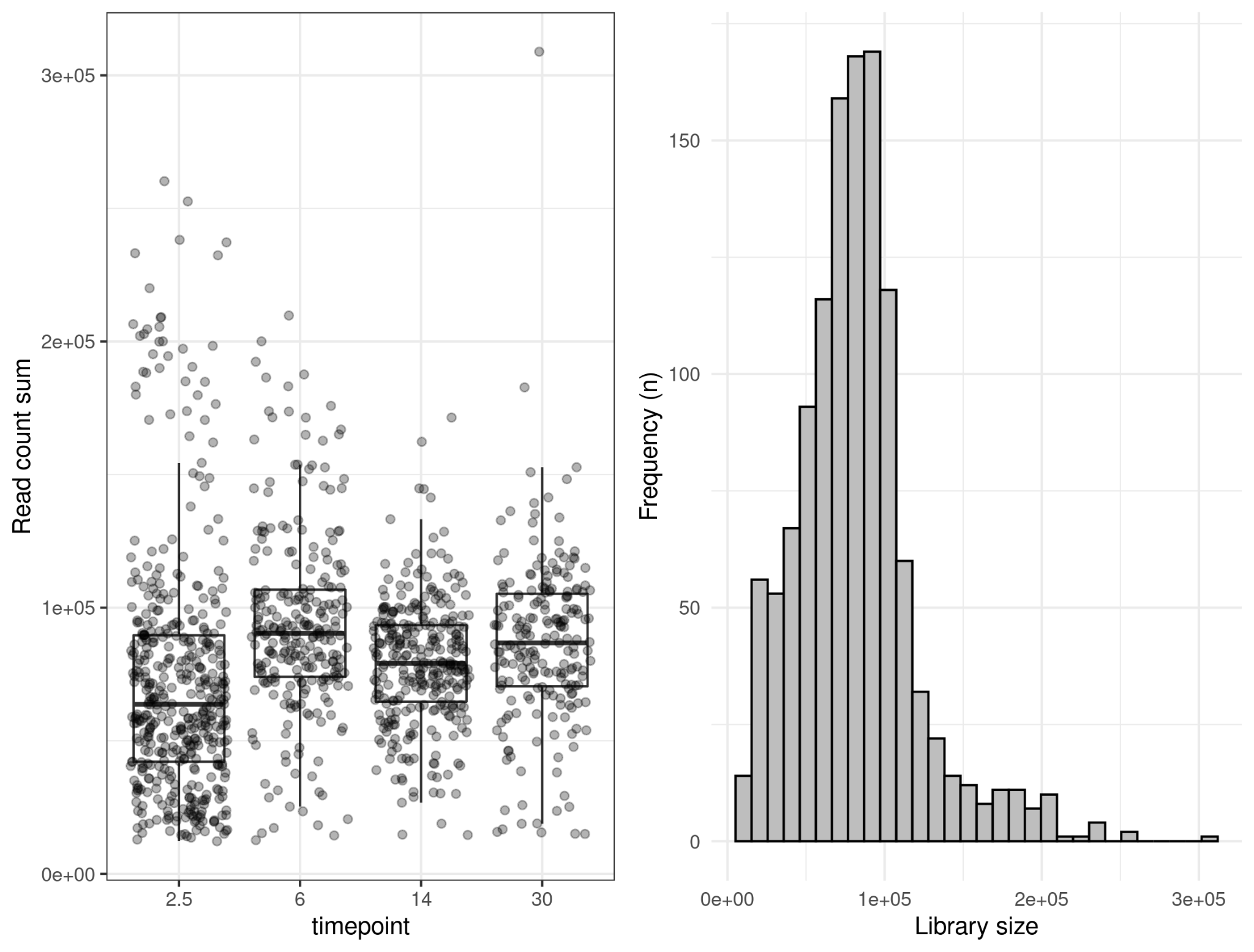

Figure 2. Library sizes in all timepoints and the whole sample.

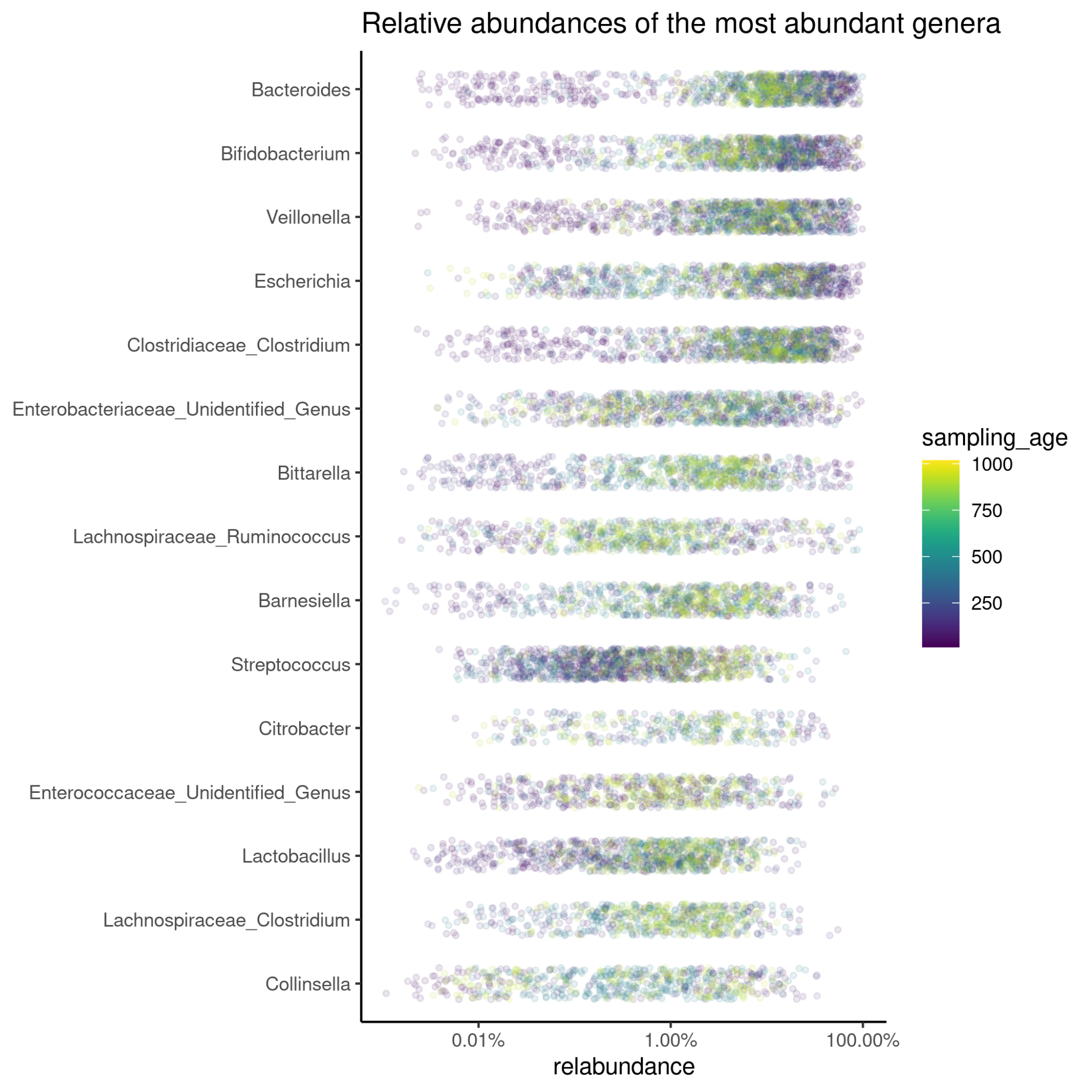

Figure 3. Relative abundances of the 15 most abundant genera.

Table 2. Genera prevalences. Genera with prevalence >= 1 % are presented.

| genus | prevalence |
| --- | --- |
| Bifidobacterium | 0.79 |
| Veillonella | 0.78 |
| Bacteroides | 0.78 |
| Clostridiaceae_Clostridium | 0.75 |
| Escherichia | 0.63 |
| Enterobacteriaceae_Unidentified_Genus | 0.45 |
| Bittarella | 0.4 |
| Barnesiella | 0.36 |
| Streptococcus | 0.3 |
| Lactobacillus | 0.29 |
| Lachnospiraceae_Clostridium | 0.24 |
| Enterococcaceae_Unidentified_Genus | 0.21 |
| Lachnospiraceae_Ruminococcus | 0.21 |
| Collinsella | 0.19 |
| Prevotella | 0.17 |
| Citrobacter | 0.17 |
| Parabacteroides | 0.13 |
| Lachnospiraceae_Unidentified_Genus | 0.12 |
| Hungatella | 0.12 |
| Flavonifractor | 0.11 |
| Sutterella | 0.11 |
| Haemophilus | 0.09 |
| Enterococcus | 0.08 |
| Roseburia | 0.08 |
| Alistipes | 0.08 |
| Blautia | 0.07 |
| Cellulosilyticum | 0.07 |
| Acidaminococcus | 0.06 |
| Faecalibacterium | 0.06 |
| Agathobacter | 0.05 |
| Staphylococcus | 0.05 |
| DTU089_Ruminococcus | 0.04 |
| Actinomyces | 0.03 |
| Lachnospiraceae_Eubacterium | 0.02 |
| Ruthenibacterium | 0.02 |
| Eggerthella | 0.02 |
| Clostridioides | 0.02 |
| Erysipelatoclostridium | 0.02 |
| 992a | 0.02 |
| Intestinibacter | 0.01 |
| Romboutsia | 0.01 |
| CAG-41 | 0.01 |
| Dorea | 0.01 |
| Desulfovibrionaceae_Unidentified_Genus | 0.01 |
| CAG-81 | 0.01 |
| GCA-900066995 | 0.01 |
| Ruminococcaceae_Unidentified_Genus | 0.01 |
| Coprococcus | 0.01 |
| Anaeroglobus | 0.01 |
| Dialister | 0.01 |
| Fusicatenibacter | 0.01 |
| Actinomycetaceae_Unidentified_Genus | 0.01 |
| Akkermansia | 0.01 |
| Erysipelotrichaceae_Eubacterium | 0.01 |

### Control samples

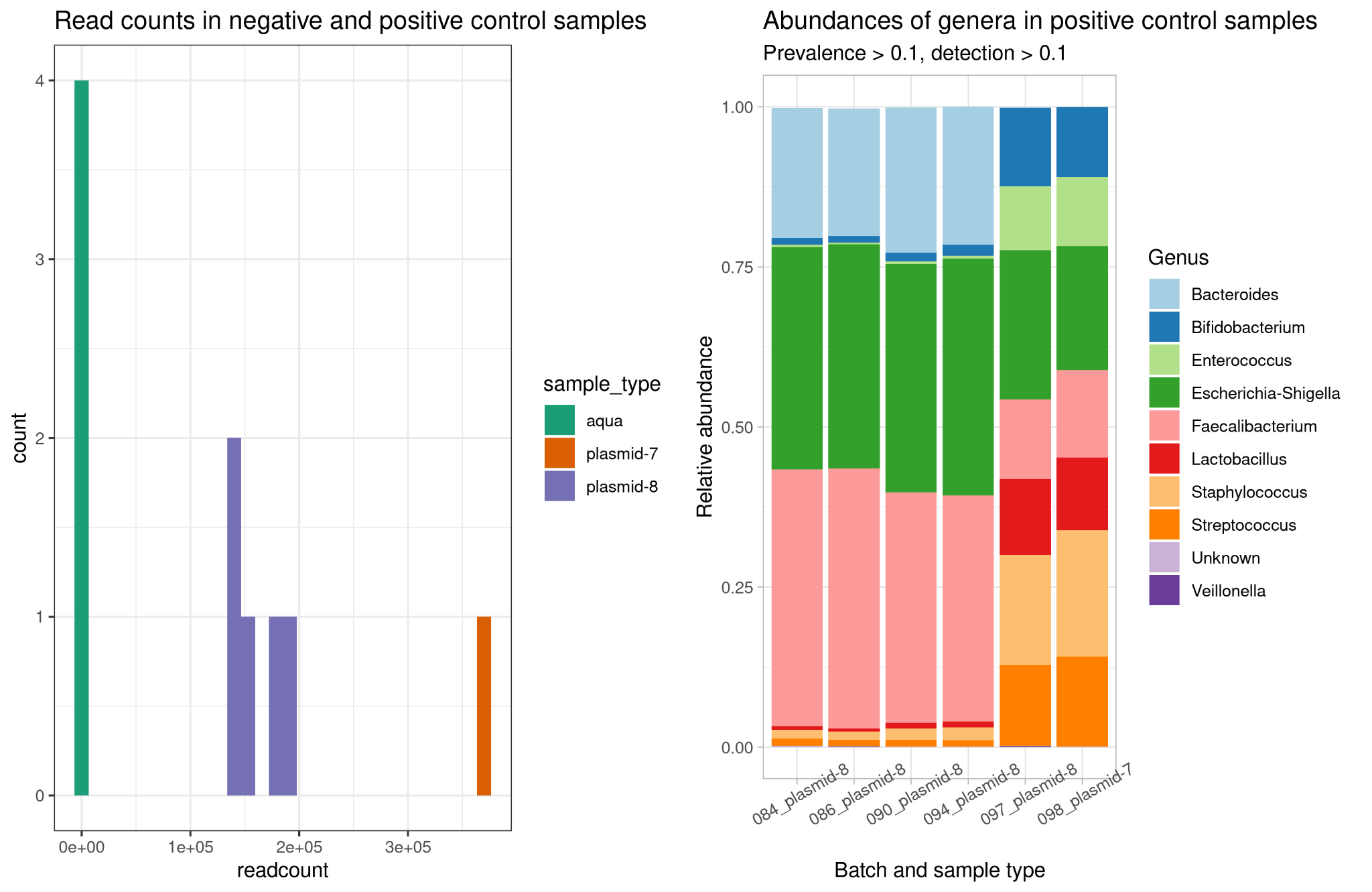

Figure 4. Positive and negative control samples in 2.5 monts time point. A. read counts across control samples. B. Relative abundances of core genera in positive control samples.

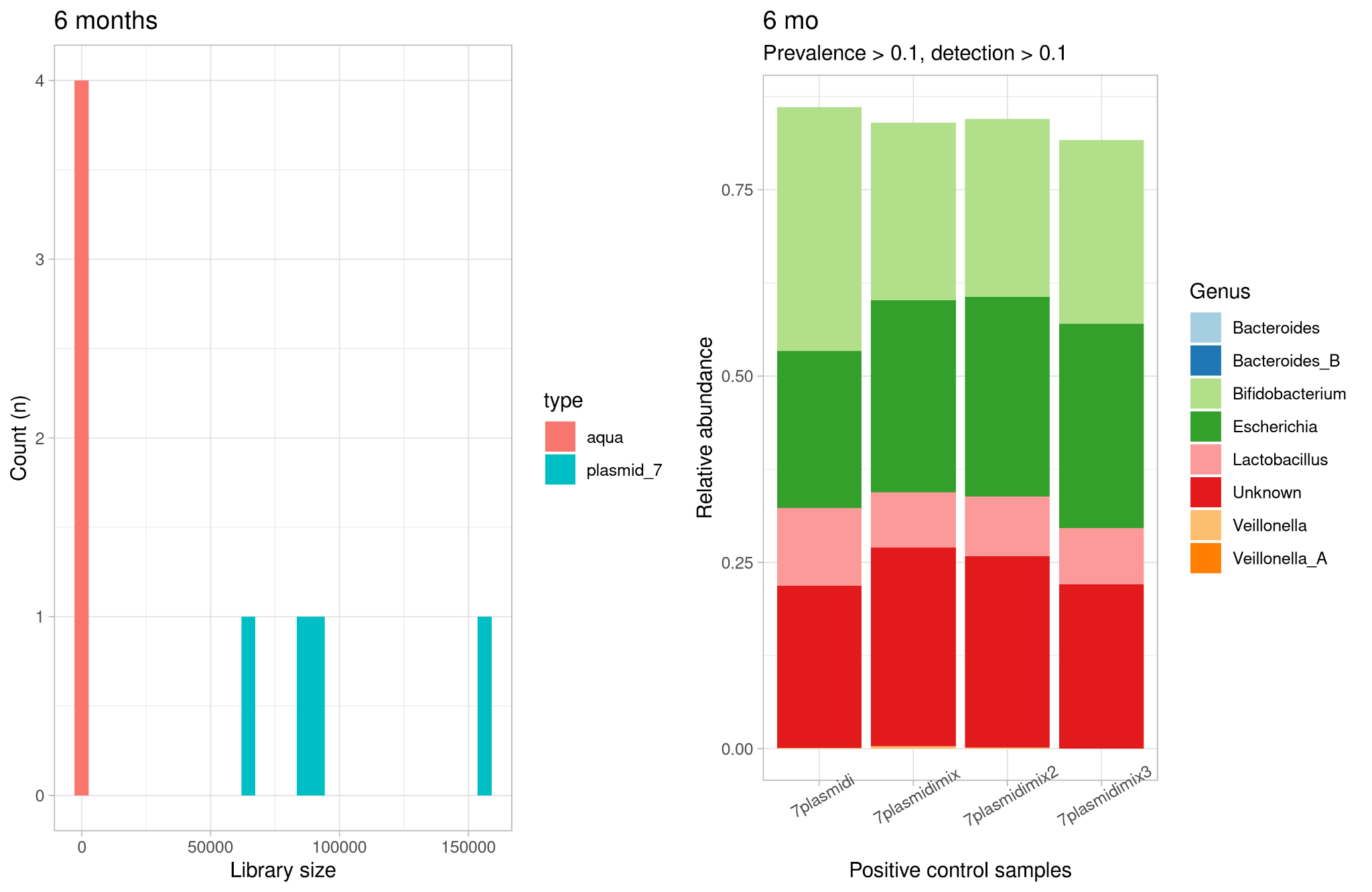

Figure 5. Positive and negative control samples in 6 months time point. A. read counts across control samples. B. Relative abundances of core genera in positive control samples.

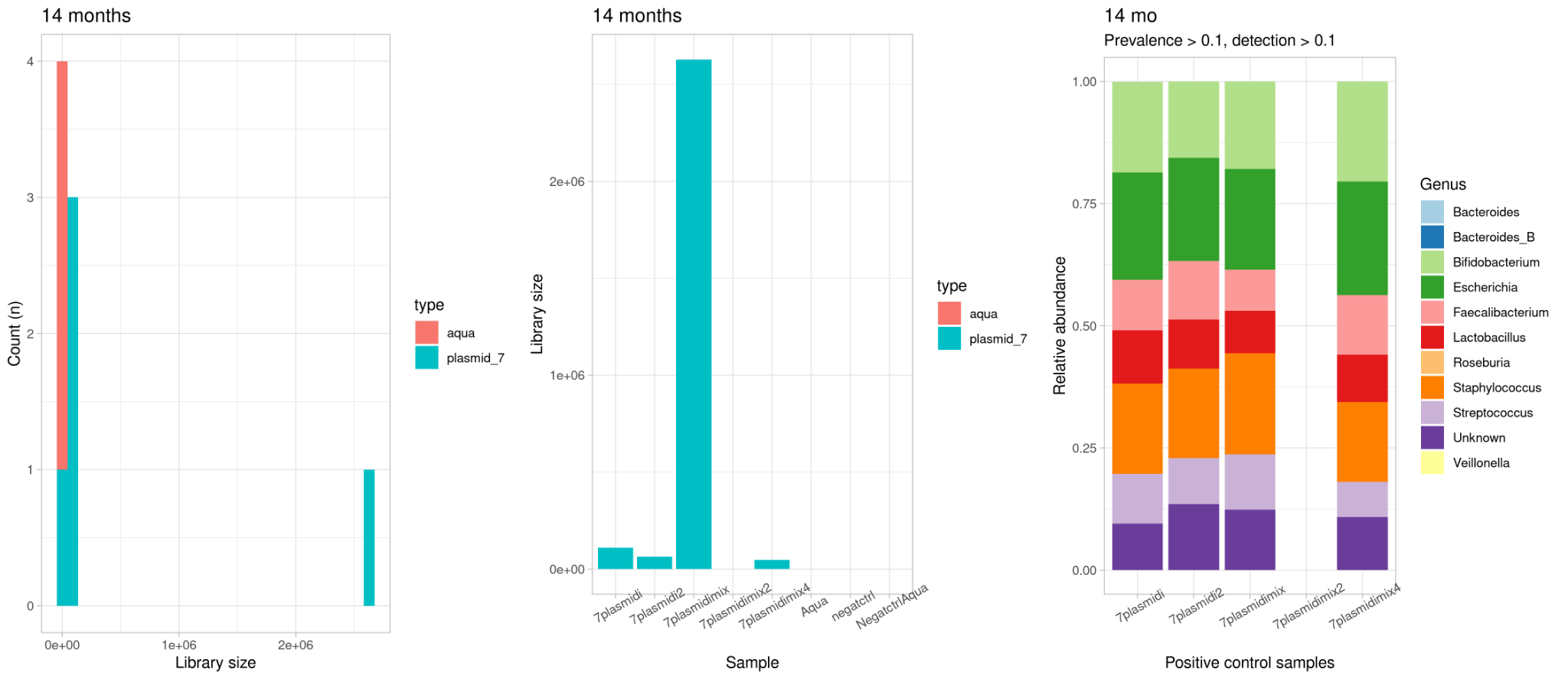

Figure 6. Positive and negative control samples in 14 months time point. A. read counts across control samples per control sample type. B. Read counts in all individual control samples. C. Relative abundances of core genera in positive control samples.

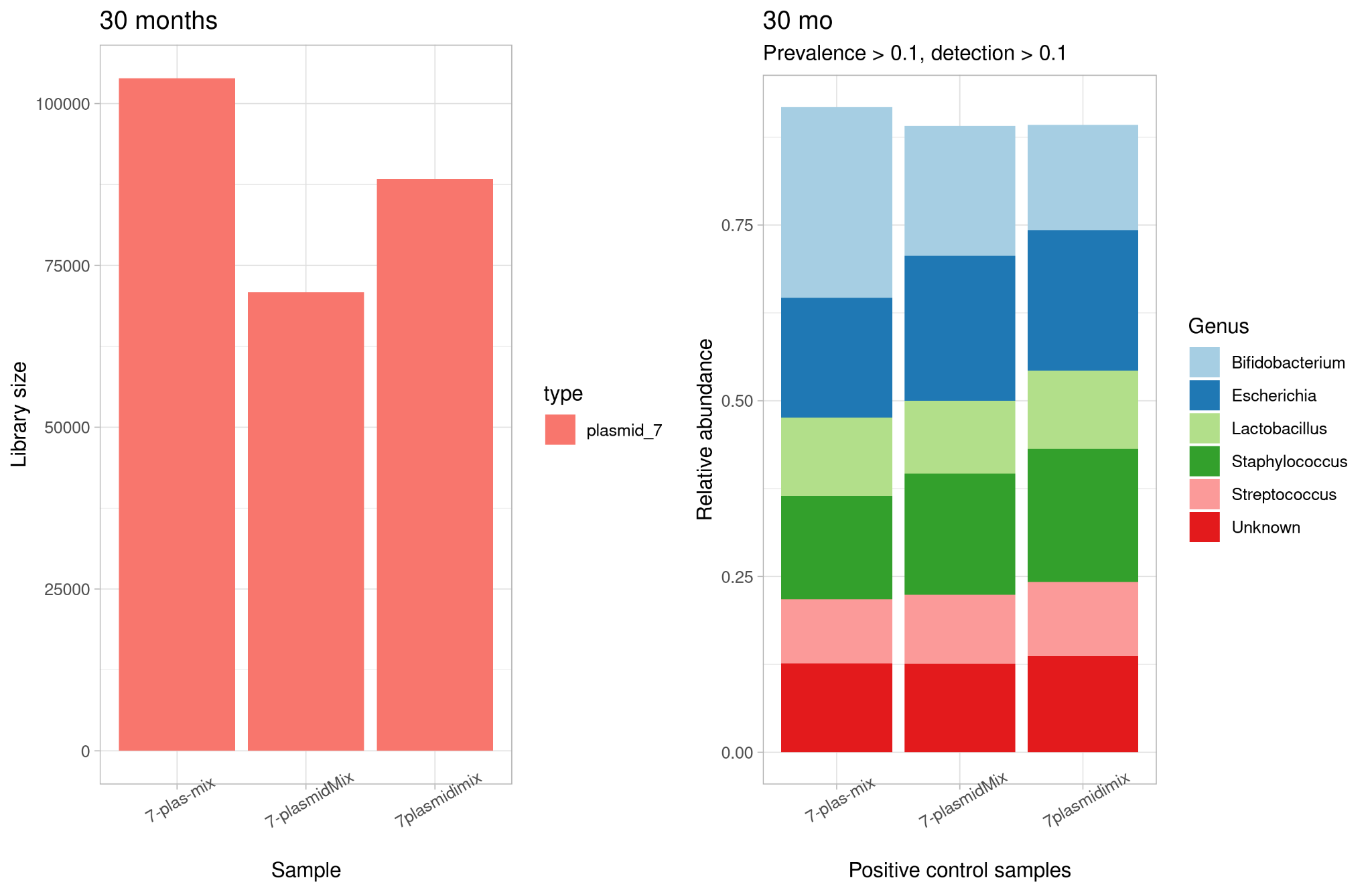

Figure 7. Positive and negative control samples in 6 months time point. A. read counts across control samples. B. Relative abundances of core genera in positive control samples.

### Metabolites and age

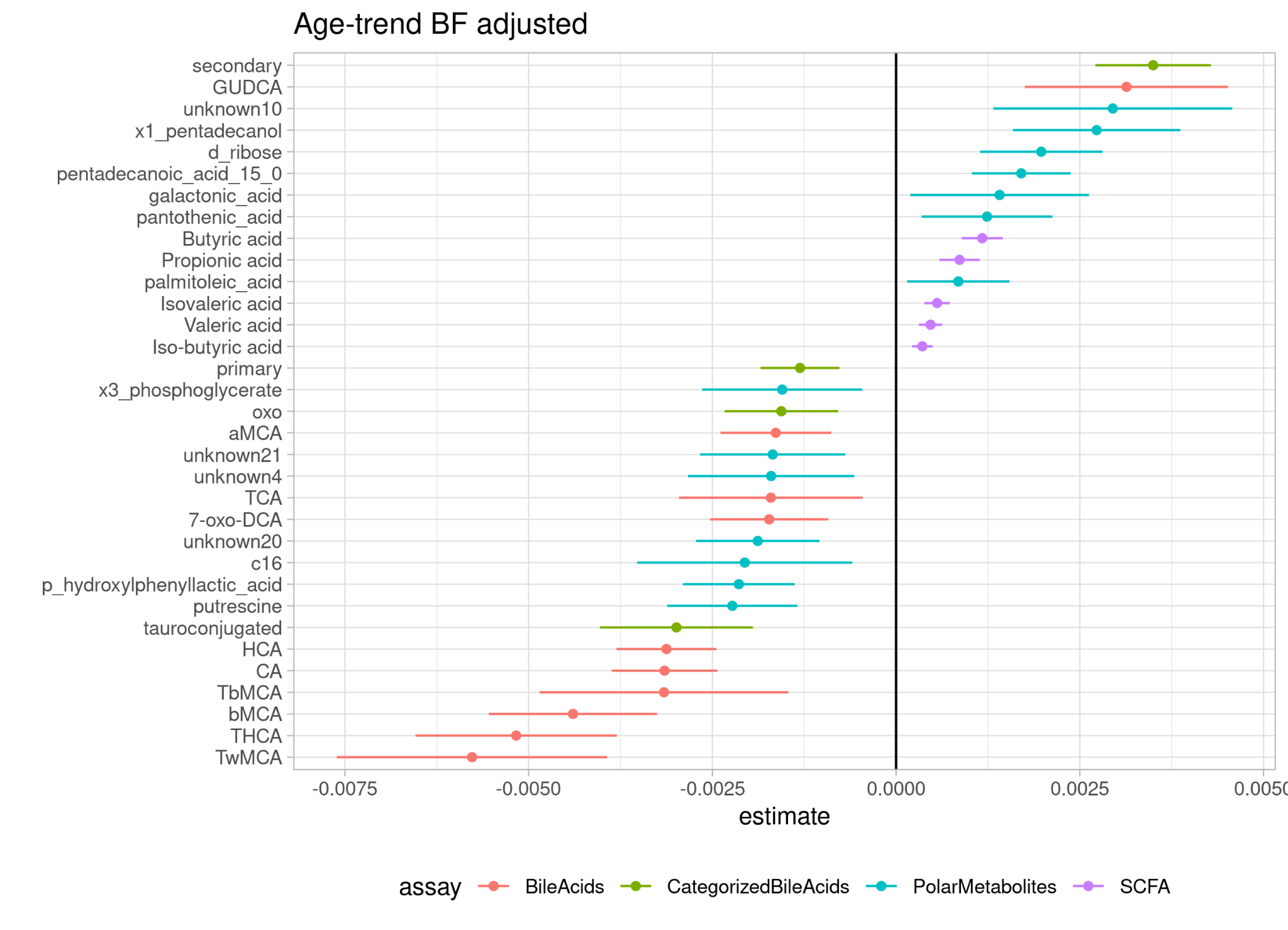

Figure 8. Estimates and 95 % confidence intervals for metabolites associated with age when adjusting for current breastfeeding.

### Metabolites and demographics

Table 3. Variance explained on average by the demographic factors.

| assay | antibiotics | neonatal_antibiotics | sex | term | pets | delivery_mode | perinatal_antibiotics | siblings | current_breastfeeding | breastfeeding_criteria |
| --- | --- | --- | --- | --- | --- | --- | --- | --- | --- | --- |
| SCFA | 0.1 | 0.06 | 0.08 | 0.13 | 0.74 | 0.04 | 0.1 | 0.09 | 0.1 | 0.18 |
| BileAcids | 0.53 | 0.13 | 0.28 | 0.13 | 0.09 | 0.21 | 0.09 | 0.16 | 0.22 | 0.17 |
| PolarMetabolites | 0.32 | 0.13 | 0.09 | 0.1 | 0.29 | 0.14 | 0.13 | 0.14 | 0.98 | 0.16 |

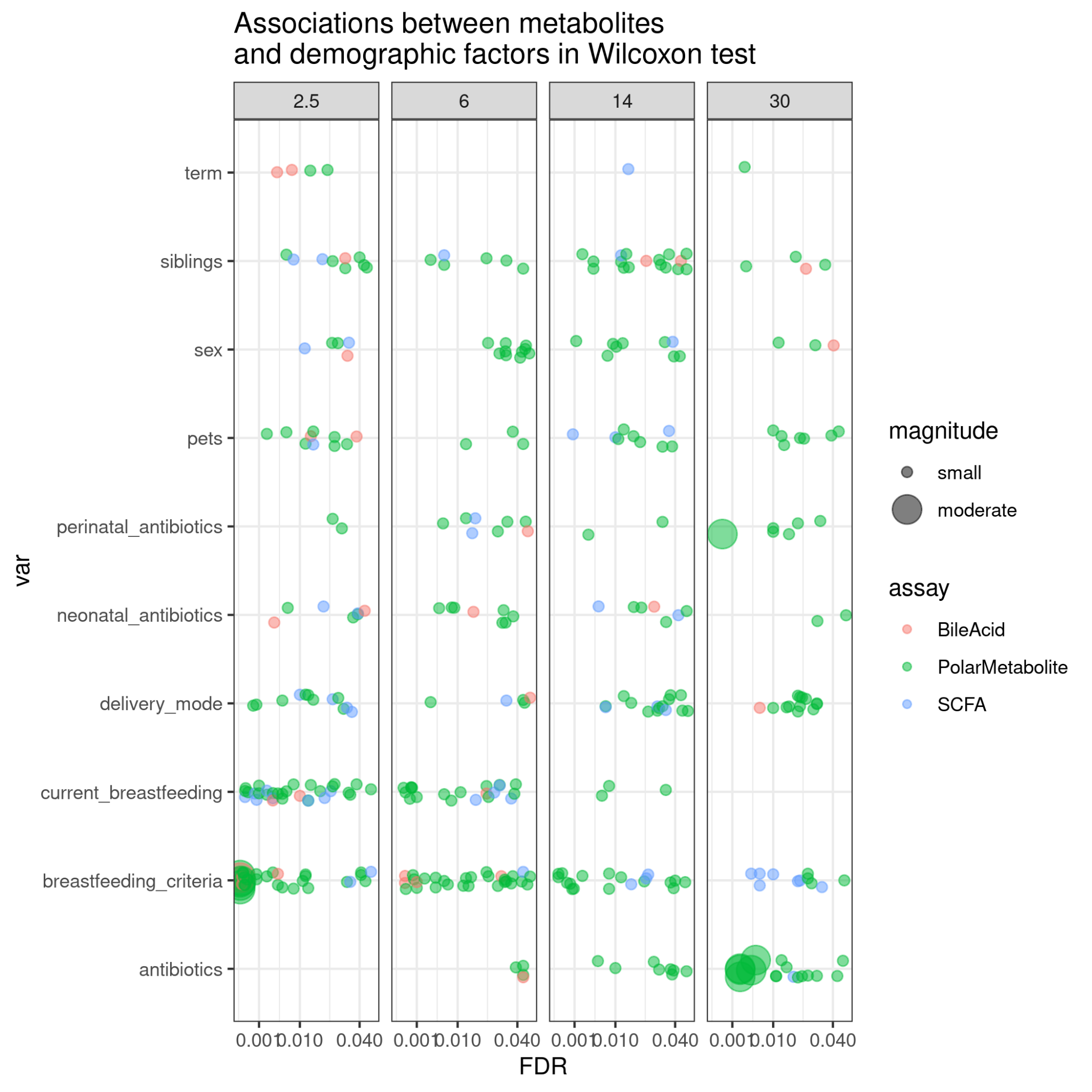

Figure 9. Associations between demographic factors and metabolite concentrations per timepoint. Effect size r is calculated as Z statistic divided by square root of the sample size, where r < 0.3 is considered small, r 0.3-<0.5 moderate and ⩾ 0.5 large. Only findings with FDR < 0.05 and moderate to large effect size are visualized.

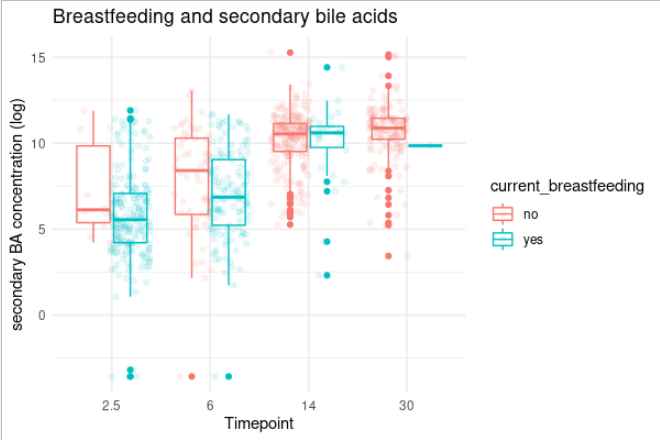

Figure 10. Secondary bile acid concentrations in breastfed and non-breastfed children per timepoint.

#### Metabolites and demographic factors

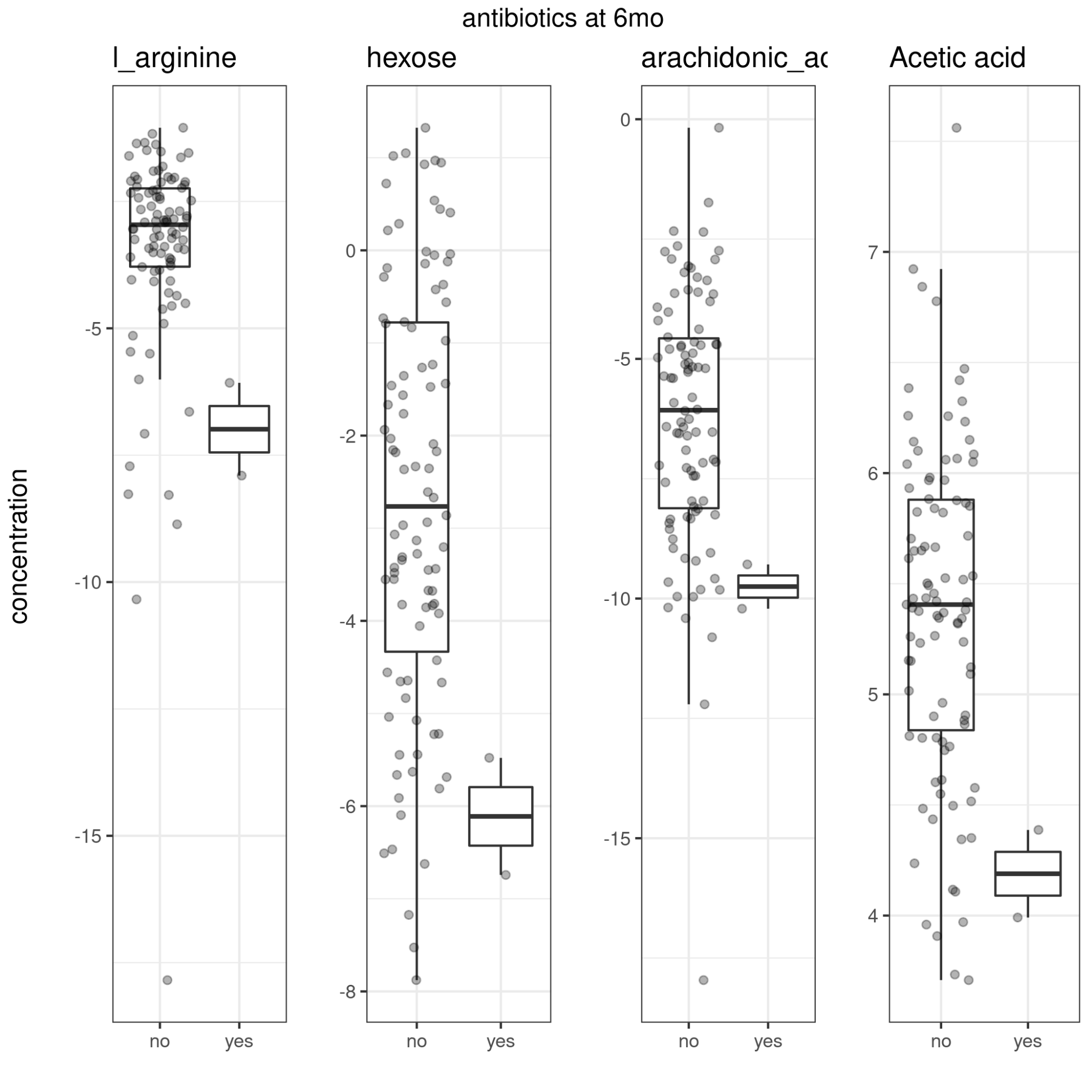

Figure 11. Antibiotic use and metabolite concentrations at 6 mo.

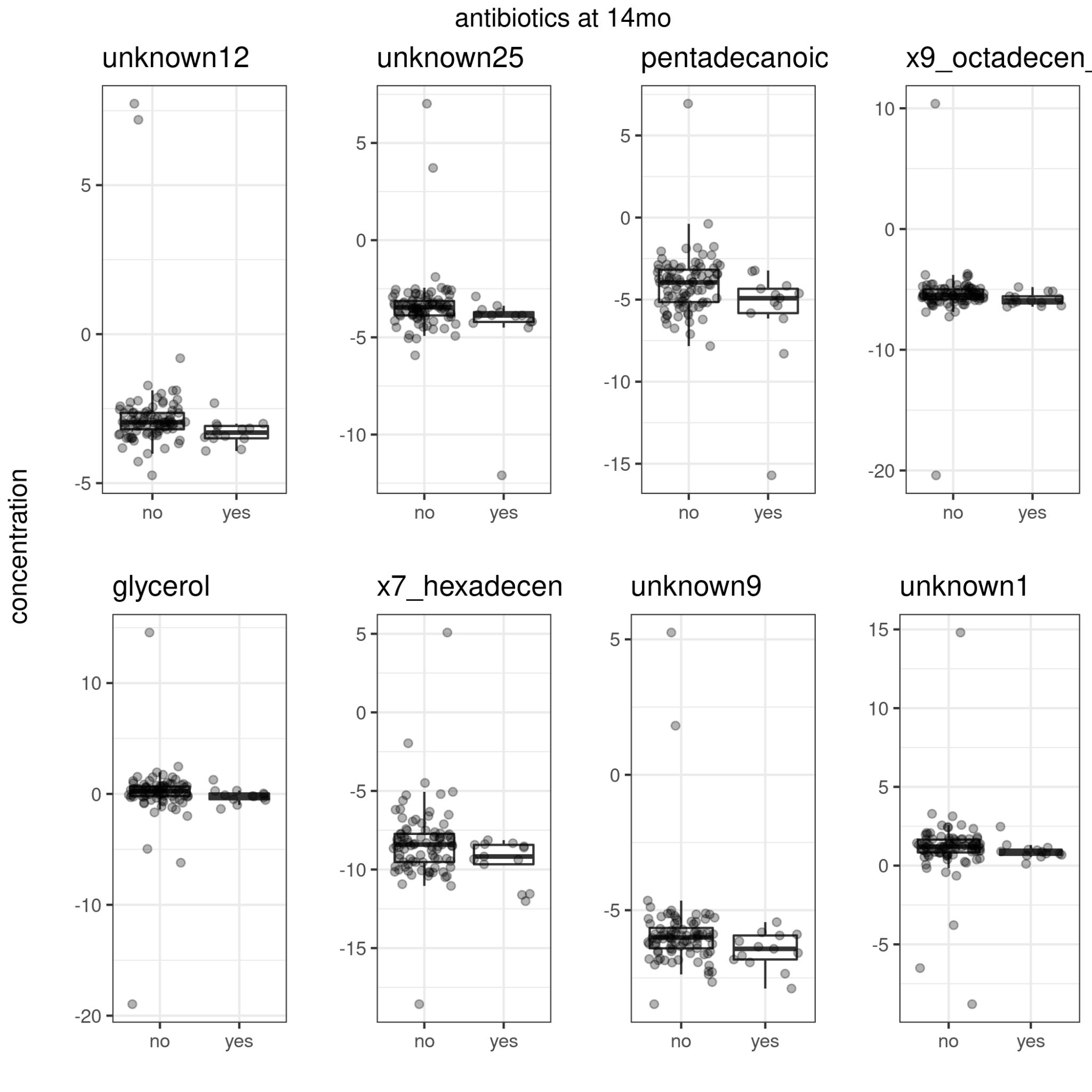

Figure 12. Antibiotic use and metabolite concentrations at 14 mo.

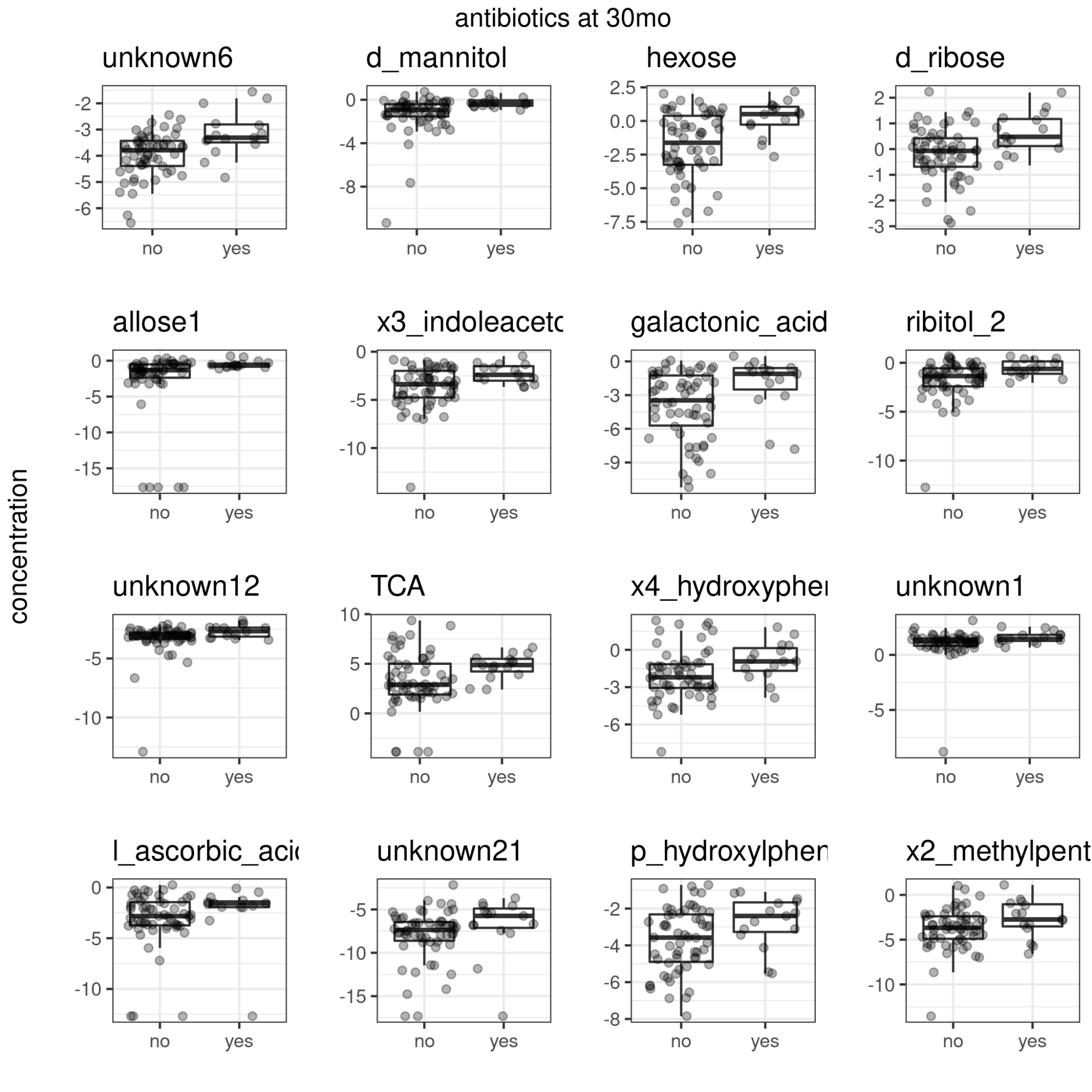

Figure 13. Antibiotic use and metabolite concentrations at 30 mo.

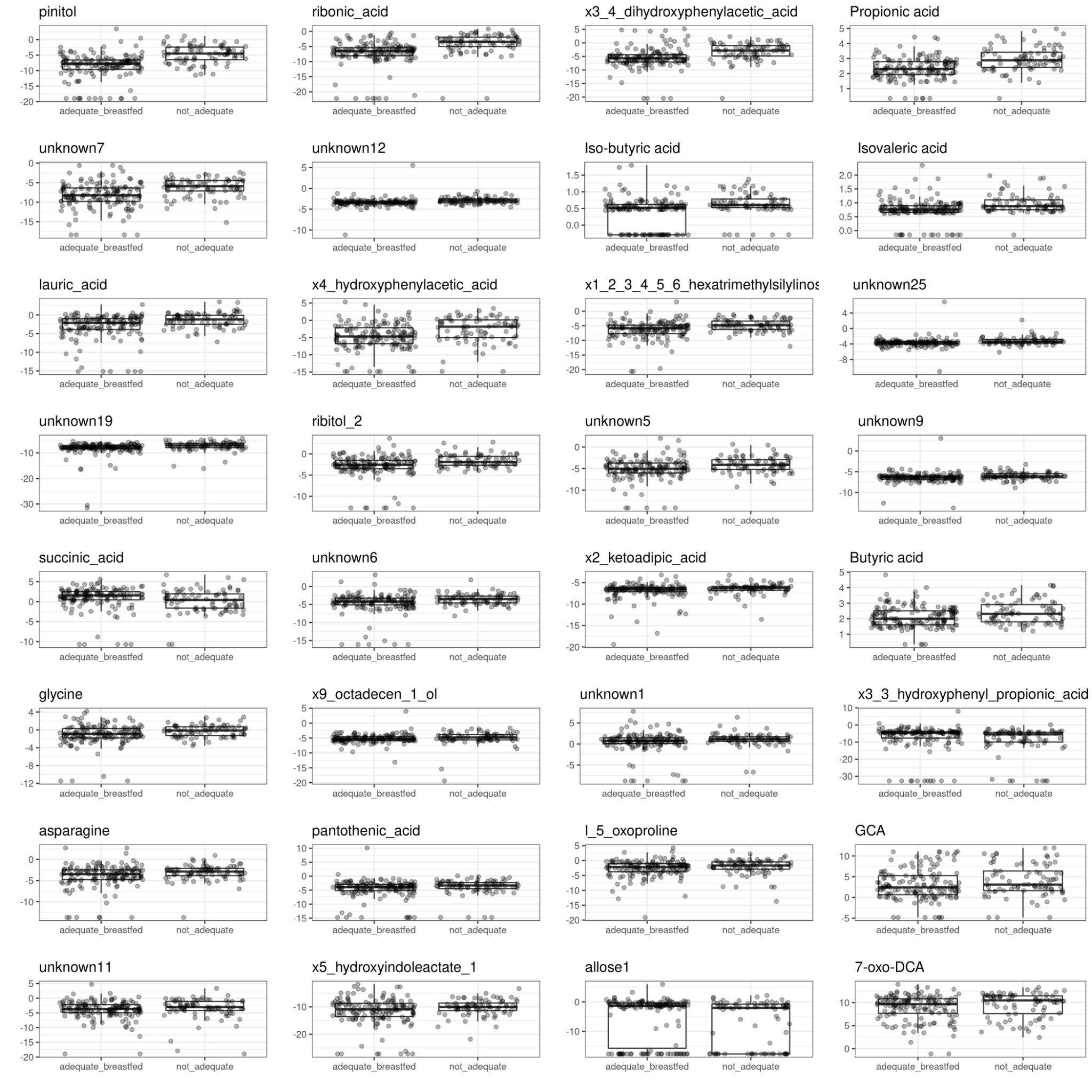

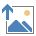

Figure 14. Adequate vs non-adequate breastfeeding and metabolite concentrations at 2.5 mo.

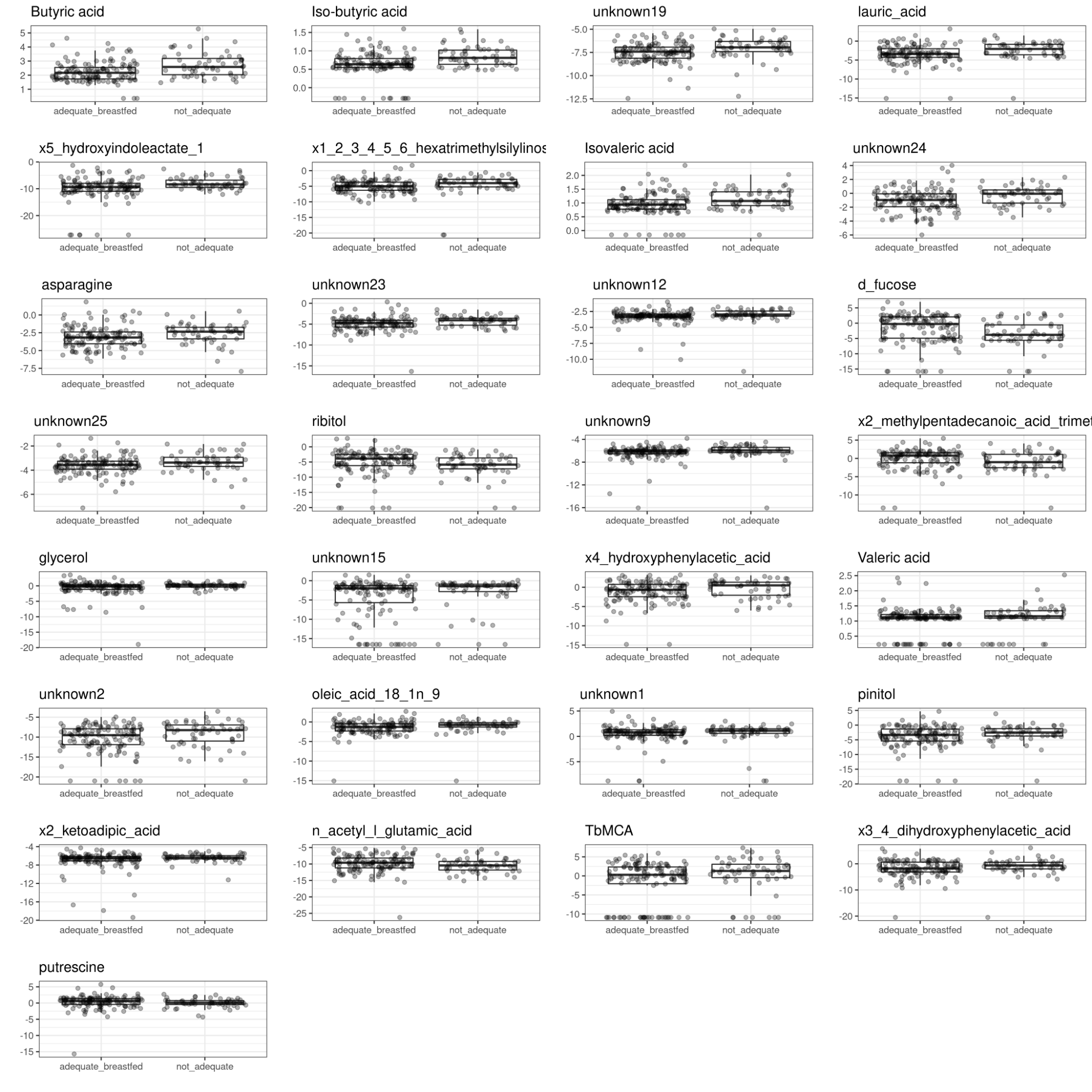

Figure 15. Adequate vs non-adequate breastfeeding and metabolite concentrations at 6 mo.

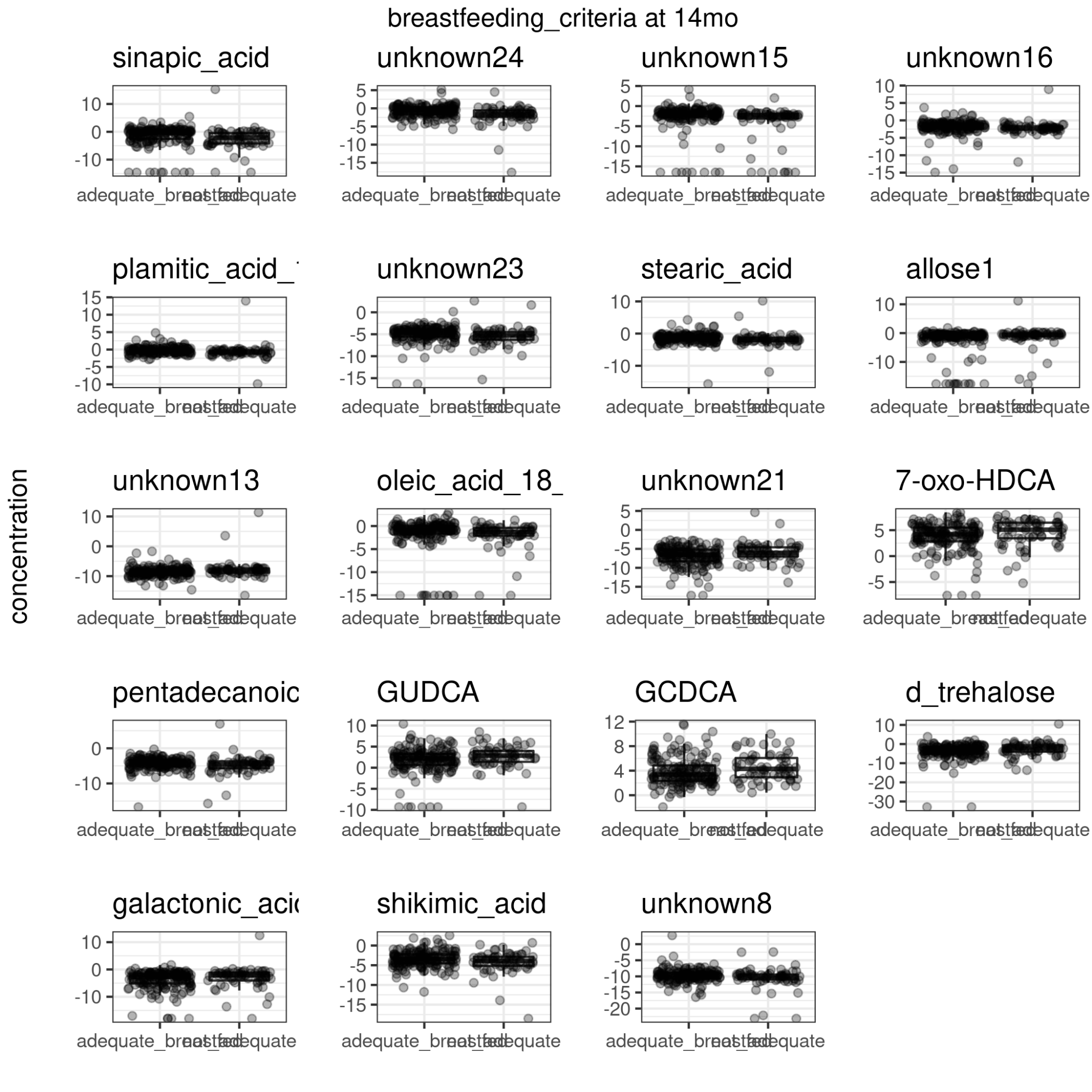

Figure 16. Adequate vs non-adequate breastfeeding and metabolite concentrations at 14 mo.

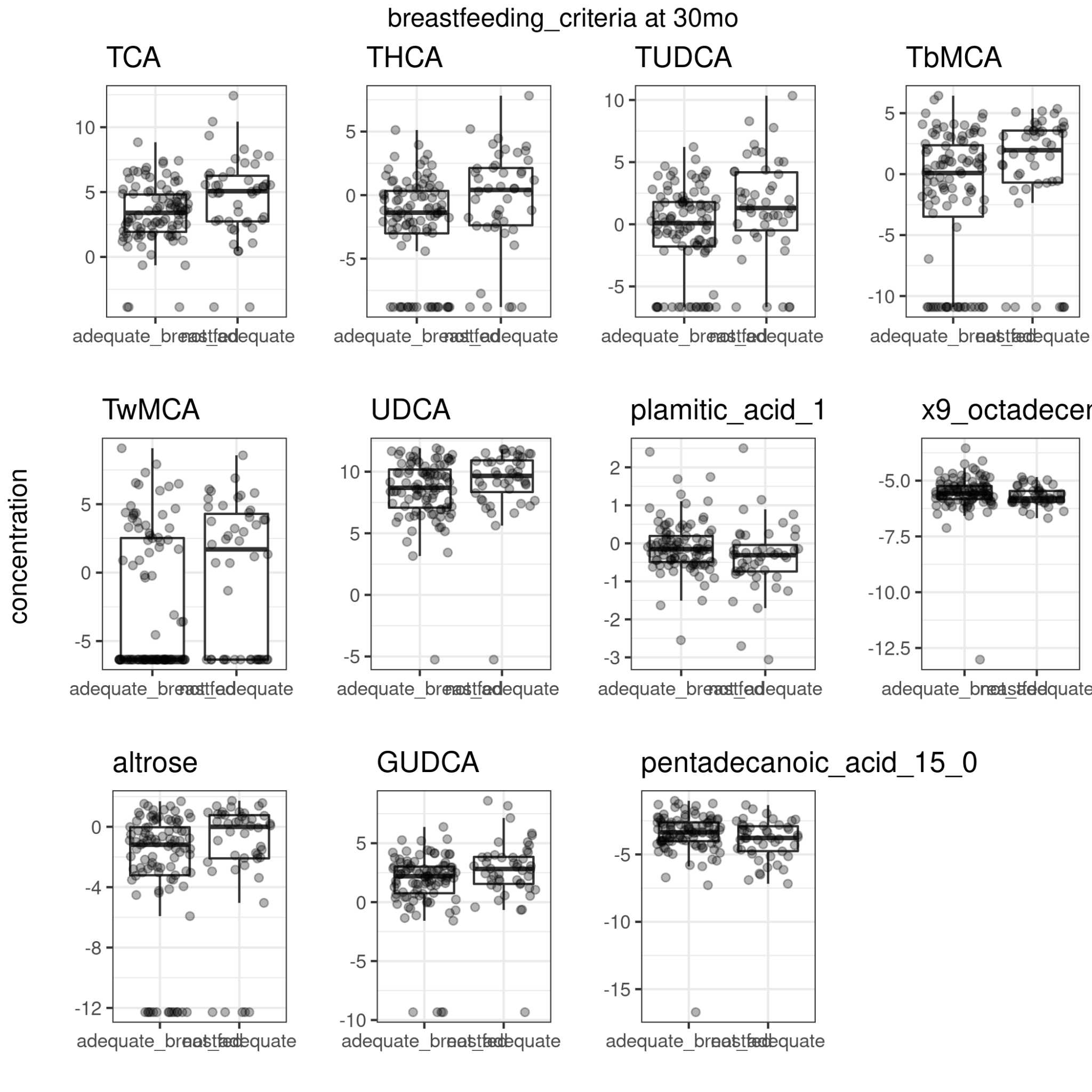

Figure 17. Adequate vs non-adequate breastfeeding and metabolite concentrations at 30 mo.

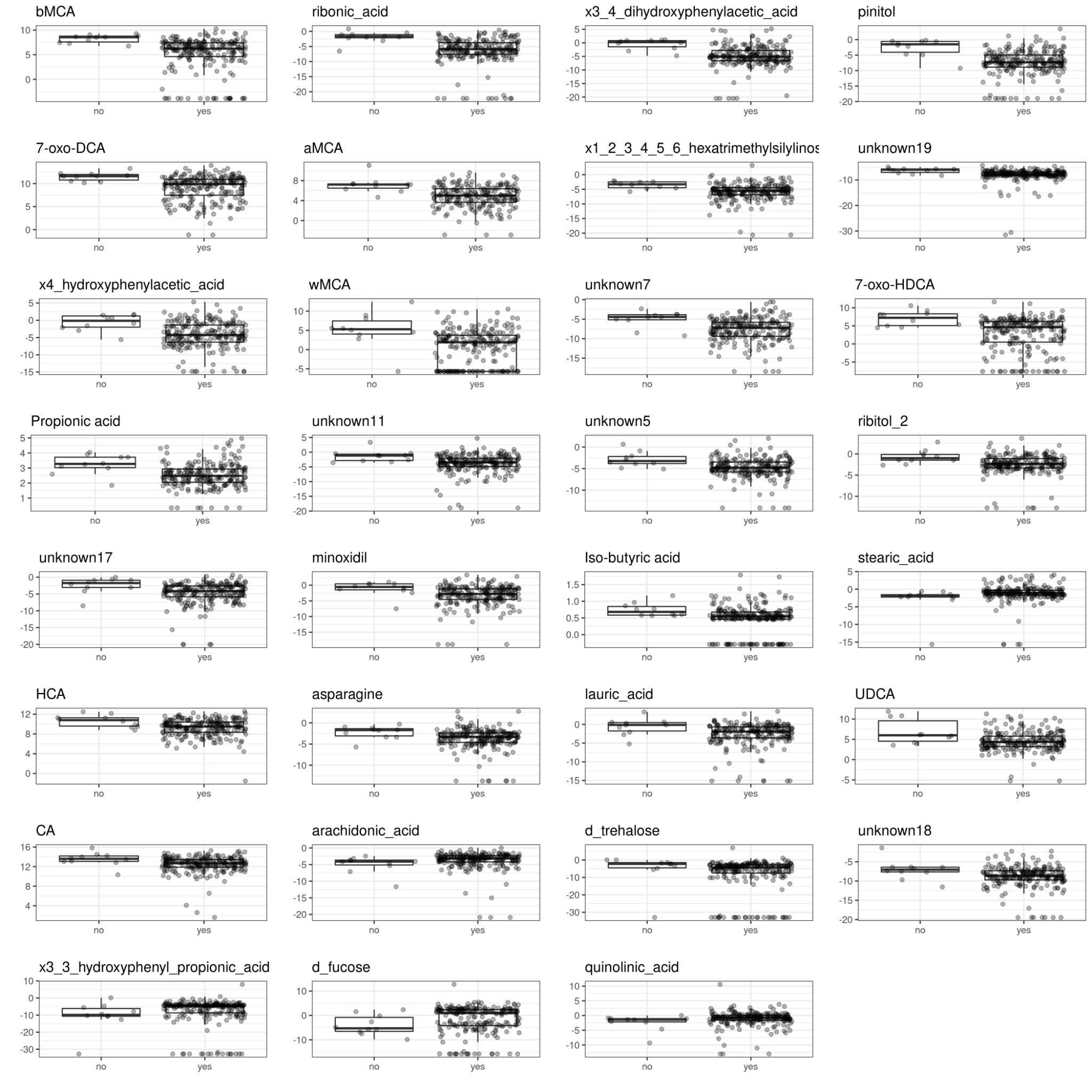

Figure 18. Current breastfeeding and metabolite concentrations at 2.5 mo.

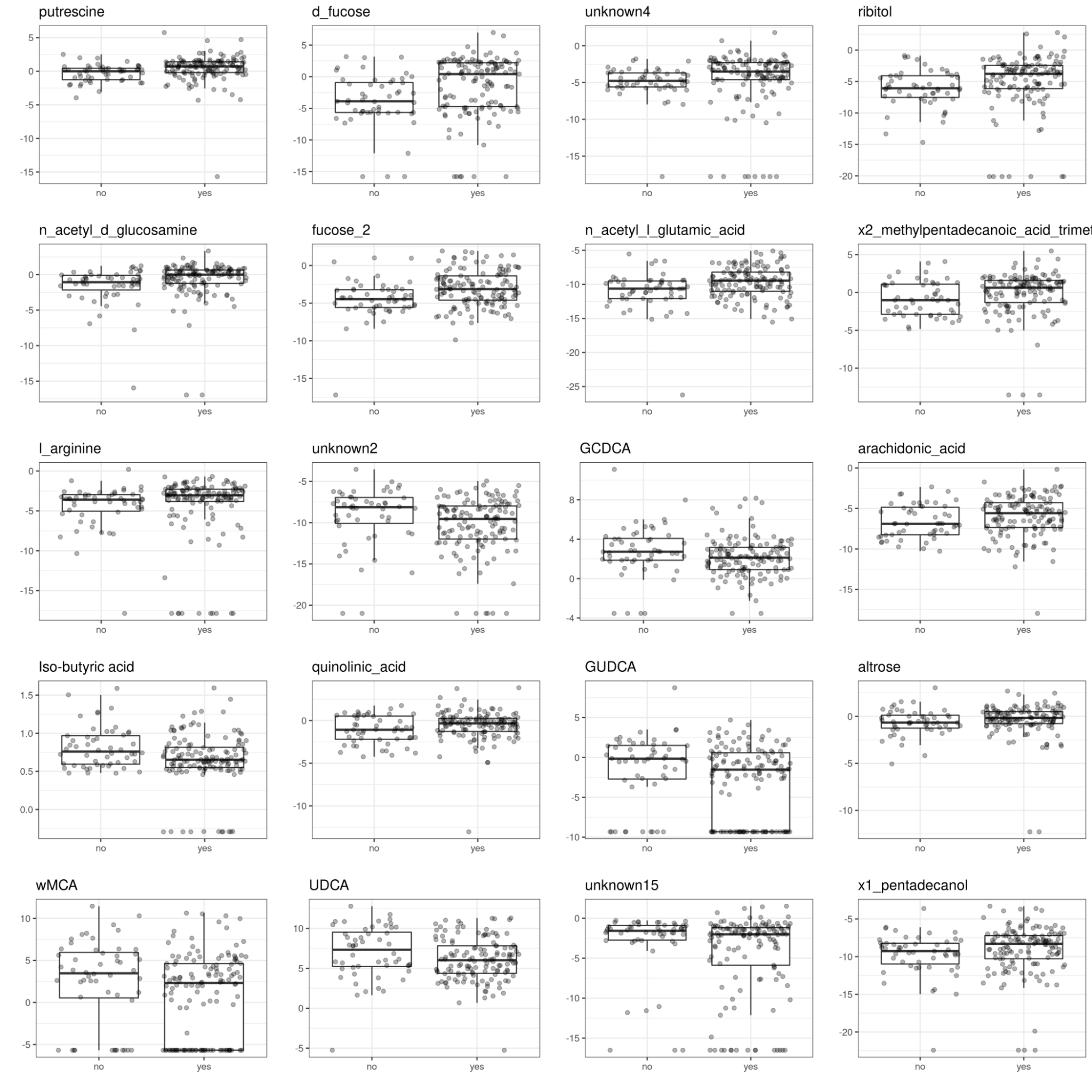

Figure 19. Current breastfeeding and metabolite concentrations at 6 mo.

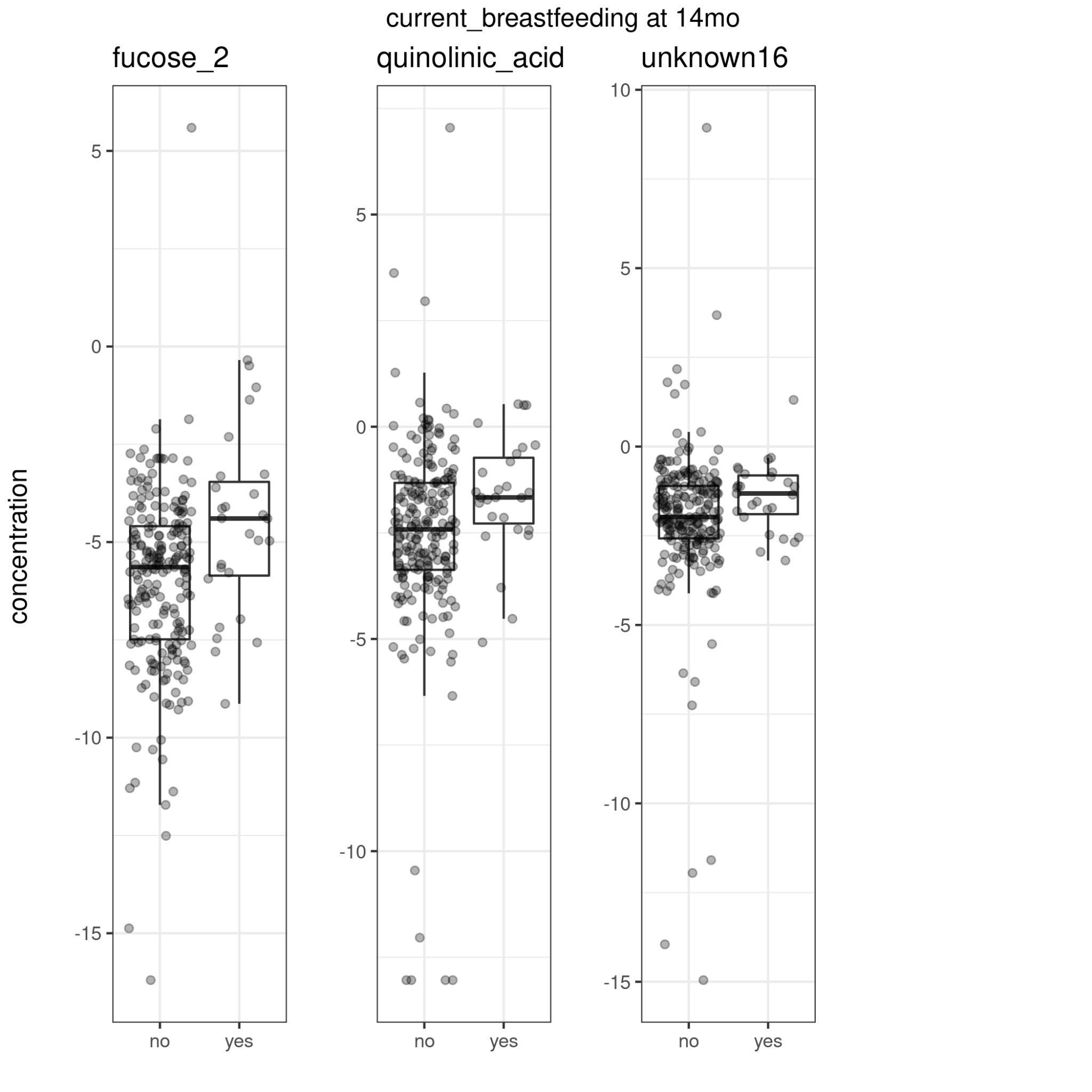

Figure 20. Current breastfeeding and metabolite concentrations at 14 mo.

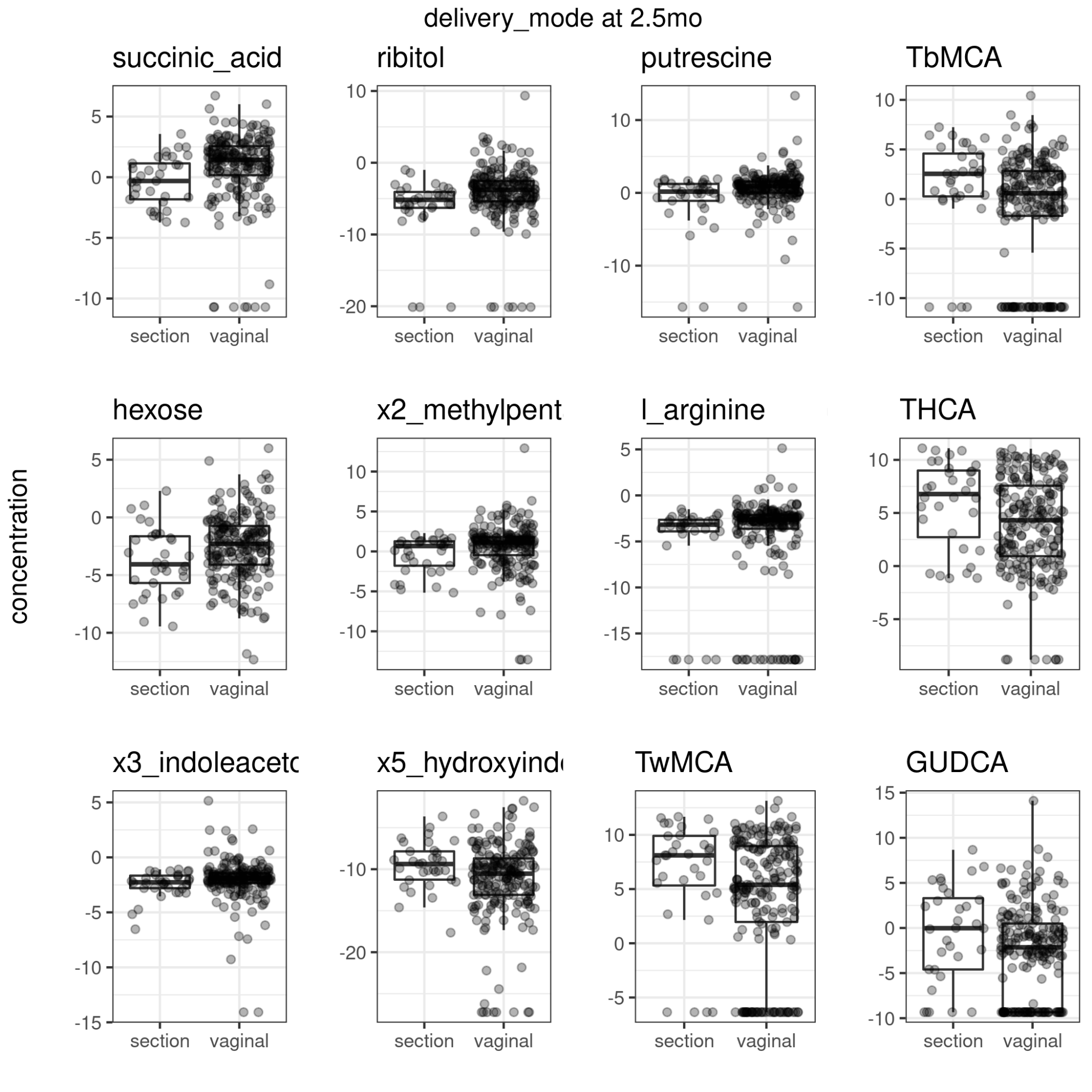

Figure 21. Delivery mode and metabolite concentrations at 2.5 mo.

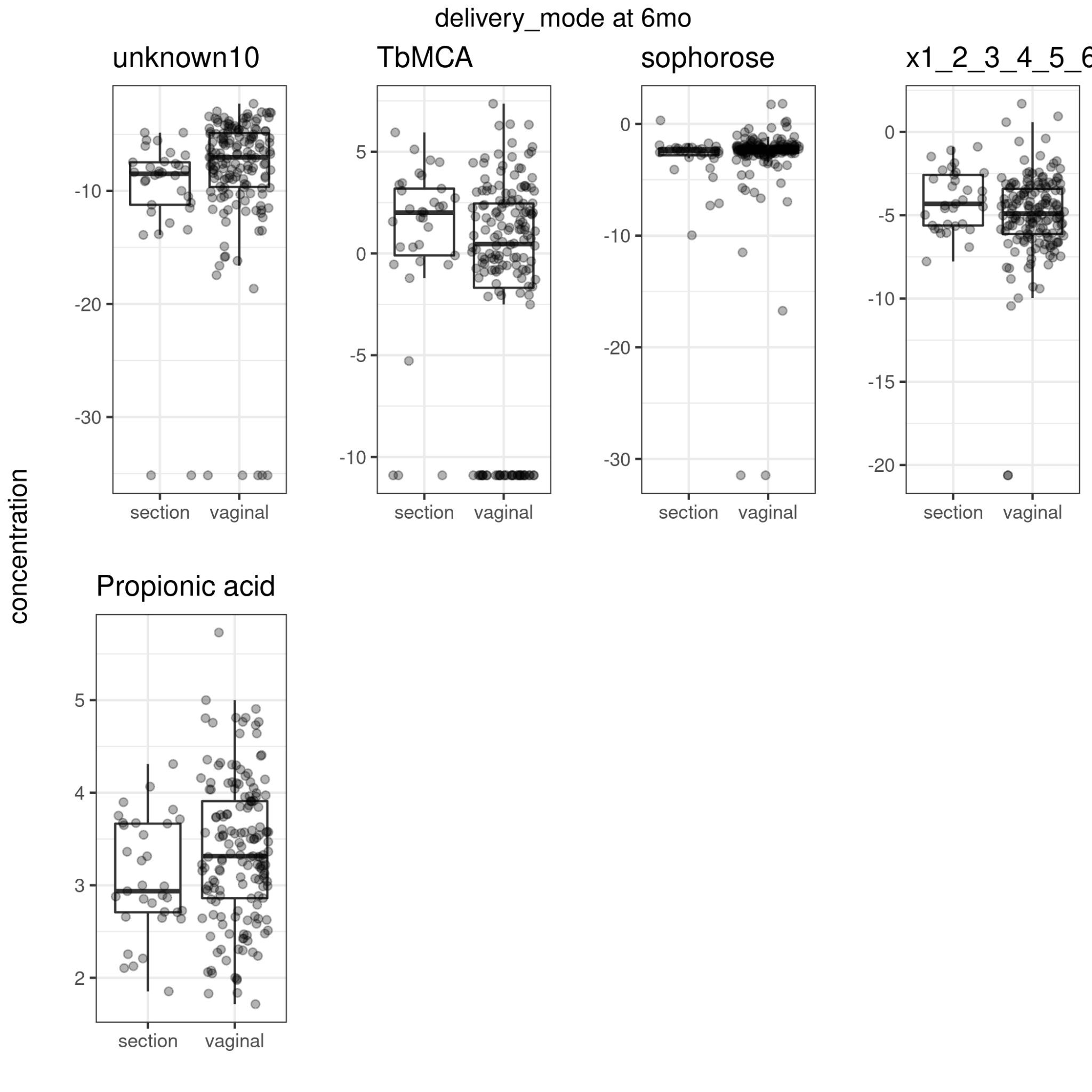

Figure 22. Delivery mode and metabolite concentrations at 6 mo.

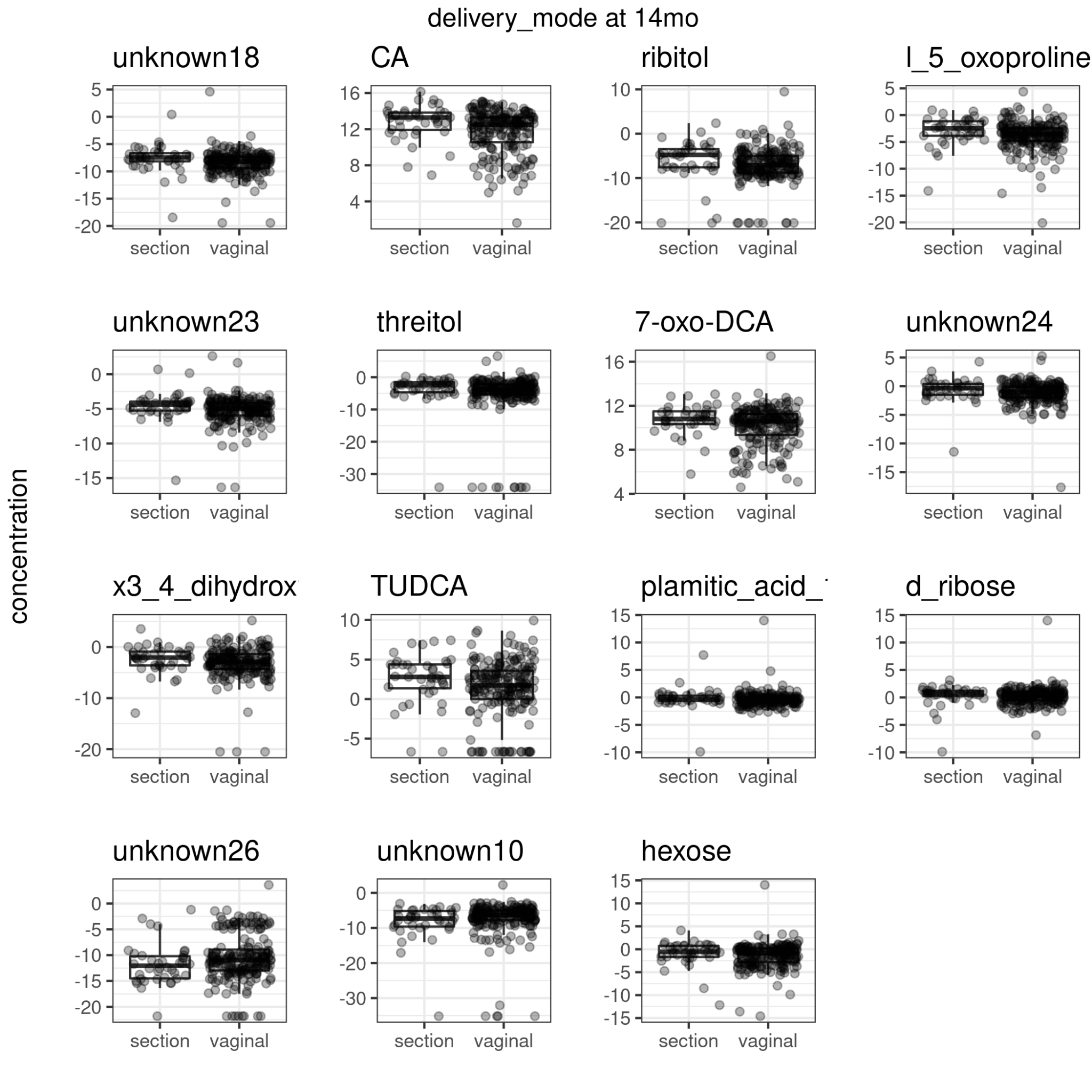

Figure 23. Delivery mode and metabolite concentrations at 14 mo.

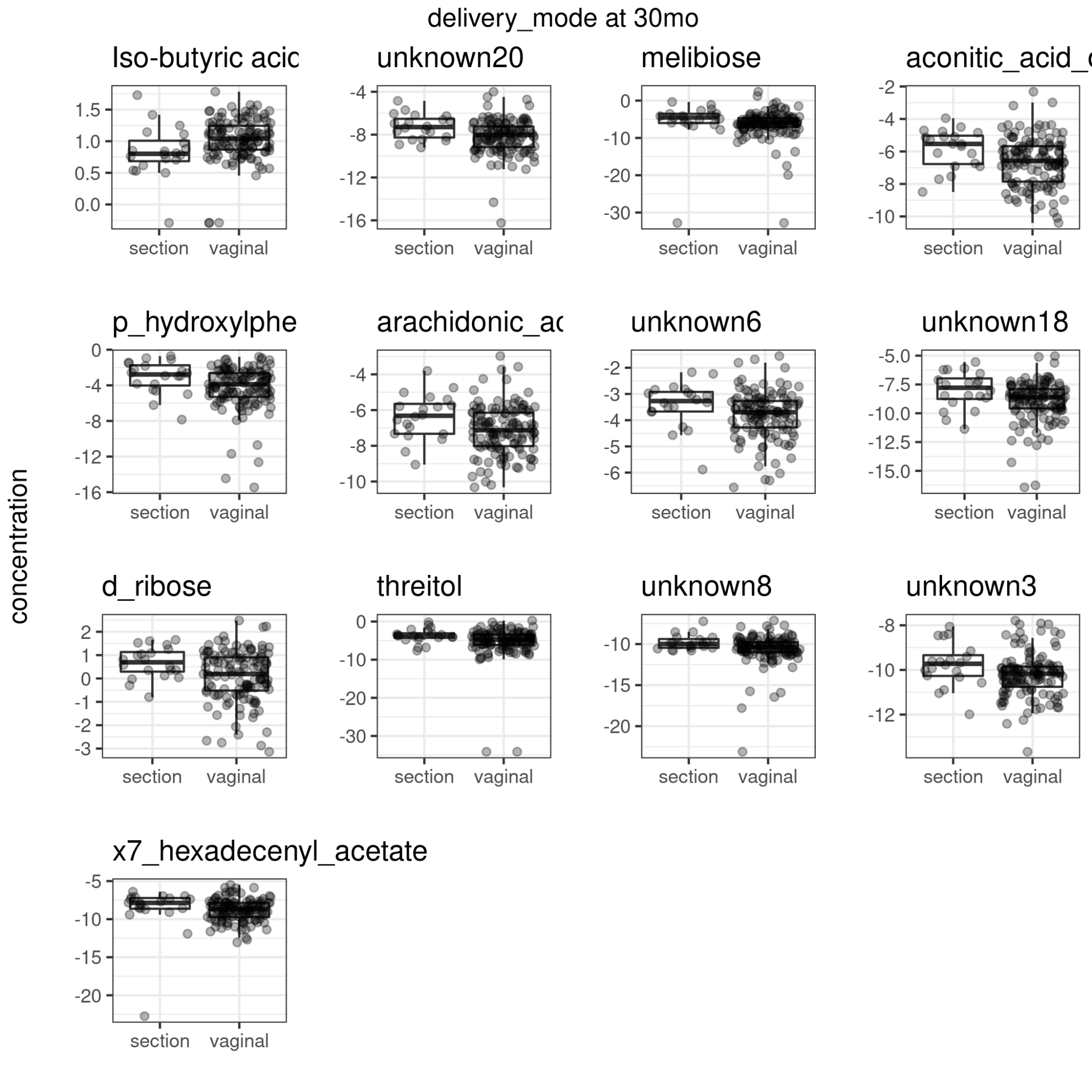

Figure 24. Delivery mode and metabolite concentrations at 30 mo.

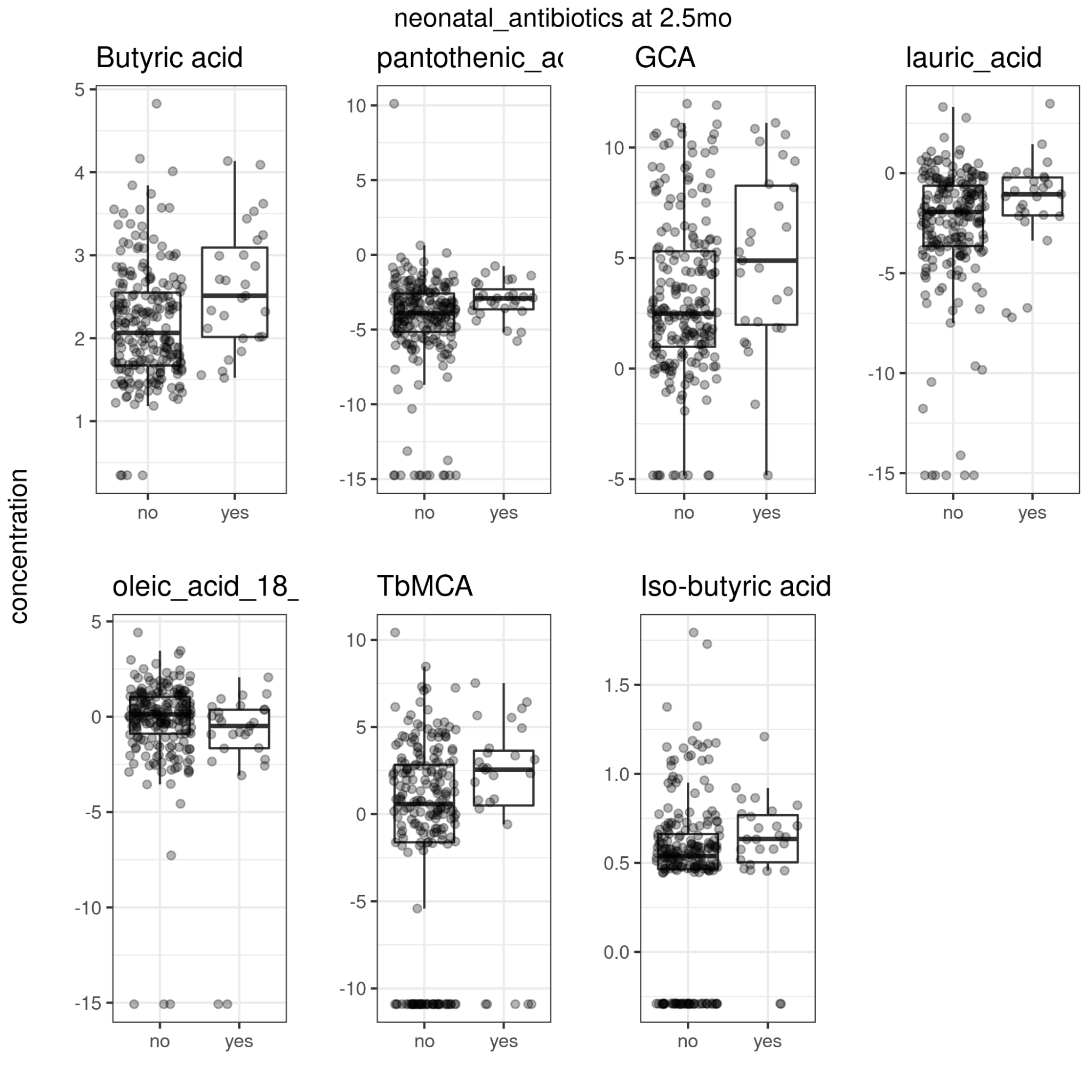

Figure 25. Neonatal antibiotic exposure and metabolite concentrations at 2.5 mo.

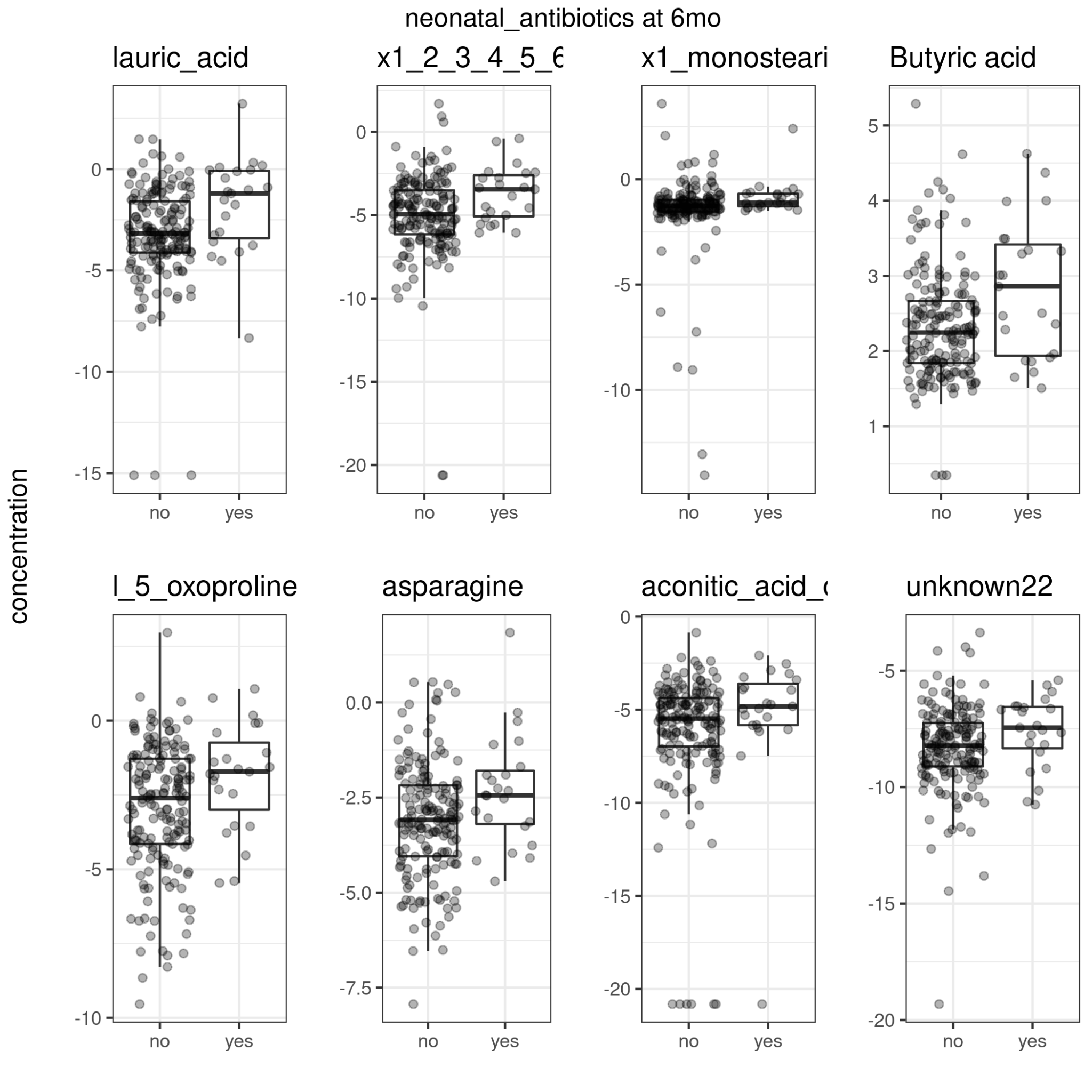

Figure 26. Neonatal antibiotic exposure and metabolite concentrations at 6 mo.

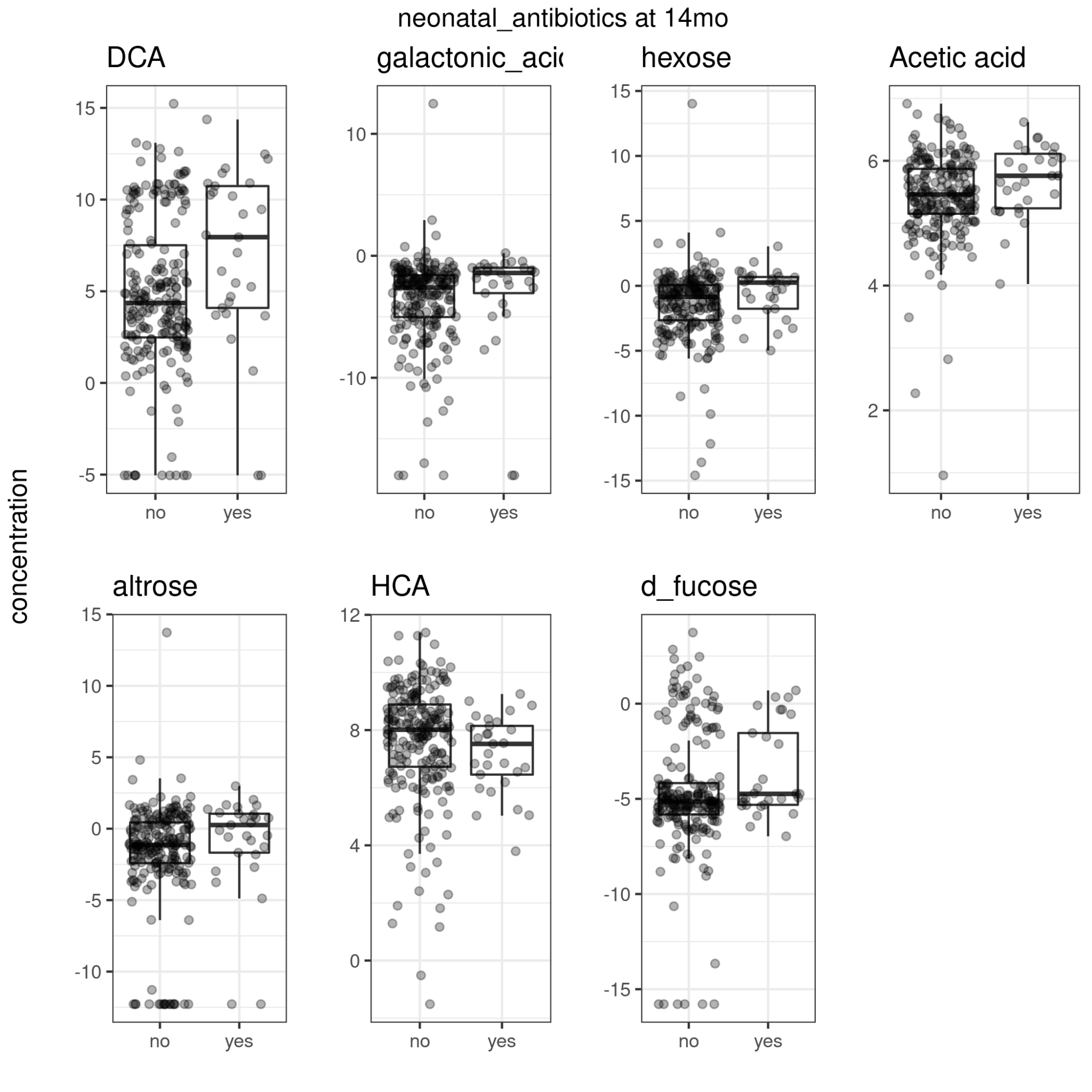

Figure 27. Neonatal antibiotic exposure and metabolite concentrations at 14 mo.

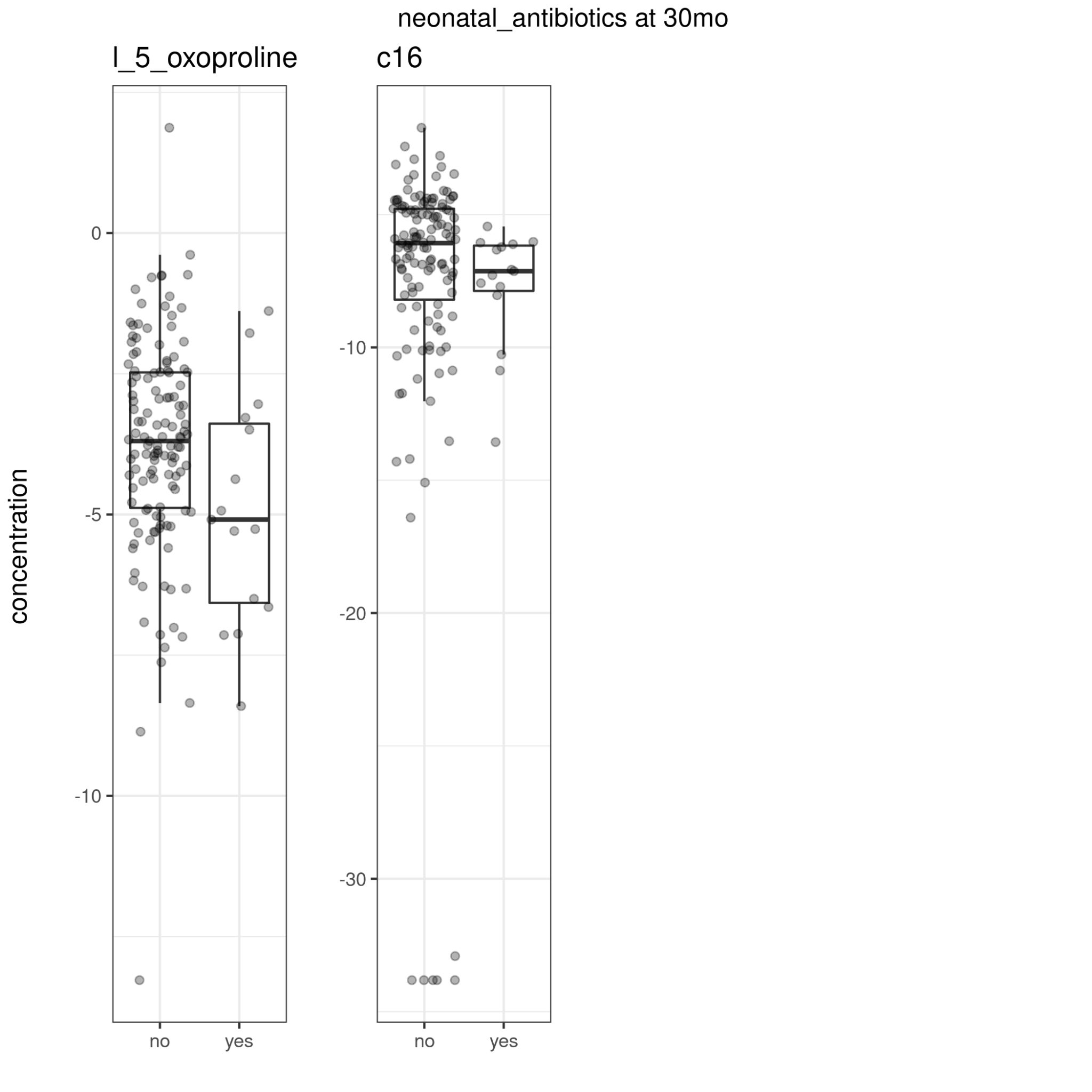

Figure 28. Neonatal antibiotic exposure and metabolite concentrations at 30 mo.

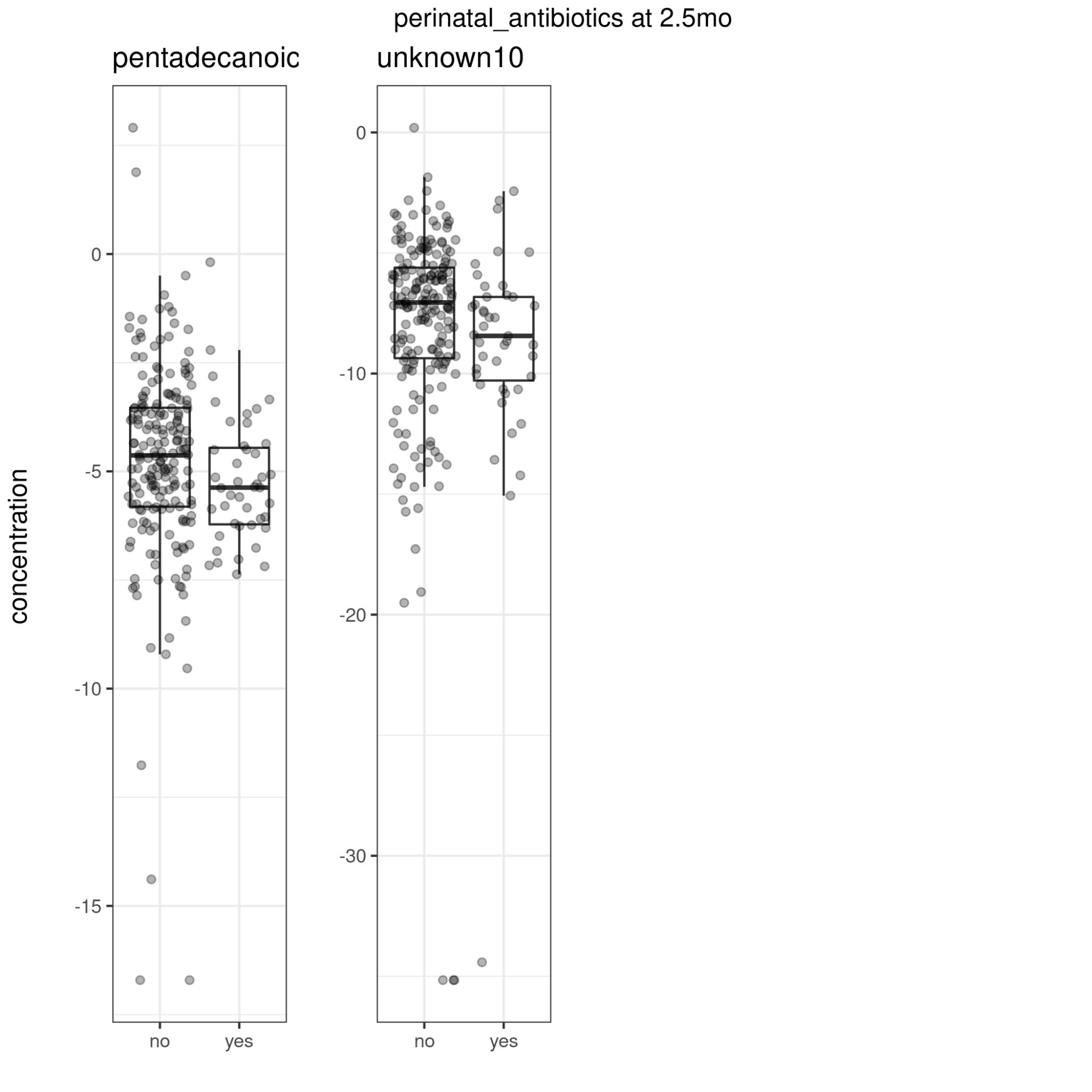

Figure 29. Maternal perinatal antibiotic exposure and metabolite concentrations at 2.5 mo.

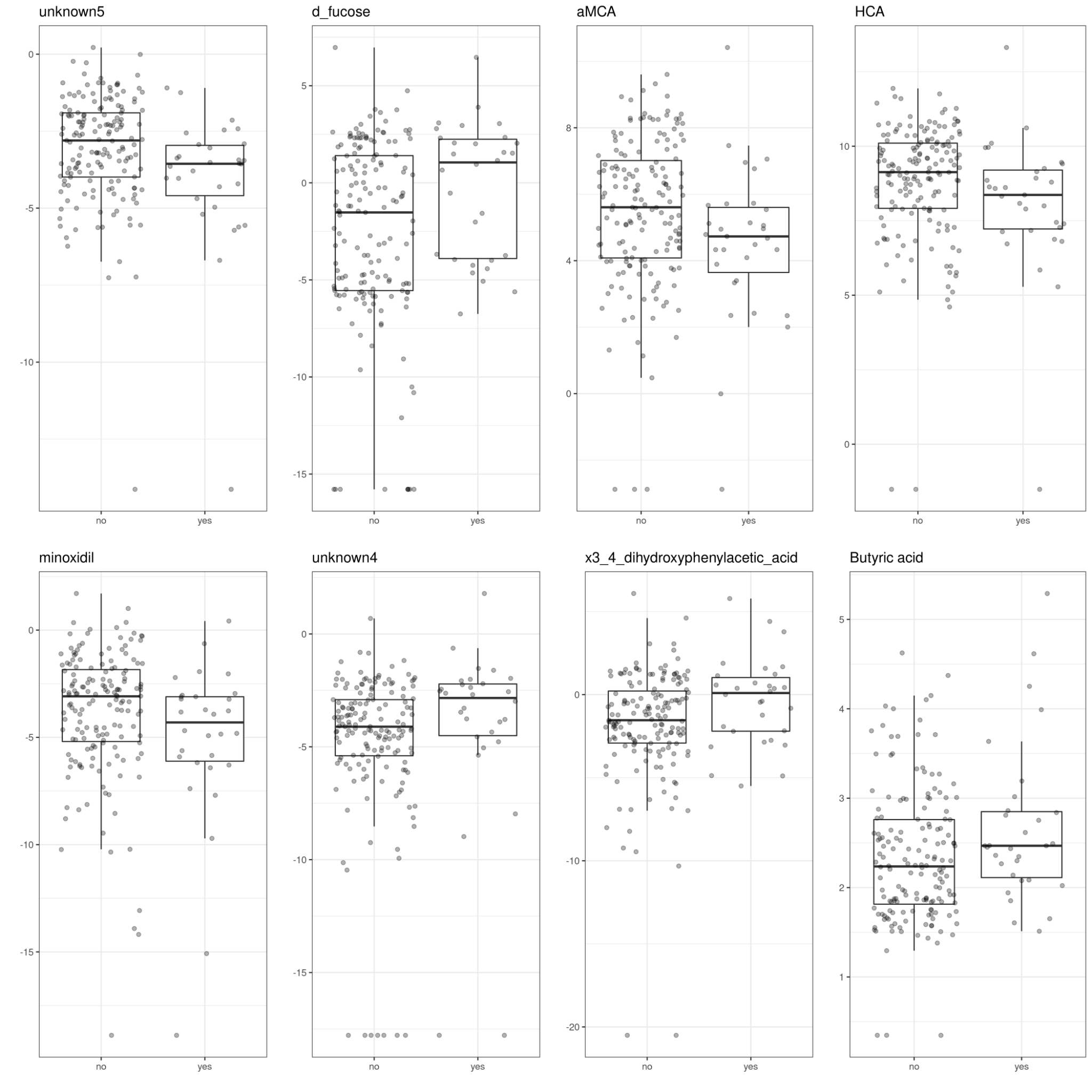

Figure 30. Maternal perinatal antibiotic exposure and metabolite concentrations at 6 mo.

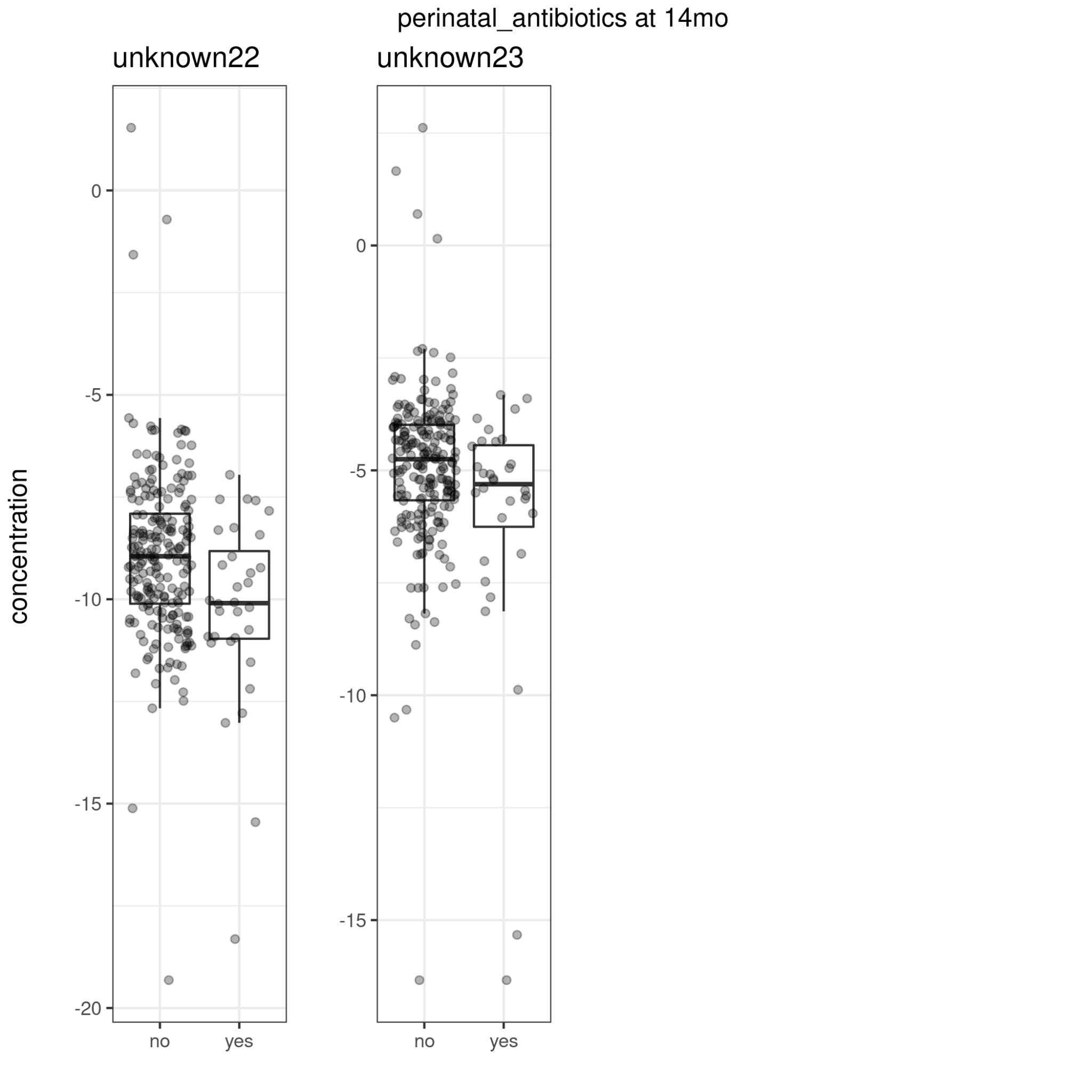

Figure 31. Maternal perinatal antibiotic exposure and metabolite concentrations at 14 mo.

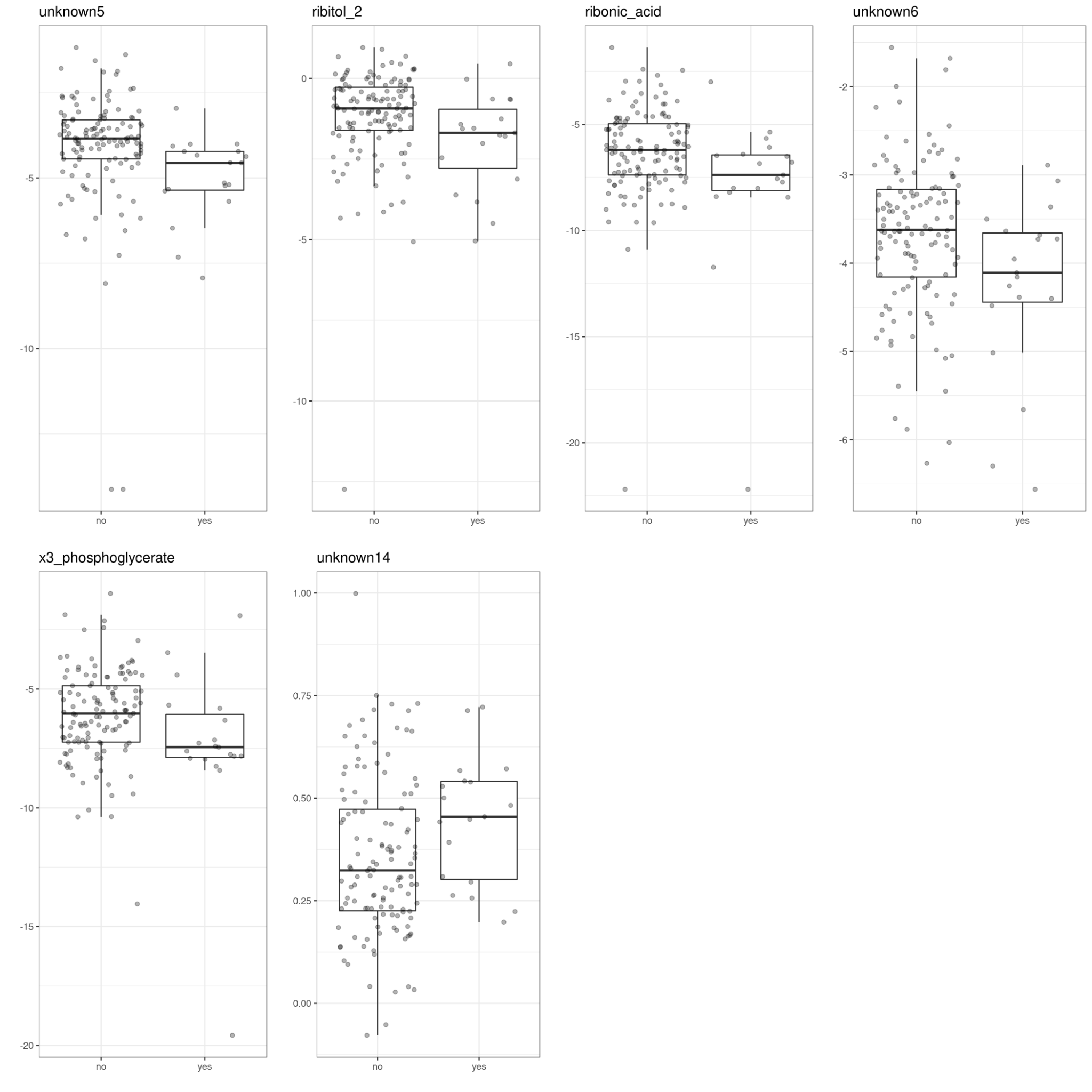

Figure 32. Maternal perinatal antibiotic exposure and metabolite concentrations at 30 mo.

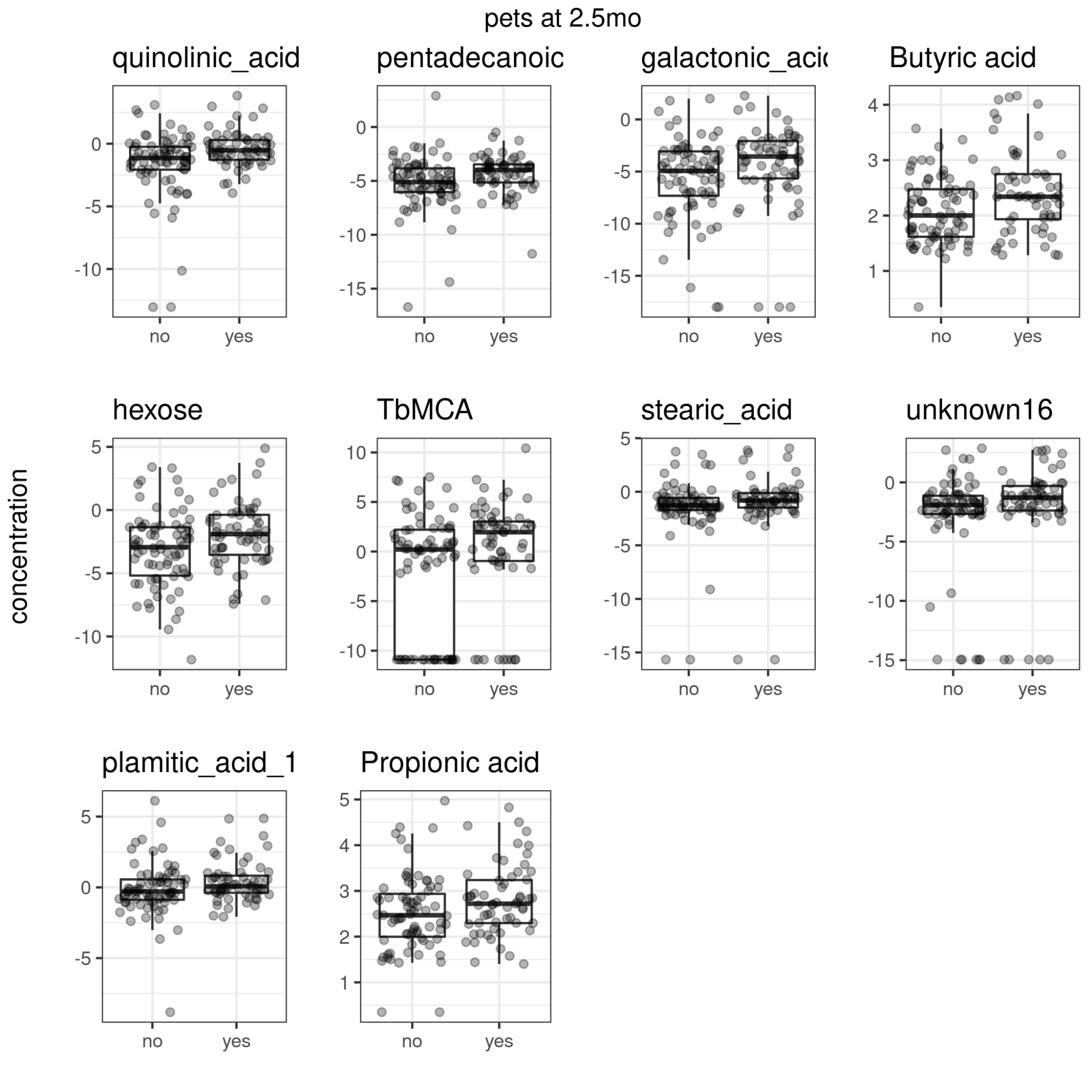

Figure 33. Having pets in household and metabolite concentrations at 2.5 mo.

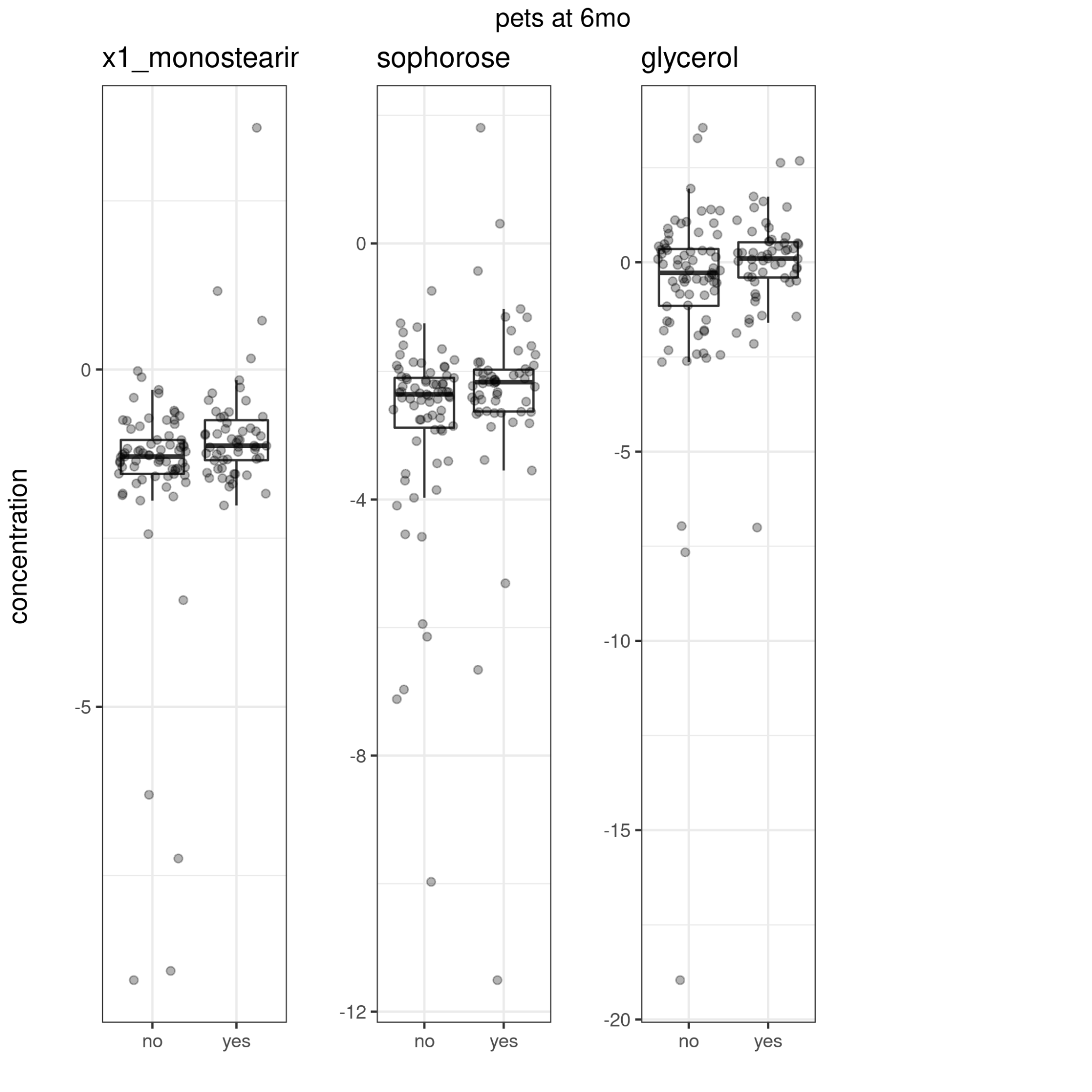

Figure 34. Having pets in the household and metabolite concentrations at 6 mo.

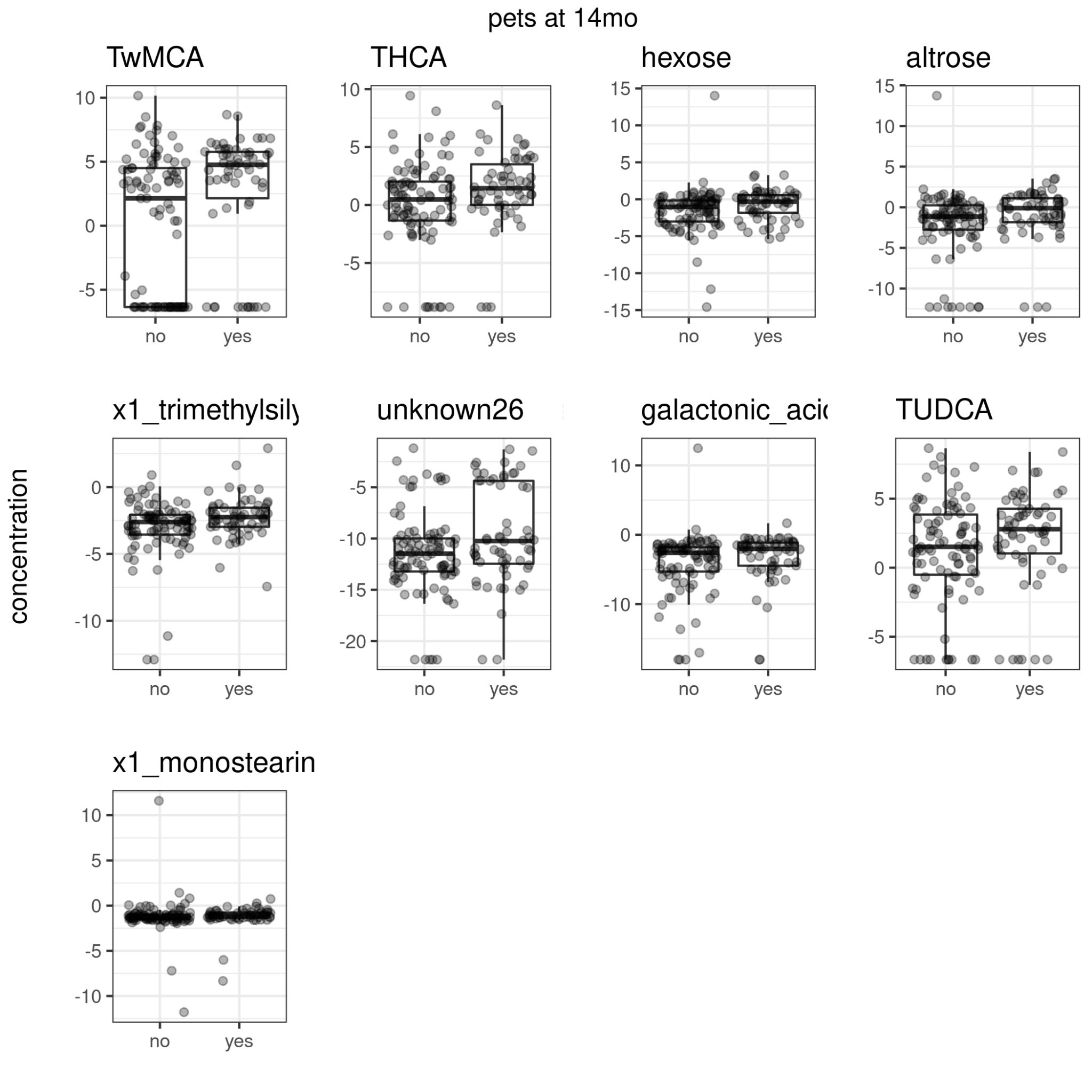

Figure 35. Having pets in the household and metabolite concentrations at 14 mo.

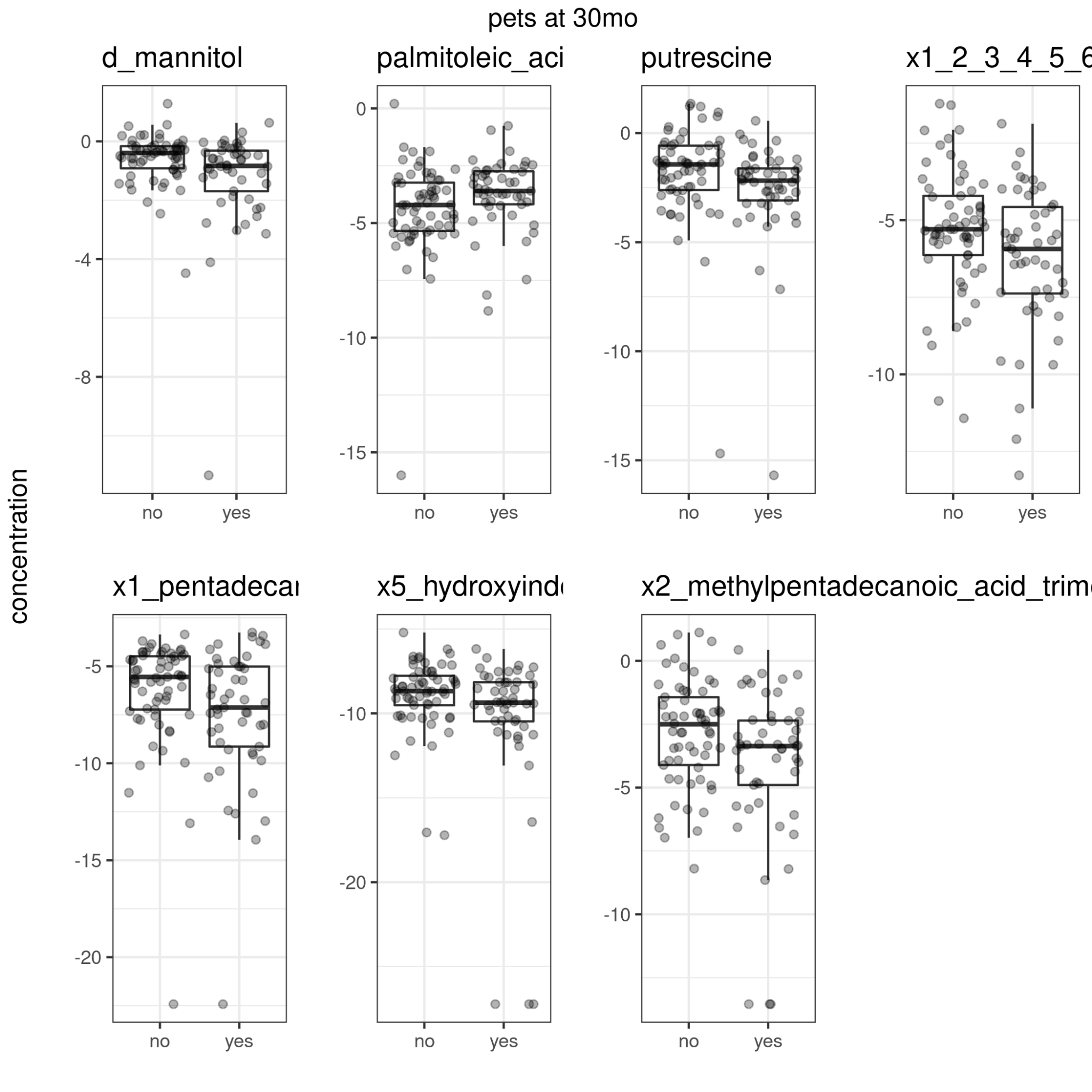

Figure 36. Having pets in the household and metabolite concentrations at 30 mo.

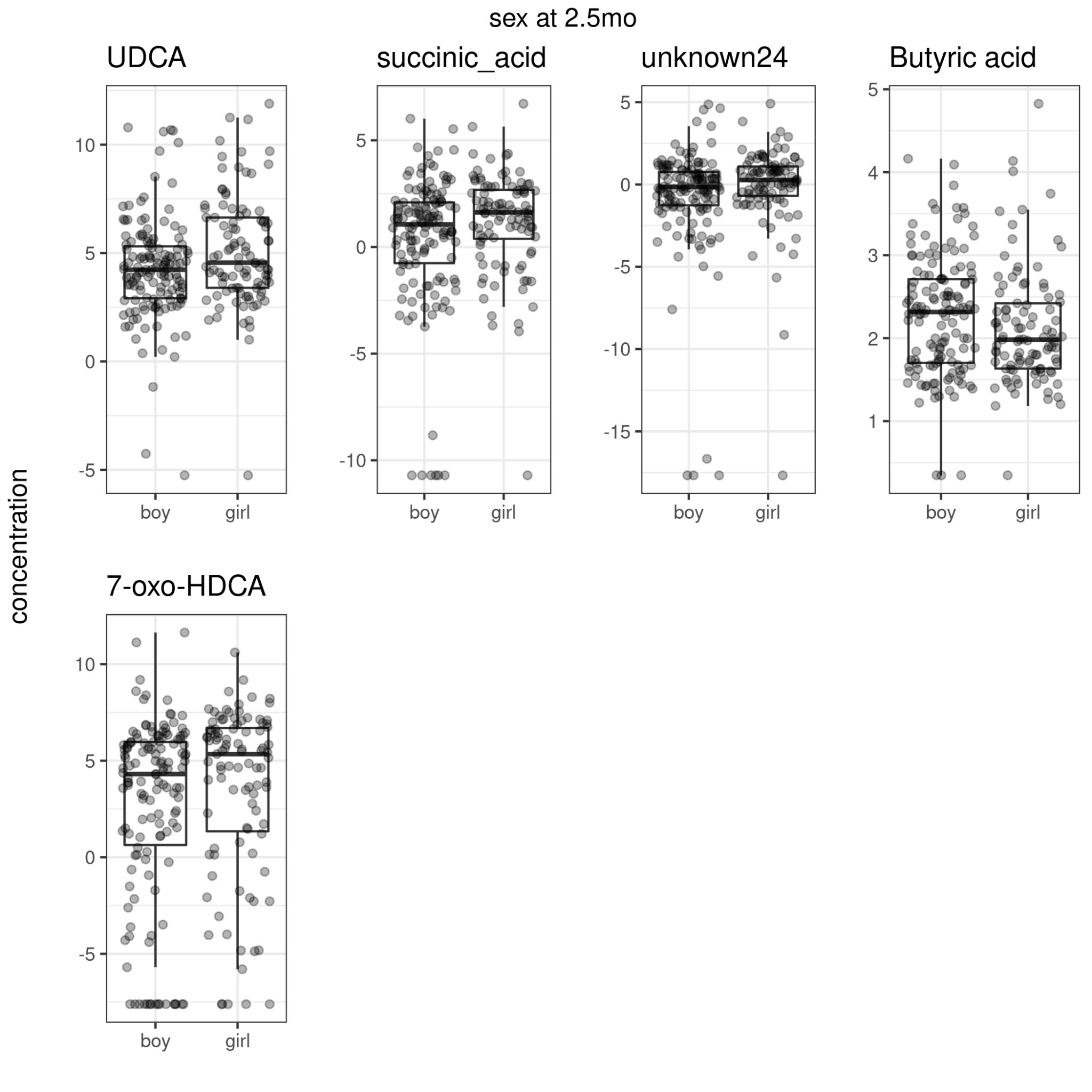

Figure 37. Child sex and metabolite concentrations at 2.5 mo.

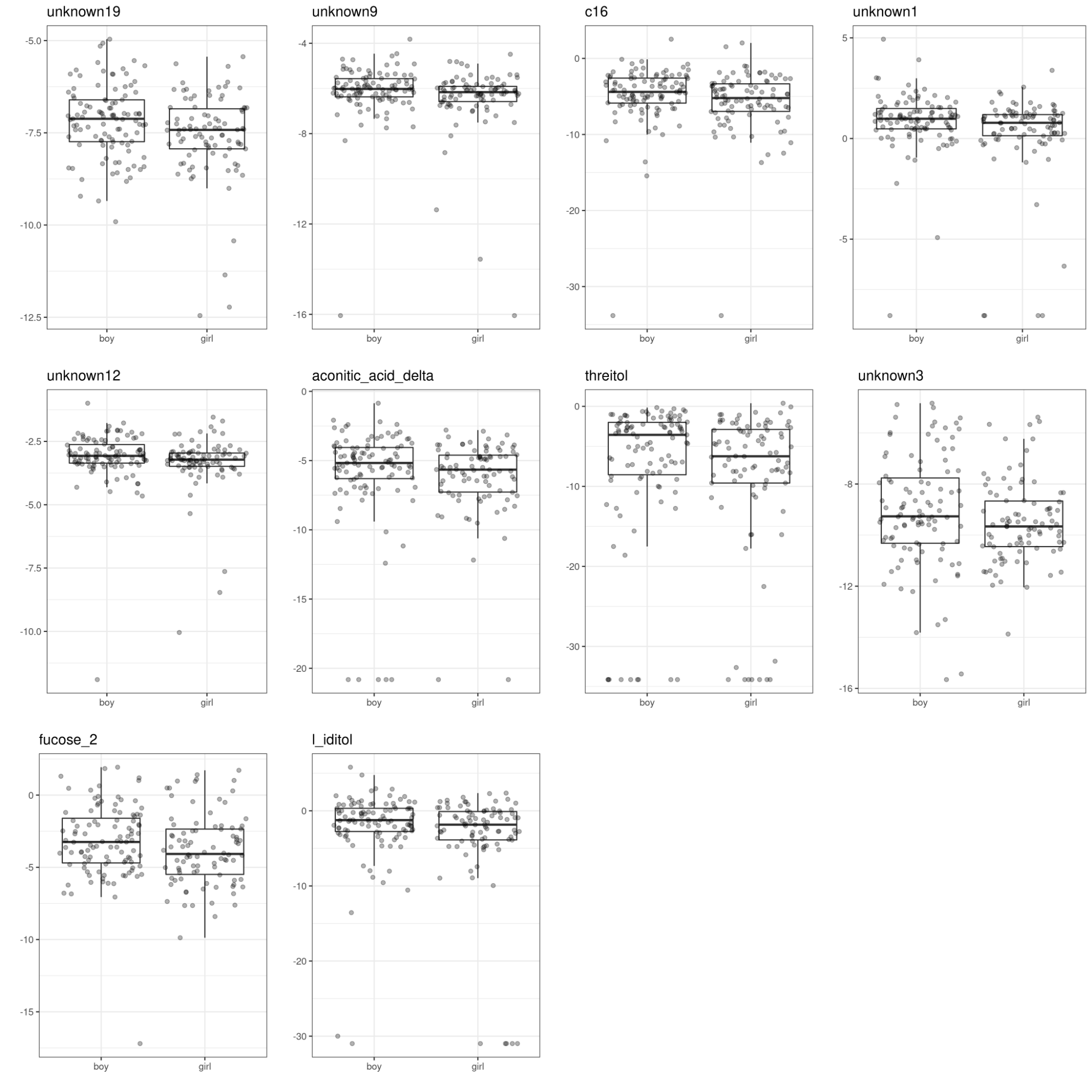

Figure 38. Child sex and metabolite concentrations at 6 mo.

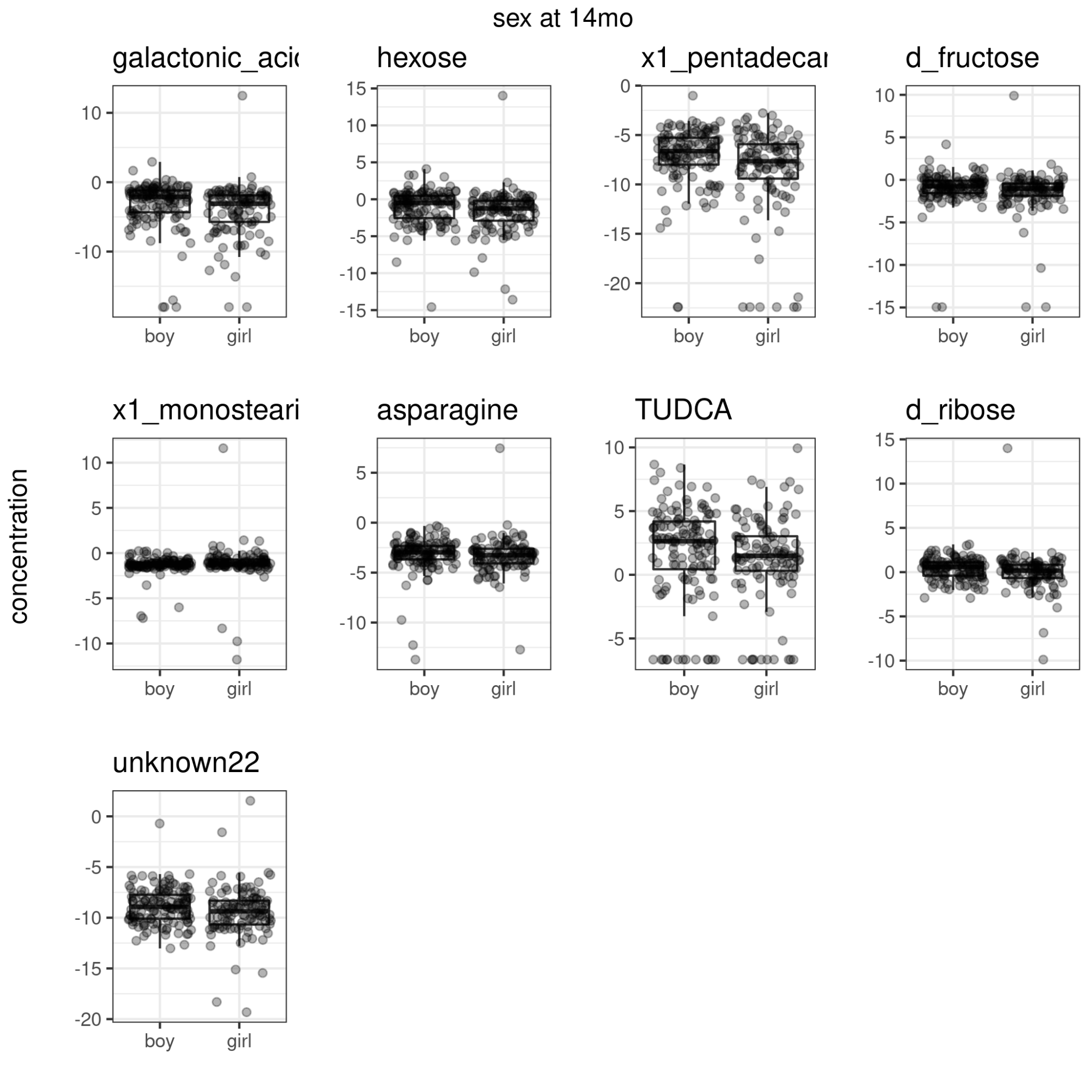

Figure 39. Child sex and metabolite concentrations at 14 mo.

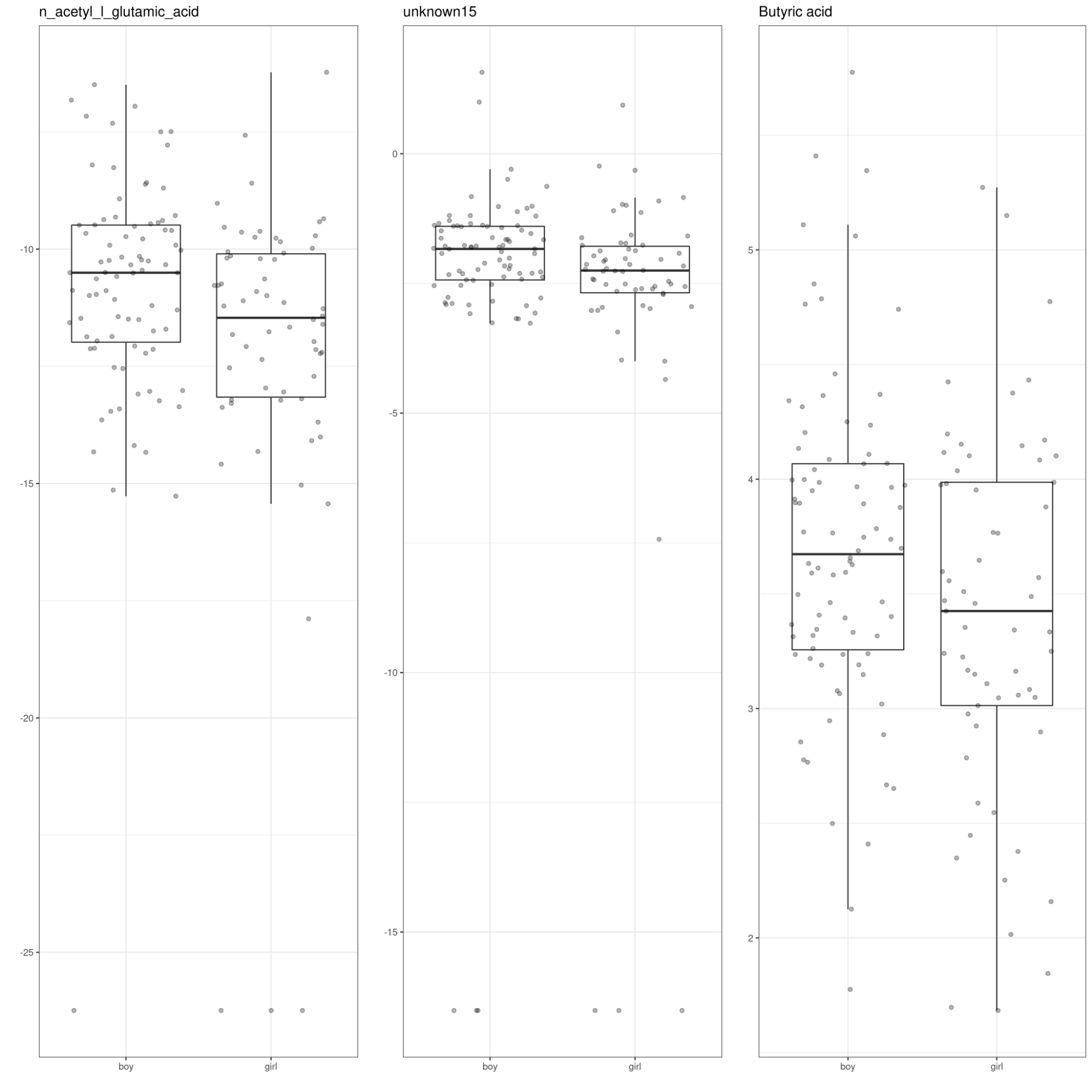

Figure 40. Child sex and metabolite concentrations at 30 mo.

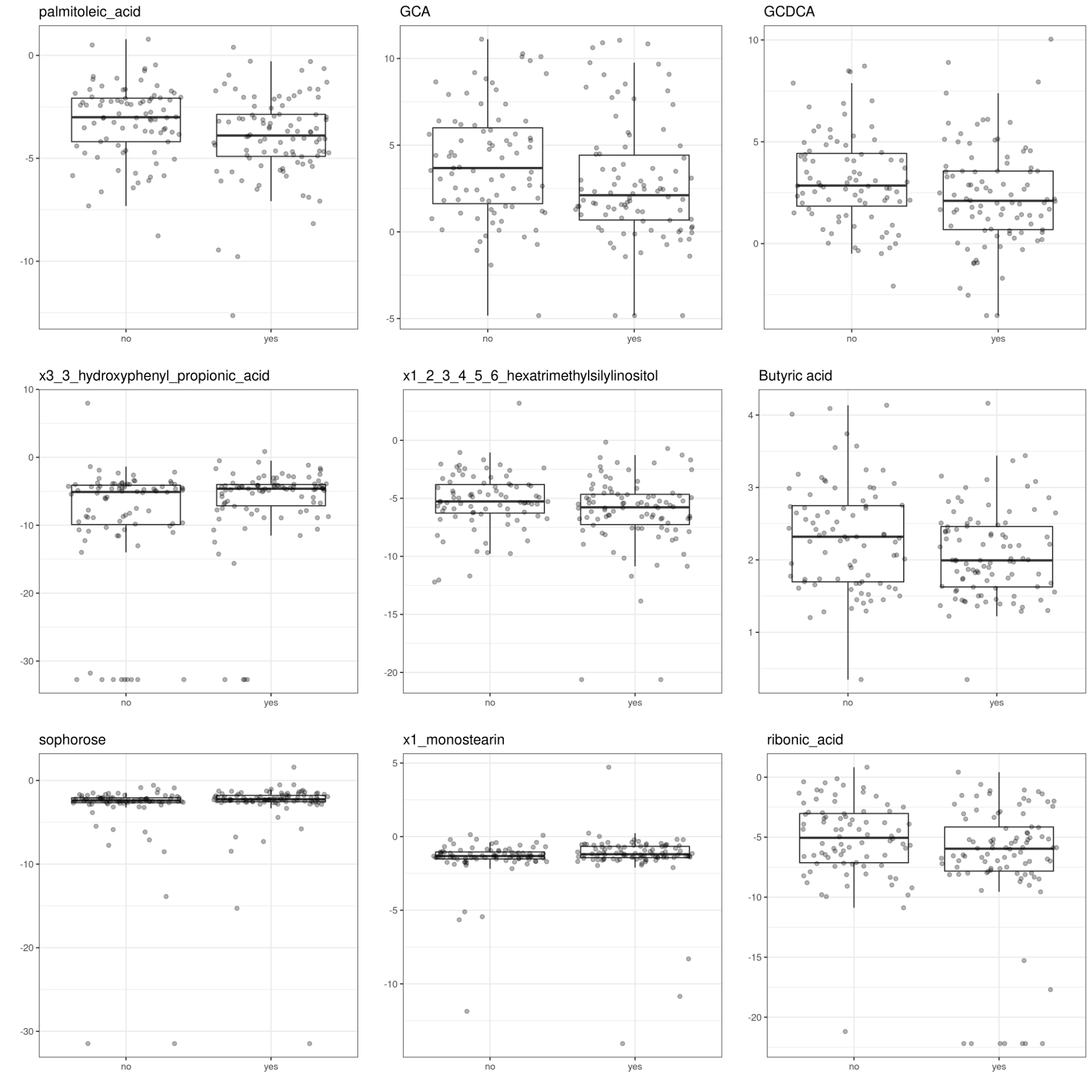

Figure 41. Having siblings and metabolite concentrations at 2.5 mo.

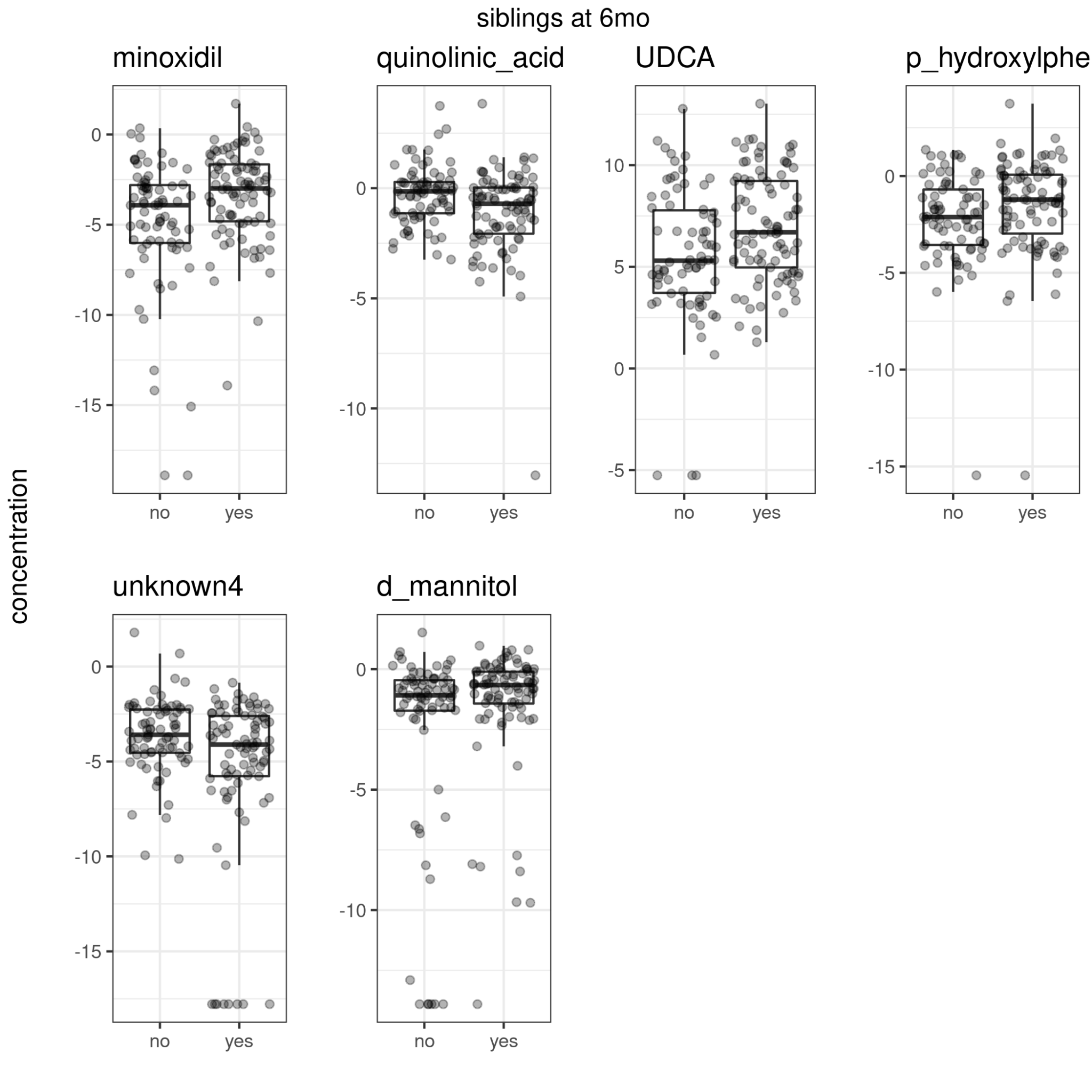

Figure 42. Having siblings and metabolite concentrations at 6 mo.

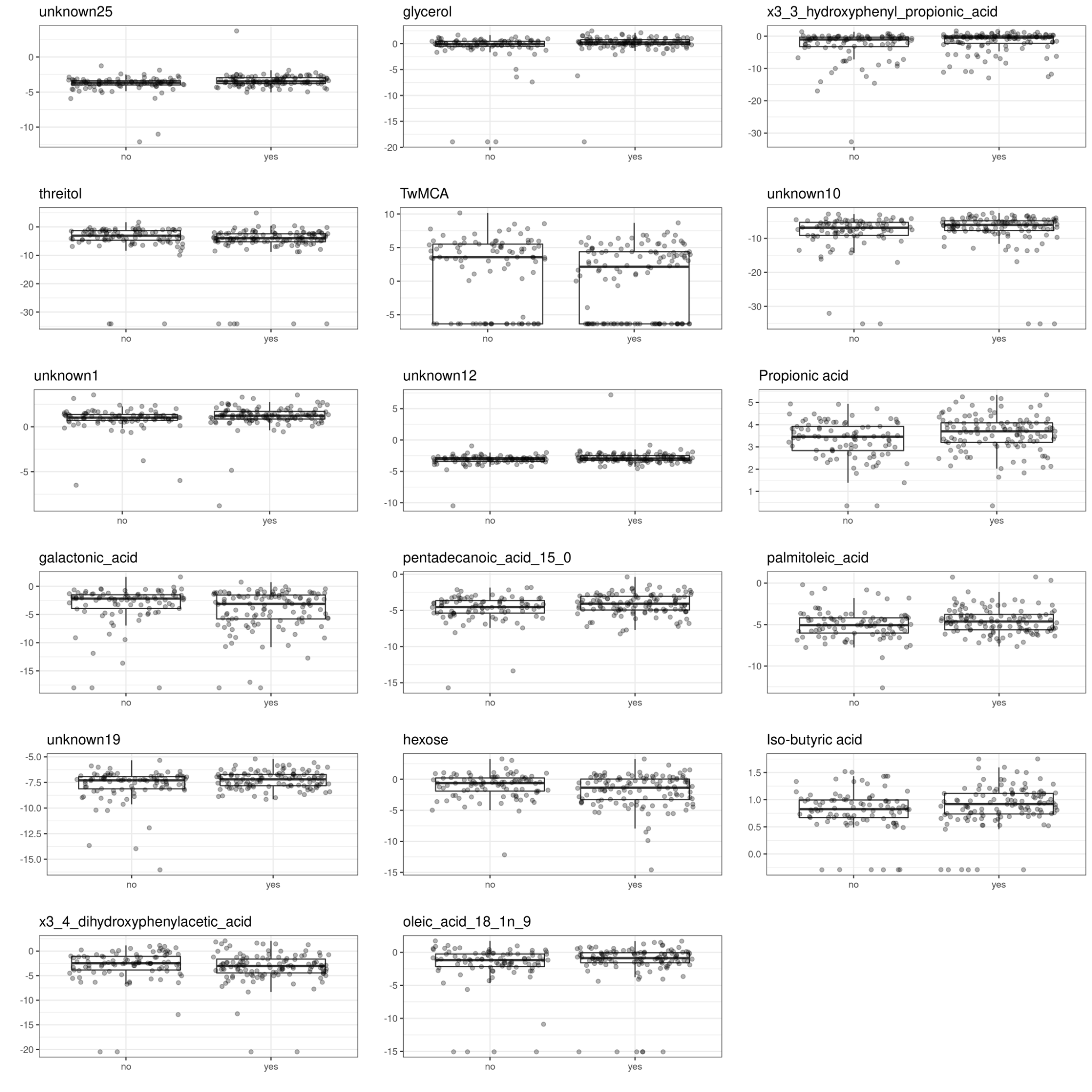

Figure 43. Having siblings and metabolite concentrations at 14 mo.

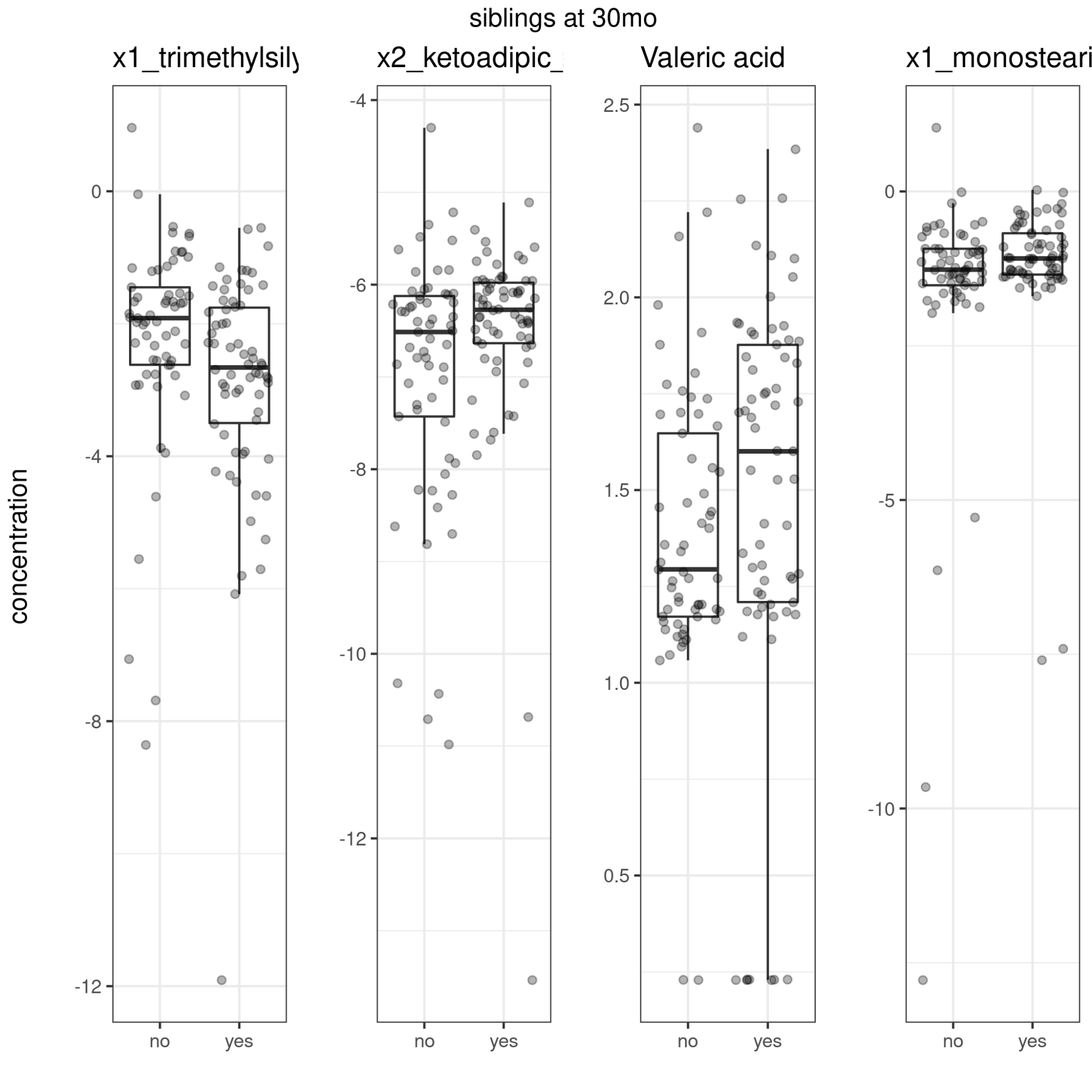

Figure 44. Having siblings and metabolite concentrations at 30 mo.

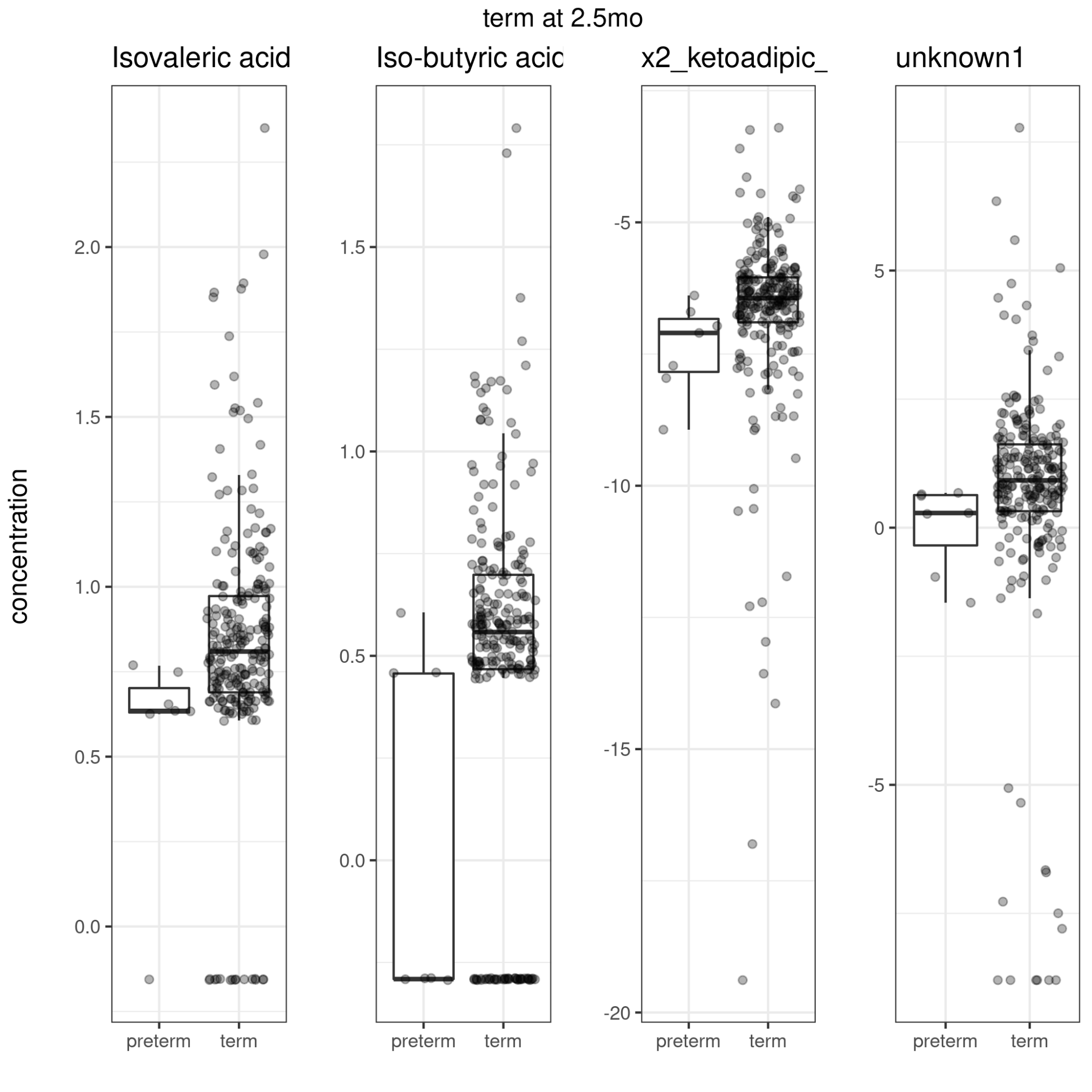

Figure 45. Term delivery and metabolite concentrations at 2.5 mo.

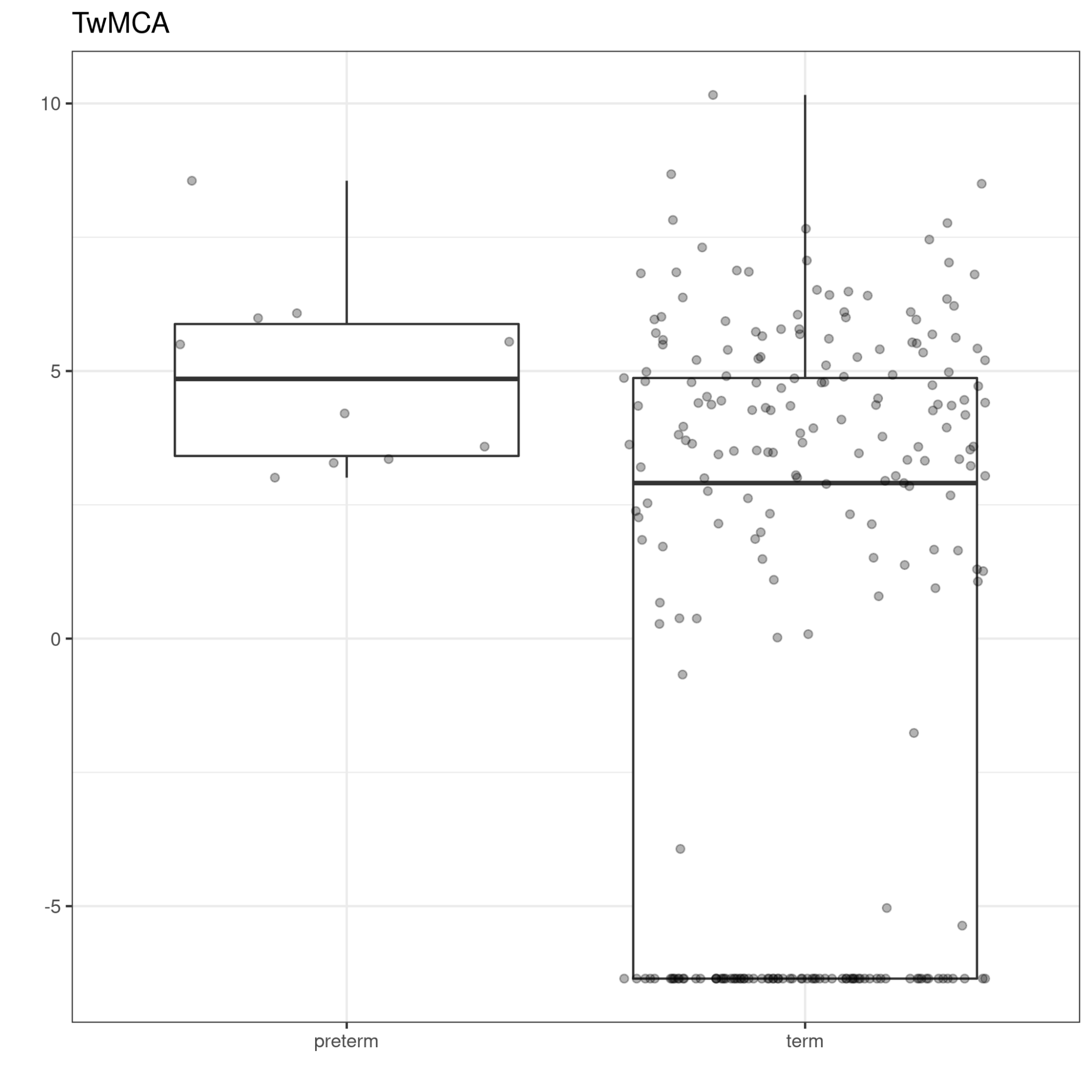

Figure 46. Term delivery and metabolite concentrations at 14 mo.

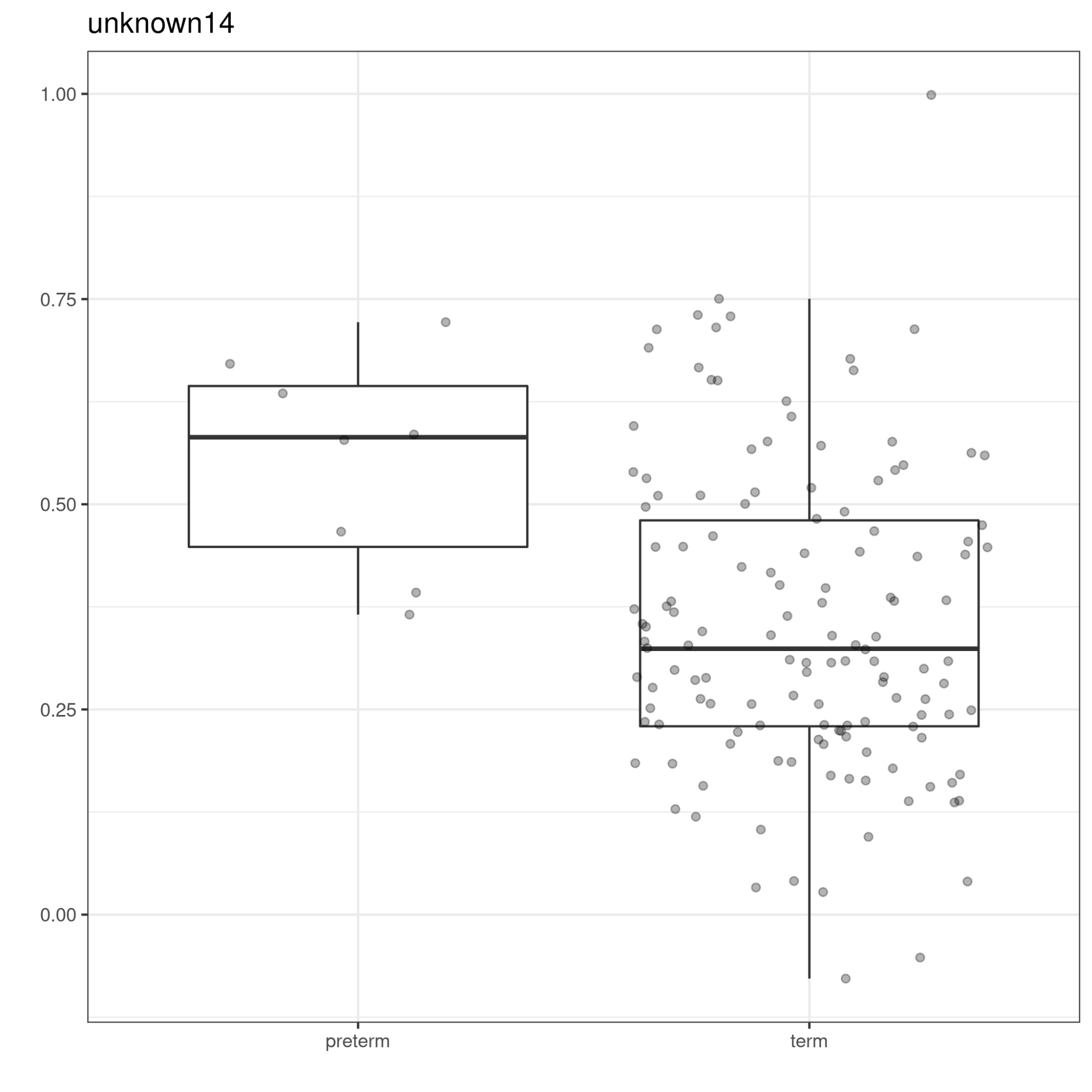

Figure 47. Term delivery and metabolite concentrations at 30 mo.

#### Categorized bile acids and demographic factors

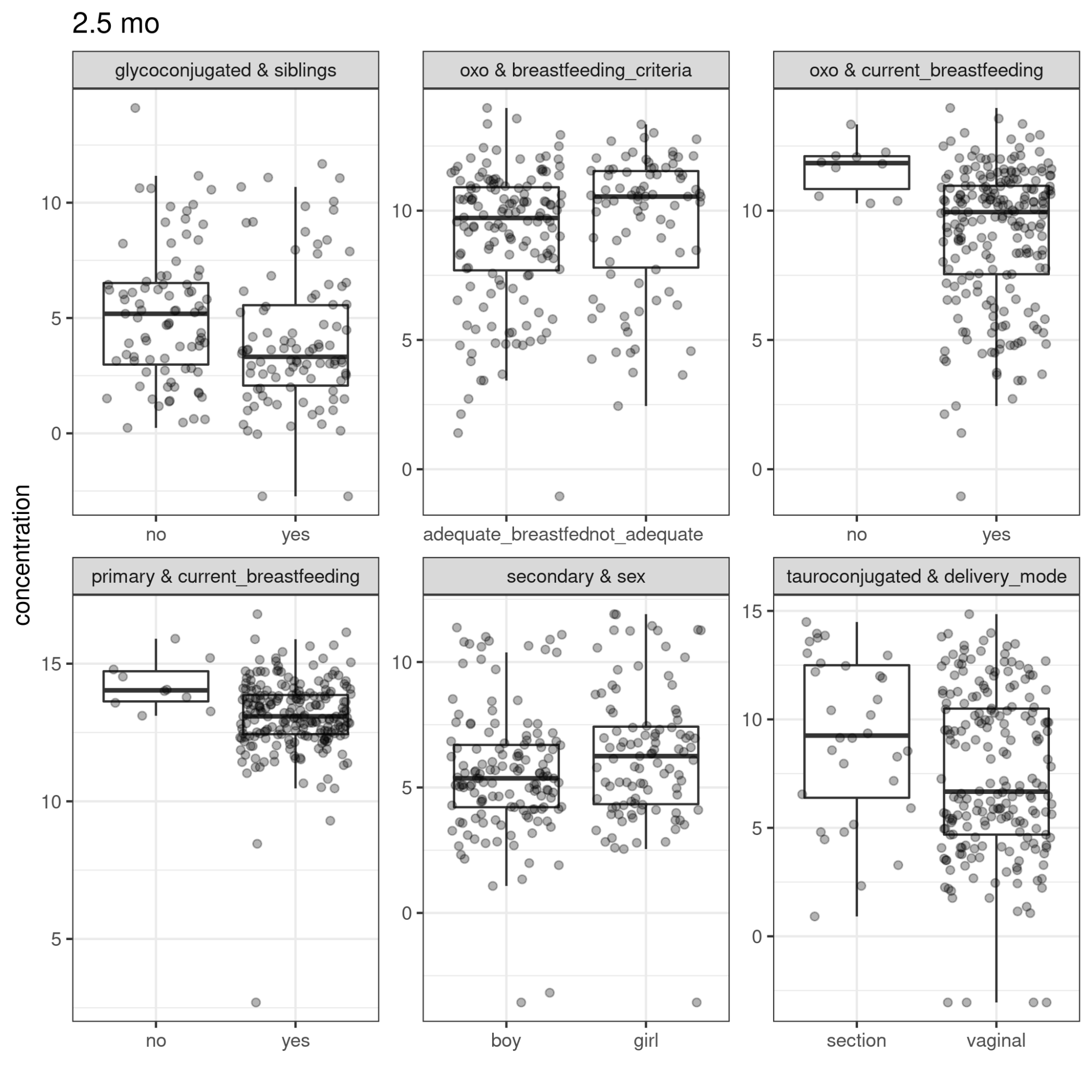

Figure 48. Categorized bile acid concentrations and demographic factors at 2.5 mo

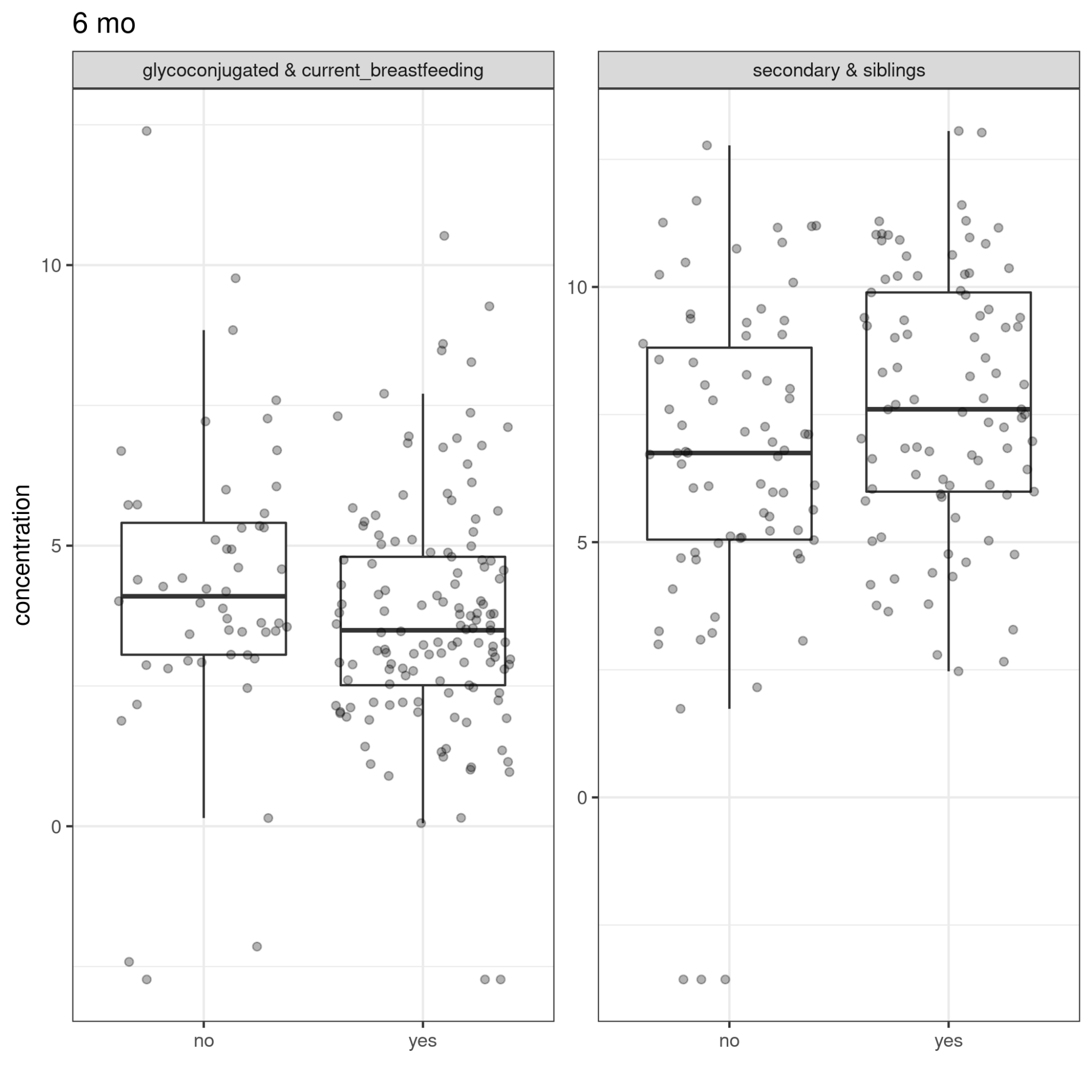

Figure 49. Categorized bile acid concentrations and demographic factors at 6 mo

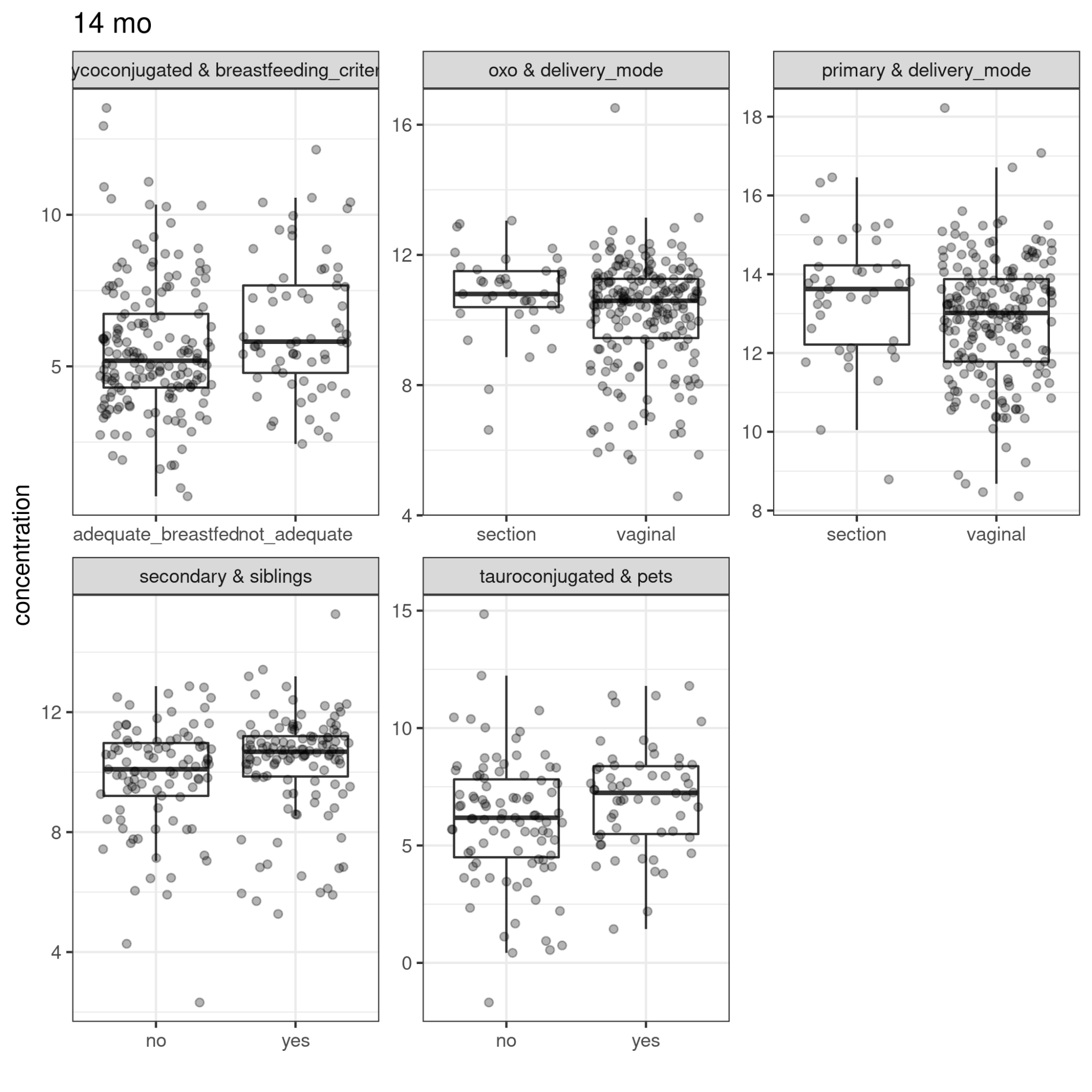

Figure 50. Categorized bile acid concentrations and demographic factors at 14 mo

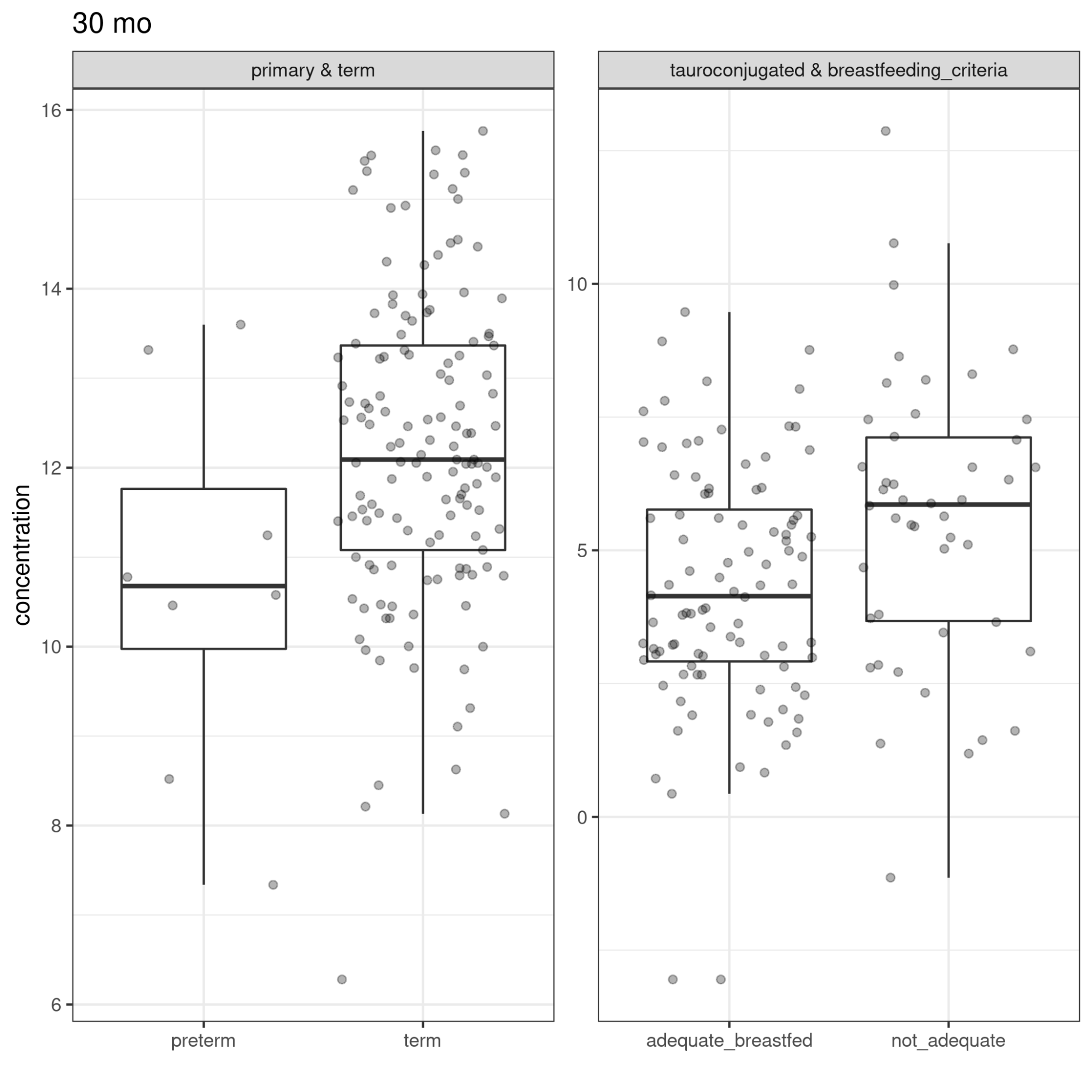

Figure 51. Categorized bile acid concentrations and demographic factors at 30 mo

### Microbiome cluster top drivers

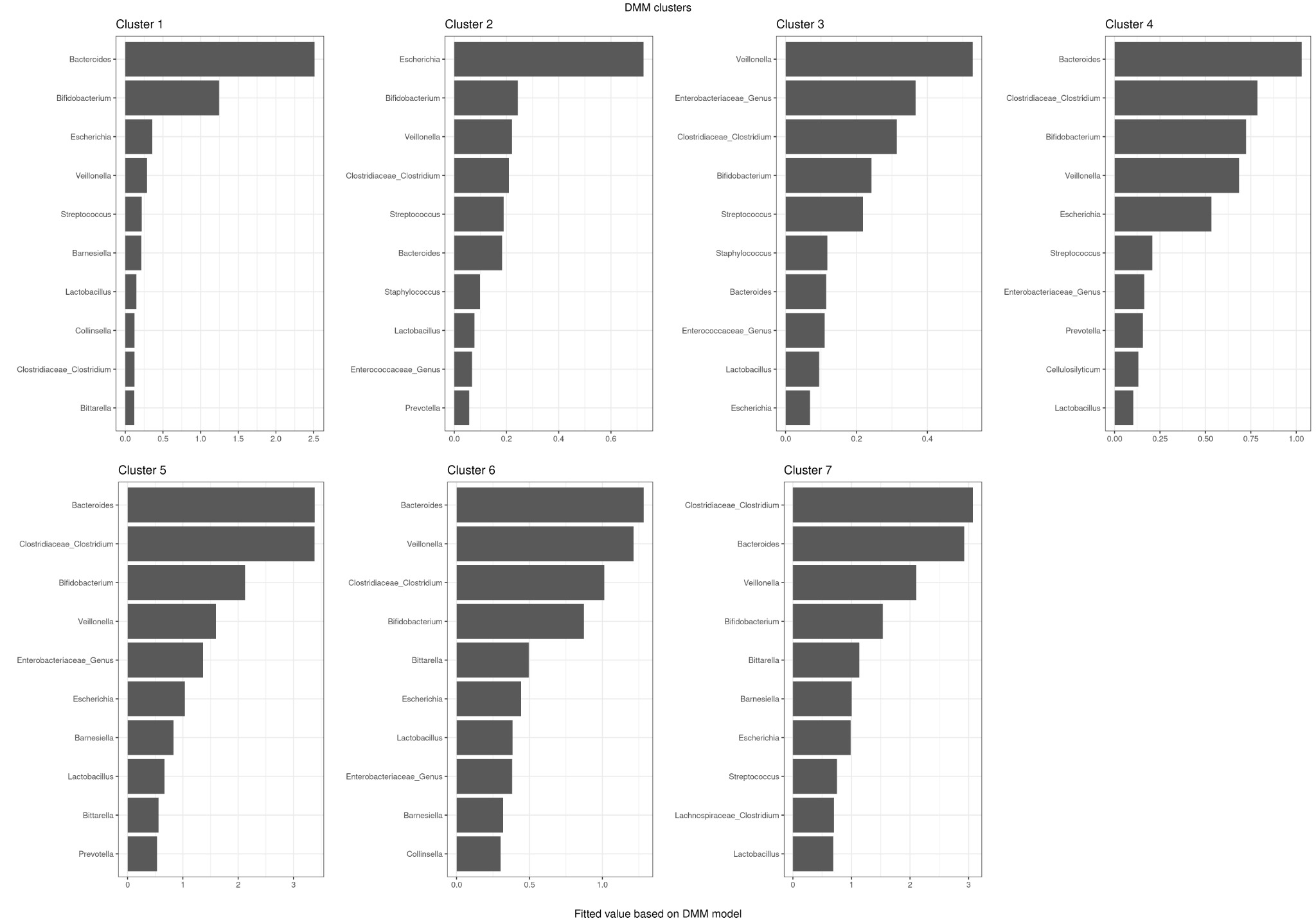

Figure 52. Contribution of taxonomic groups to the DMM clusters.

#### Cluster variability and weights

Table 4. Clusters differed by the variability (theta). Lower theta values indicate higher variance, and the C5 and C7 have the most variance in the taxonomic contributions. Moreover, the C1 seems to have higher variance compared with C2 and C3.

| cluster | pi | theta |
| --- | --- | --- |
| 1 | 0.1 | 6.48 |
| 2 | 0.14 | 2.48 |
| 3 | 0.14 | 2.73 |
| 4 | 0.2 | 5.73 |
| 5 | 0.09 | 23.67 |
| 6 | 0.17 | 10.54 |
| 7 | 0.15 | 25.67 |

### Clusters and demographic factors

Table 5. Demographic factors associated with cluster membership per timepoint.

| timepoint | var | statistic | chisq_df | p_value | dunns_groups | q |
| --- | --- | --- | --- | --- | --- | --- |
| 6 | term | 36.2 | 3 | 0.00E+00 | 4.00E+00 | 0 |
| 2.5 | delivery_mode | 33.7 | 3 | 0.00E+00 | 3.00E+00 | 0 |
| 2.5 | perinatal_antibiotics | 12.3 | 3 | 0.00647 | 1 | 0.0733 |
| 2.5 | siblings | 11.2 | 3 | 0.01065 | 2 | 0.09054 |
| 2.5 | breastfeeding_criteria | 9.5 | 3.00E+00 | 0.02354 | 0 | 0.16007 |
| 30 | perinatal_antibiotics | 6.7 | 2 | 0.03541 | 2 | 0.20064 |

Table 6. Dunn’s posthoc test for significant cluster differences in timepoint-wise group comparison

| timepoint | var | clusters | p.adj | statistic |
| --- | --- | --- | --- | --- |
| 2.5 | delivery_mode | 1 vs 2 | 0.03566 | -2.60E+00 |
| 2.5 | delivery_mode | 1 vs 3 | 0 | -5.70E+00 |
| 2.5 | delivery_mode | 2 vs 3 | 0.0034 | -3.4 |
| 6 | term | 1 vs 2 | 0.01729 | 2.9 |
| 6 | term | 1 vs 4 | 4.70E-04 | 3.9 |
| 6 | term | 2 vs 3 | 0.04675 | -2.4 |
| 6 | term | 3 vs 4 | 7.00E-05 | 4.4 |

Table 7. Cross-tabulations for delivery mode at 2.5 months and preterm delivery at 6 months

| cluster | section | vaginal | vag_prop |
| --- | --- | --- | --- |
| 1 | 4 | 114 | 97% |
| 2 | 25 | 139 | 85% |
| 3 | 45 | 107 | 70% |
| 4 | 0 | 3 | 100% |
| cluster | preterm | term | term_prop |
| 1 | 1 | 1 | 50% |
| 2 | 1 | 8 | 89% |
| 3 | 2 | 4 | 67% |
| 4 | 4 | 235 | 98% |

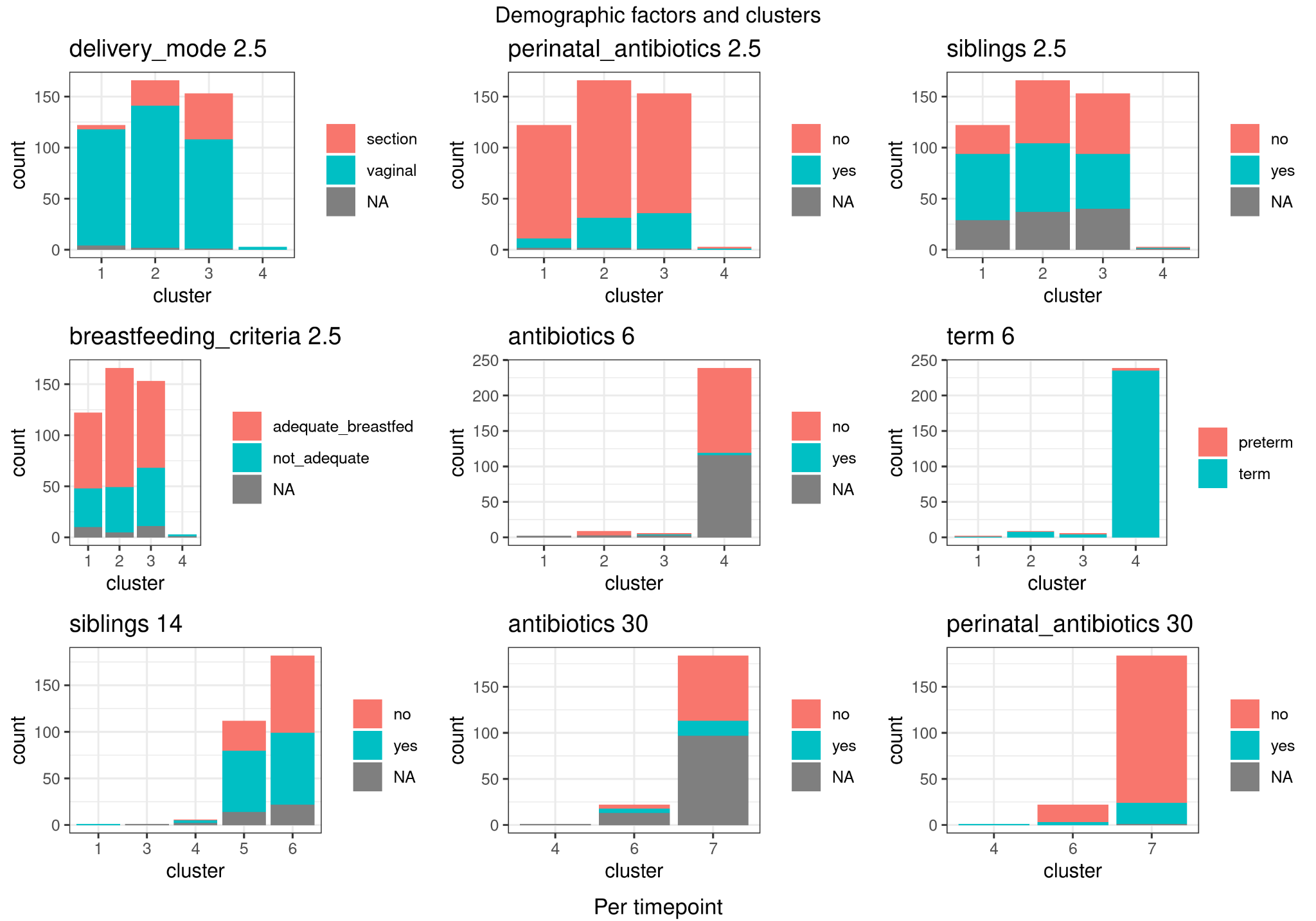

Figure 53. Differences in demographic factors in clusters per timepoint. Findings with p-value <0.05 are visualized.

### Metabolome and microbiome

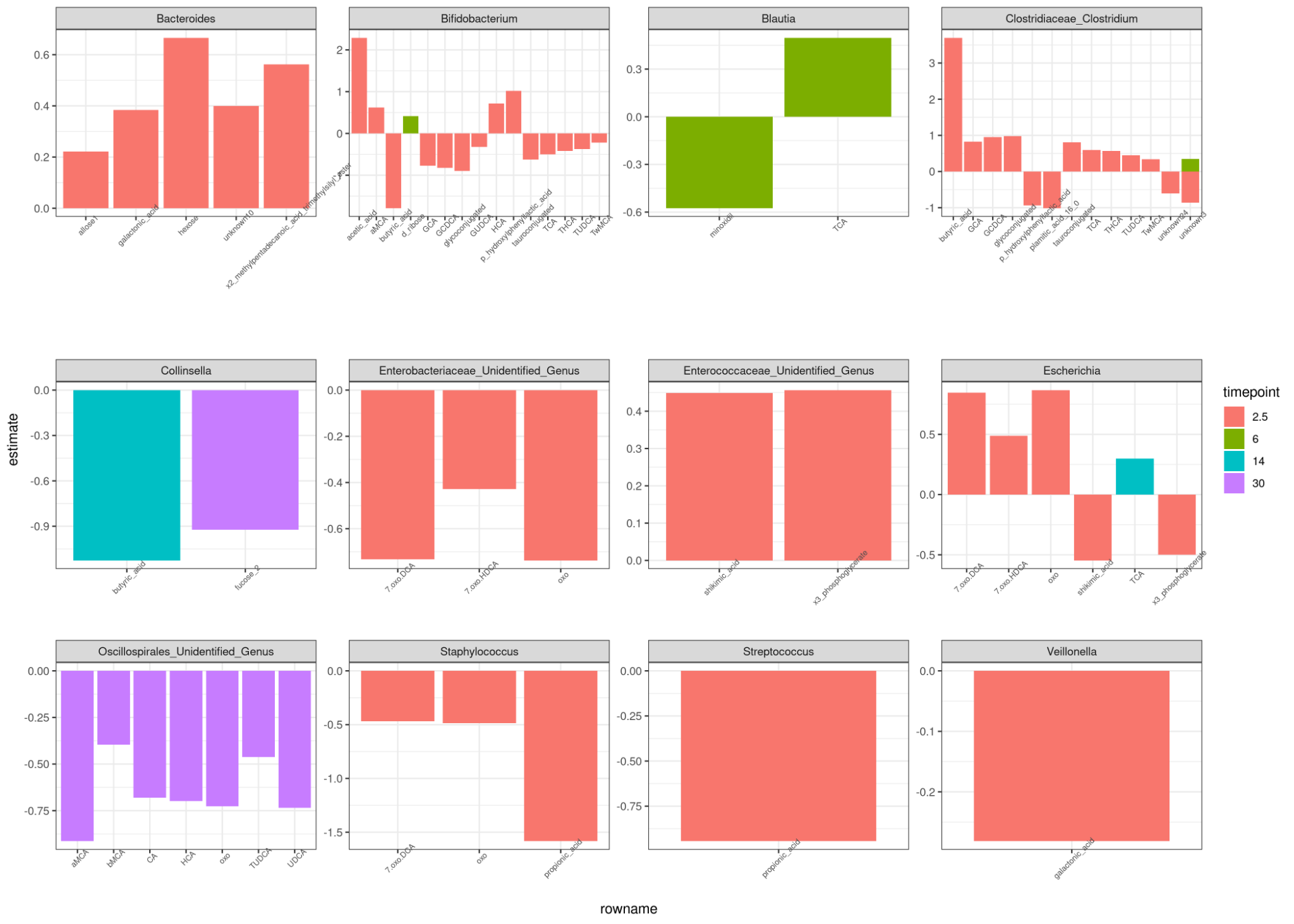

Figure 54. Estimates for significant (q < 0.05) findings from ALDEx2 across all timepoints per genus.

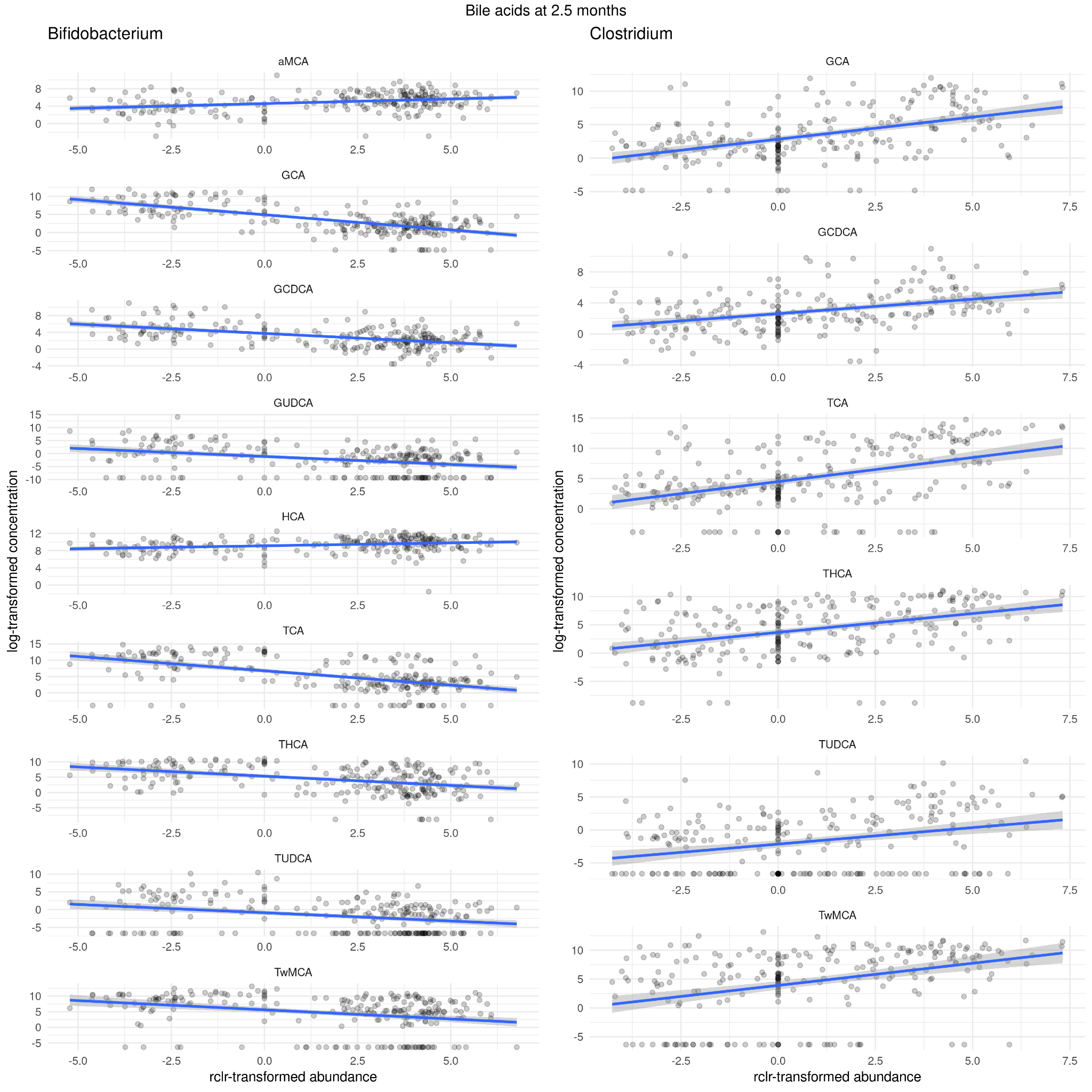

Figure 55. ALDEx2 indicated multiple associations between bile acids concentrations and Bifidobacterium and Clostridium abundances at 2.5 mo. Interestingly, they were in opposing directions. Bile acid data is log-transformed whereas abundance data is robust centered log-transformed (rclr) for the visualization.

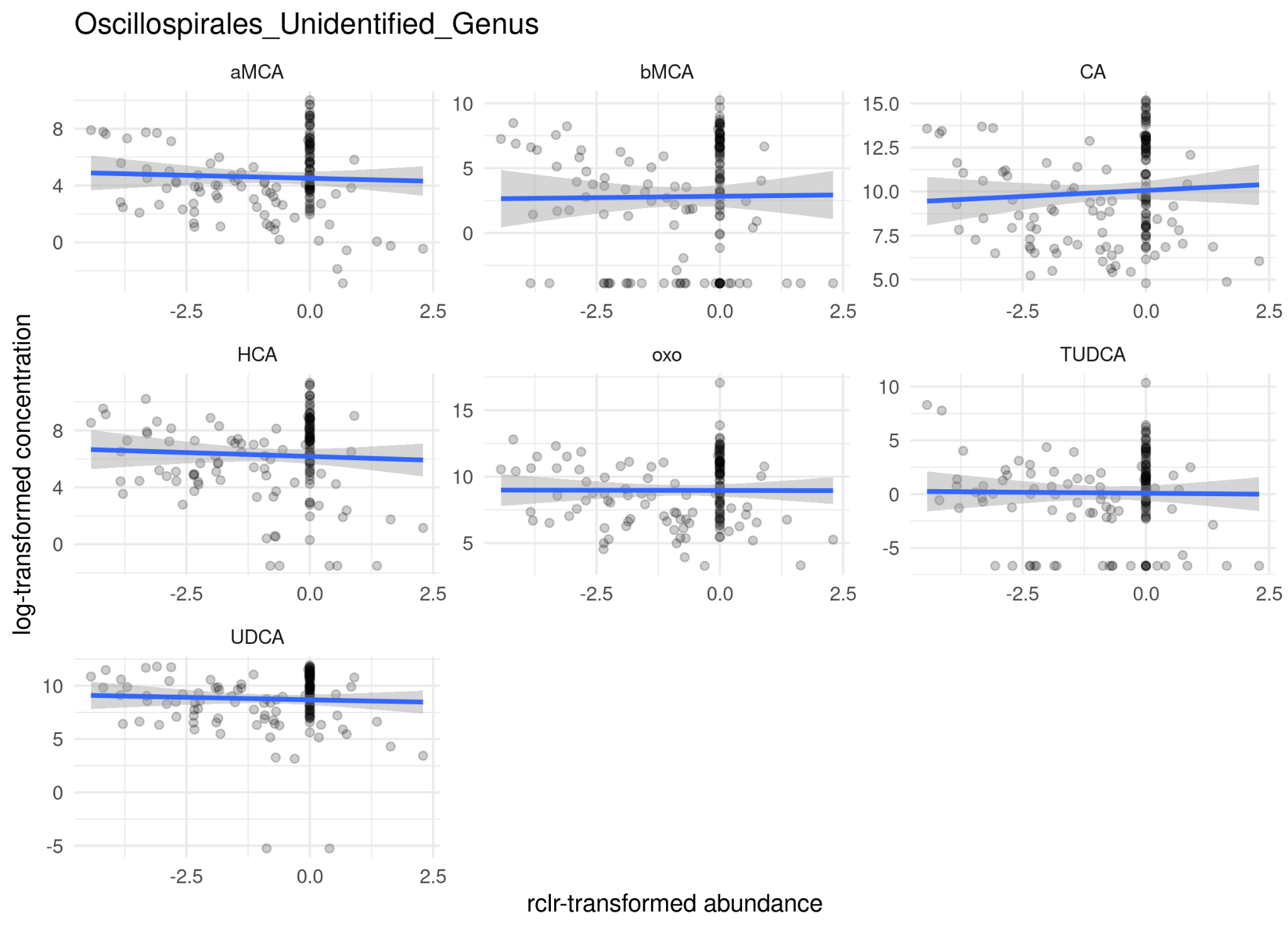

Figure 56. At 30 months unidentified genus in Oscillospirales order associated negative with bile acids, including 7-oxo-converted bile acids. Bile acid data is log-transformed whereas abundance data is robust centered log-transformed (rclr) for the visualization.

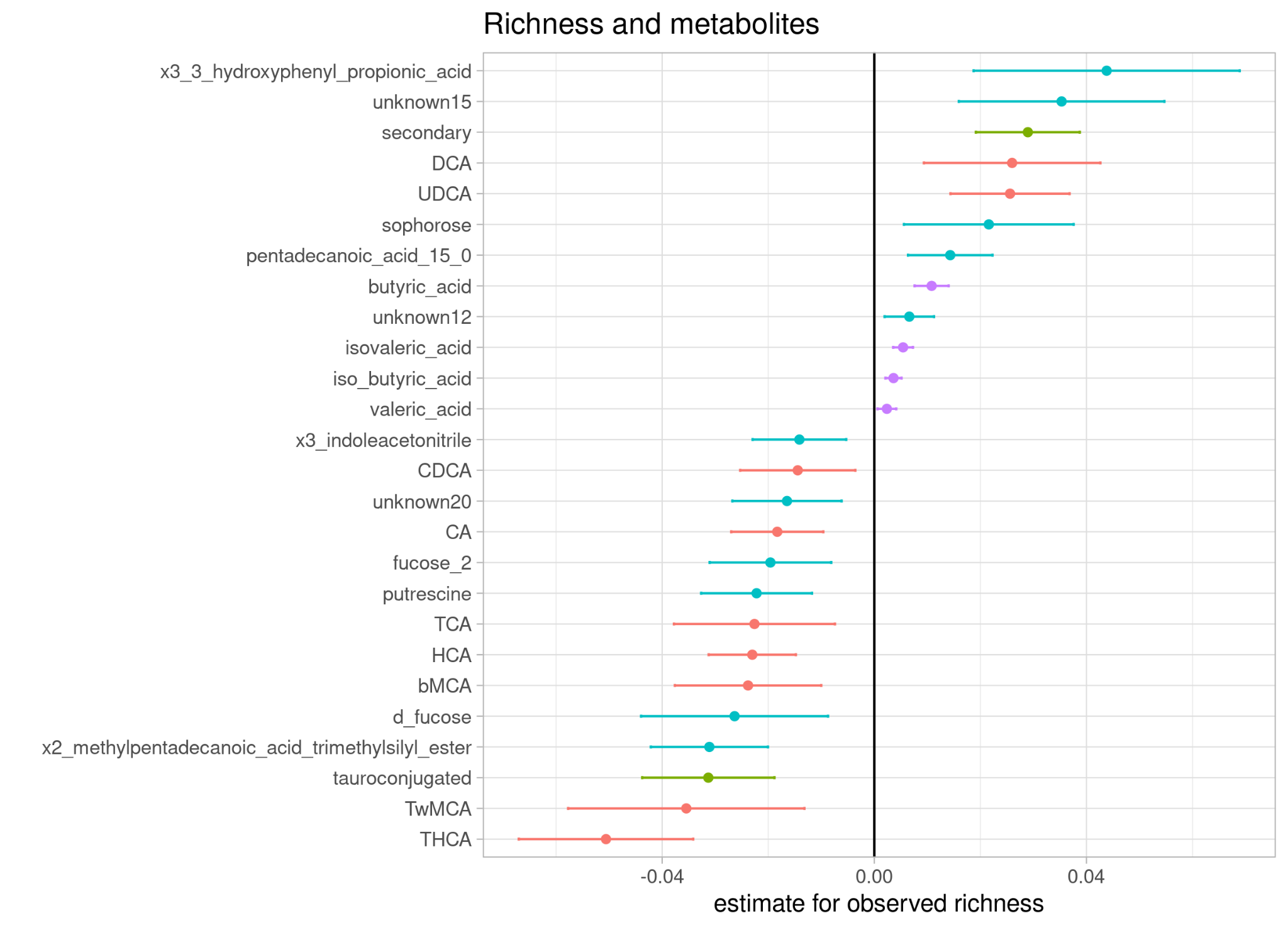

Figure 57. Estimates for richness in mixed-model adjusted for age.

### Clusters have different levels of metabolites

Table 8. Metabolite concentration differences between clusters with Kruskall-Wallis test and Dunn’s Posthoc test. * Adjusted p-value from Dunn’s posthoc test ** adjusted p-value from Kruskall-Wallis test. Only results with q < 0.05 and Dunn’s test adjusted p-value < 0.05 are shown.

| assay | rowname | timepoint | group1 | group2 | n1 | n2 | effsize | magnitude | p_adjusted_dunns | q |
| --- | --- | --- | --- | --- | --- | --- | --- | --- | --- | --- |
| BileAcids | TwMCA | 2.5 | C1 | C2 | 60 | 90 | 0.19 | large | 0 | 5.16E-10 |
| BileAcids | TwMCA | 2.5 | C1 | C3 | 60 | 86 | 0.19 | large | 0 | 5.16E-10 |
| PolarMetabolites | unknown10 | 2.5 | C1 | C2 | 59 | 91 | 0.18 | large | 0 | 1.08E-09 |
| PolarMetabolites | unknown10 | 2.5 | C1 | C3 | 59 | 87 | 0.18 | large | 0 | 1.08E-09 |
| BileAcids | GCA | 2.5 | C1 | C2 | 60 | 90 | 0.17 | large | 0 | 4.91E-09 |
| BileAcids | GCA | 2.5 | C1 | C3 | 60 | 86 | 0.17 | large | 0 | 4.91E-09 |
| PolarMetabolites | succinic_acid | 2.5 | C1 | C3 | 59 | 87 | 0.16 | large | 0 | 1.27E-08 |
| PolarMetabolites | succinic_acid | 2.5 | C2 | C3 | 91 | 87 | 0.16 | large | 2.00E-05 | 1.27E-08 |
| BileAcids | THCA | 2.5 | C1 | C2 | 60 | 90 | 0.14 | large | 0 | 5.59E-08 |
| BileAcids | THCA | 2.5 | C1 | C3 | 60 | 86 | 0.14 | large | 0 | 5.59E-08 |
| BileAcids | TCA | 2.5 | C1 | C2 | 60 | 90 | 0.14 | large | 0 | 8.47E-08 |
| BileAcids | TCA | 2.5 | C1 | C3 | 60 | 86 | 0.14 | large | 0 | 8.47E-08 |
| SCFA | butyric_acid | 14 | C4 | C6 | 4 | 149 | 0.12 | moderate | 0.01935 | 1.42E-07 |
| SCFA | butyric_acid | 14 | C5 | C6 | 90 | 149 | 0.12 | moderate | 0 | 1.42E-07 |
| BileAcids | GCDCA | 2.5 | C1 | C2 | 60 | 90 | 0.11 | moderate | 1.00E-05 | 1.76E-06 |
| BileAcids | GCDCA | 2.5 | C1 | C3 | 60 | 86 | 0.11 | moderate | 1.00E-05 | 1.76E-06 |
| PolarMetabolites | x9_octadecen_1_ol | 2.5 | C1 | C2 | 59 | 91 | 0.11 | moderate | 0 | 3.66E-06 |
| PolarMetabolites | x9_octadecen_1_ol | 2.5 | C1 | C3 | 59 | 87 | 0.11 | moderate | 0.00247 | 3.66E-06 |
| PolarMetabolites | unknown11 | 2.5 | C1 | C2 | 59 | 91 | 0.10 | moderate | 0 | 9.01E-06 |
| PolarMetabolites | unknown11 | 2.5 | C1 | C3 | 59 | 87 | 0.10 | moderate | 0.00574 | 9.01E-06 |
| PolarMetabolites | shikimic_acid | 2.5 | C2 | C3 | 91 | 87 | 0.09 | moderate | 1.00E-05 | 1.40E-05 |
| PolarMetabolites | pentadecanoic_acid_15_0 | 2.5 | C1 | C2 | 59 | 91 | 0.09 | moderate | 7.70E-04 | 1.89E-05 |
| PolarMetabolites | pentadecanoic_acid_15_0 | 2.5 | C1 | C3 | 59 | 87 | 0.09 | moderate | 3.00E-05 | 1.89E-05 |
| PolarMetabolites | x2_methylpentadecanoic_acid_trimethylsilyl_ester | 2.5 | C1 | C3 | 59 | 87 | 0.09 | moderate | 9.00E-05 | 2.50E-05 |
| PolarMetabolites | x2_methylpentadecanoic_acid_trimethylsilyl_ester | 2.5 | C2 | C3 | 91 | 87 | 0.09 | moderate | 6.80E-04 | 2.50E-05 |
| PolarMetabolites | unknown8 | 2.5 | C1 | C2 | 59 | 91 | 0.07 | moderate | 0.00461 | 1.33E-04 |
| PolarMetabolites | unknown8 | 2.5 | C2 | C3 | 91 | 87 | 0.07 | moderate | 4.80E-04 | 1.33E-04 |
| BileAcids | TbMCA | 2.5 | C1 | C2 | 60 | 90 | 0.07 | moderate | 0.00628 | 2.34E-04 |
| BileAcids | TbMCA | 2.5 | C1 | C3 | 60 | 86 | 0.07 | moderate | 3.10E-04 | 2.34E-04 |
| PolarMetabolites | asparagine | 2.5 | C1 | C2 | 59 | 91 | 0.07 | moderate | 0.02141 | 3.76E-04 |
| PolarMetabolites | asparagine | 2.5 | C2 | C3 | 91 | 87 | 0.07 | moderate | 6.70E-04 | 3.76E-04 |
| PolarMetabolites | hexose | 2.5 | C1 | C3 | 59 | 87 | 0.06 | moderate | 0.00178 | 4.05E-04 |
| PolarMetabolites | hexose | 2.5 | C2 | C3 | 91 | 87 | 0.06 | moderate | 0.00141 | 4.05E-04 |
| PolarMetabolites | arachidonic_acid | 2.5 | C1 | C2 | 59 | 91 | 0.06 | moderate | 3.40E-04 | 5.19E-04 |
| PolarMetabolites | arachidonic_acid | 2.5 | C1 | C3 | 59 | 87 | 0.06 | moderate | 0.00646 | 5.19E-04 |
| PolarMetabolites | x3_phosphoglycerate | 2.5 | C2 | C3 | 91 | 87 | 0.06 | moderate | 4.30E-04 | 5.85E-04 |
| PolarMetabolites | succinic_acid | 30 | C4 | C6 | 1 | 15 | 0.07 | moderate | 0.04446 | 0.00307 |
| PolarMetabolites | succinic_acid | 30 | C6 | C7 | 15 | 130 | 0.07 | moderate | 0.00842 | 0.00307 |
| SCFA | valeric_acid | 30 | C6 | C7 | 15 | 129 | 0.07 | moderate | 0.00252 | 0.00319 |
| PolarMetabolites | unknown15 | 30 | C6 | C7 | 15 | 130 | 0.06 | moderate | 0.00333 | 0.00371 |
| BileAcids | bMCA | 30 | C6 | C7 | 15 | 129 | 0.06 | moderate | 0.00657 | 0.00382 |
| PolarMetabolites | d_trehalose | 14 | C5 | C6 | 88 | 150 | 0.06 | small | 0.0011 | 4.58E-04 |
| BileAcids | UDCA | 2.5 | C1 | C2 | 60 | 90 | 0.06 | small | 0.04861 | 7.88E-04 |
| BileAcids | UDCA | 2.5 | C1 | C3 | 60 | 86 | 0.06 | small | 9.70E-04 | 7.88E-04 |
| BileAcids | 7-oxo-DCA | 2.5 | C1 | C3 | 60 | 86 | 0.06 | small | 0.00314 | 0.00113 |
| BileAcids | 7-oxo-DCA | 2.5 | C2 | C3 | 90 | 86 | 0.06 | small | 0.00858 | 0.00113 |
| SCFA | propionic_acid | 2.5 | C1 | C2 | 60 | 91 | 0.05 | small | 0.00111 | 0.00131 |
| PolarMetabolites | unknown24 | 2.5 | C1 | C2 | 59 | 91 | 0.05 | small | 0.0027 | 0.00195 |
| PolarMetabolites | unknown24 | 2.5 | C1 | C3 | 59 | 87 | 0.05 | small | 0.02119 | 0.00195 |
| PolarMetabolites | pentadecanoic_acid_15_0 | 14 | C5 | C6 | 88 | 150 | 0.04 | small | 0.00253 | 0.00212 |
| PolarMetabolites | unknown20 | 2.5 | C1 | C2 | 59 | 91 | 0.05 | small | 0.00265 | 0.00326 |
| PolarMetabolites | unknown20 | 2.5 | C1 | C3 | 59 | 87 | 0.05 | small | 0.01741 | 0.00326 |
| PolarMetabolites | d_fructose | 2.5 | C2 | C3 | 91 | 87 | 0.05 | small | 0.00923 | 0.00351 |
| PolarMetabolites | d_mannitol | 14 | C4 | C5 | 4 | 88 | 0.04 | small | 0.01007 | 0.00355 |
| PolarMetabolites | d_mannitol | 14 | C4 | C6 | 4 | 150 | 0.04 | small | 0.0325 | 0.00355 |
| PolarMetabolites | d_mannitol | 14 | C5 | C6 | 88 | 150 | 0.04 | small | 0.0358 | 0.00355 |
| PolarMetabolites | unknown6 | 2.5 | C1 | C2 | 59 | 91 | 0.04 | small | 0.01907 | 0.00423 |
| PolarMetabolites | unknown6 | 2.5 | C2 | C3 | 91 | 87 | 0.04 | small | 0.0193 | 0.00423 |
| BileAcids | 7-oxo-HDCA | 2.5 | C1 | C3 | 60 | 86 | 0.04 | small | 0.03775 | 0.00487 |
| BileAcids | 7-oxo-HDCA | 2.5 | C2 | C3 | 90 | 86 | 0.04 | small | 0.00799 | 0.00487 |
| PolarMetabolites | unknown16 | 2.5 | C1 | C3 | 59 | 87 | 0.04 | small | 0.00277 | 0.00527 |
| BileAcids | TwMCA | 6 | C2 | C4 | 7 | 181 | 0.05 | small | 0.00573 | 0.00528 |
| PolarMetabolites | unknown2 | 14 | C5 | C6 | 88 | 150 | 0.03 | small | 0.00515 | 0.00698 |
| SCFA | iso_butyric_acid | 2.5 | C1 | C2 | 60 | 91 | 0.04 | small | 0.00399 | 0.00725 |
| PolarMetabolites | galactonic_acid | 2.5 | C1 | C3 | 59 | 87 | 0.04 | small | 0.01041 | 0.00763 |
| PolarMetabolites | galactonic_acid | 2.5 | C2 | C3 | 91 | 87 | 0.04 | small | 0.03262 | 0.00763 |
| PolarMetabolites | arachidonic_acid | 14 | C5 | C6 | 88 | 150 | 0.03 | small | 0.00644 | 0.00884 |
| BileAcids | 7-oxo-DCA | 30 | C6 | C7 | 15 | 129 | 0.05 | small | 0.01723 | 0.0093 |
| PolarMetabolites | allose1 | 2.5 | C1 | C3 | 59 | 87 | 0.04 | small | 0.00552 | 0.00944 |
| BileAcids | CA | 30 | C6 | C7 | 15 | 129 | 0.05 | small | 0.02345 | 0.00971 |
| PolarMetabolites | arachidonic_acid | 6 | C2 | C4 | 7 | 180 | 0.04 | small | 0.00606 | 0.0104 |
| PolarMetabolites | ribitol_2 | 2.5 | C1 | C2 | 59 | 91 | 0.04 | small | 0.01424 | 0.0106 |
| PolarMetabolites | x4_hydroxyphenylacetic_acid | 30 | C6 | C7 | 15 | 130 | 0.05 | small | 0.01656 | 0.0107 |
| PolarMetabolites | plamitic_acid_16_0 | 2.5 | C1 | C3 | 59 | 87 | 0.03 | small | 0.00896 | 0.0114 |
| PolarMetabolites | unknown12 | 14 | C5 | C6 | 88 | 150 | 0.03 | small | 0.00842 | 0.0115 |
| PolarMetabolites | unknown17 | 14 | C5 | C6 | 88 | 150 | 0.03 | small | 0.01509 | 0.0125 |
| BileAcids | wMCA | 2.5 | C1 | C3 | 60 | 86 | 0.03 | small | 0.02439 | 0.0134 |
| BileAcids | HCA | 30 | C6 | C7 | 15 | 129 | 0.05 | small | 0.01601 | 0.0134 |
| PolarMetabolites | unknown15 | 2.5 | C1 | C2 | 59 | 91 | 0.03 | small | 0.03976 | 0.0135 |
| PolarMetabolites | unknown15 | 2.5 | C1 | C3 | 59 | 87 | 0.03 | small | 0.03361 | 0.0135 |
| PolarMetabolites | unknown17 | 2.5 | C1 | C3 | 59 | 87 | 0.03 | small | 0.03836 | 0.0137 |
| BileAcids | TCA | 30 | C6 | C7 | 15 | 129 | 0.05 | small | 0.03485 | 0.0138 |
| PolarMetabolites | x1_2_3_4_5_6_hexatrimethylsilylinositol | 2.5 | C1 | C2 | 59 | 91 | 0.03 | small | 0.03569 | 0.014 |
| PolarMetabolites | x3_4_dihydroxyhydrocinnamic_acid | 30 | C6 | C7 | 15 | 130 | 0.04 | small | 0.02547 | 0.0148 |
| SCFA | isovaleric_acid | 2.5 | C1 | C2 | 60 | 91 | 0.03 | small | 0.0193 | 0.0165 |
| PolarMetabolites | d_mannitol | 30 | C6 | C7 | 15 | 130 | 0.04 | small | 0.02629 | 0.0166 |
| SCFA | iso_butyric_acid | 14 | C4 | C5 | 4 | 90 | 0.03 | small | 0.01443 | 0.0176 |
| SCFA | iso_butyric_acid | 14 | C4 | C6 | 4 | 149 | 0.03 | small | 0.01443 | 0.0176 |
| PolarMetabolites | d_ribose | 14 | C4 | C5 | 4 | 88 | 0.02 | small | 0.01556 | 0.0194 |
| PolarMetabolites | d_ribose | 14 | C4 | C6 | 4 | 150 | 0.02 | small | 0.0152 | 0.0194 |
| PolarMetabolites | sinapic_acid | 2.5 | C2 | C3 | 91 | 87 | 0.03 | small | 0.04039 | 0.0203 |
| BileAcids | TbMCA | 30 | C6 | C7 | 15 | 129 | 0.04 | small | 0.02015 | 0.0209 |
| PolarMetabolites | minoxidil | 2.5 | C1 | C2 | 59 | 91 | 0.03 | small | 0.04502 | 0.0234 |
| PolarMetabolites | fucose_2 | 2.5 | C1 | C2 | 59 | 91 | 0.03 | small | 0.02504 | 0.0235 |
| PolarMetabolites | putrescine | 6 | C2 | C4 | 7 | 180 | 0.03 | small | 0.01644 | 0.0259 |
| PolarMetabolites | minoxidil | 14 | C5 | C6 | 88 | 150 | 0.02 | small | 0.02699 | 0.0272 |
| PolarMetabolites | d_fucose | 6 | C2 | C3 | 7 | 5 | 0.03 | small | 0.04157 | 0.0281 |
| PolarMetabolites | d_fucose | 6 | C2 | C4 | 7 | 180 | 0.03 | small | 0.04157 | 0.0281 |
| PolarMetabolites | hexose | 14 | C4 | C5 | 4 | 88 | 0.02 | small | 0.0311 | 0.0311 |
| PolarMetabolites | hexose | 14 | C4 | C6 | 4 | 150 | 0.02 | small | 0.04122 | 0.0311 |
| PolarMetabolites | l_iditol | 30 | C6 | C7 | 15 | 130 | 0.03 | small | 0.03287 | 0.0313 |
| PolarMetabolites | unknown24 | 14 | C5 | C6 | 88 | 150 | 0.02 | small | 0.04322 | 0.0325 |
| PolarMetabolites | hexose | 30 | C6 | C7 | 15 | 130 | 0.03 | small | 0.0359 | 0.0349 |
| BileAcids | UDCA | 30 | C6 | C7 | 15 | 129 | 0.03 | small | 0.04416 | 0.0374 |
| PolarMetabolites | unknown20 | 14 | C5 | C6 | 88 | 150 | 0.02 | small | 0.03345 | 0.0382 |
| PolarMetabolites | minoxidil | 6 | C2 | C3 | 7 | 5 | 0.03 | small | 0.04451 | 0.0385 |
| PolarMetabolites | ribitol | 6 | C2 | C4 | 7 | 180 | 0.03 | small | 0.02531 | 0.041 |
| BileAcids | TUDCA | 2.5 | C1 | C3 | 60 | 86 | 0.02 | small | 0.04839 | 0.041 |
| BileAcids | 7-oxo-HDCA | 30 | C6 | C7 | 15 | 129 | 0.03 | small | 0.04049 | 0.044 |
| SCFA | butyric_acid | 30 | C6 | C7 | 15 | 129 | 0.03 | small | 0.04548 | 0.0467 |

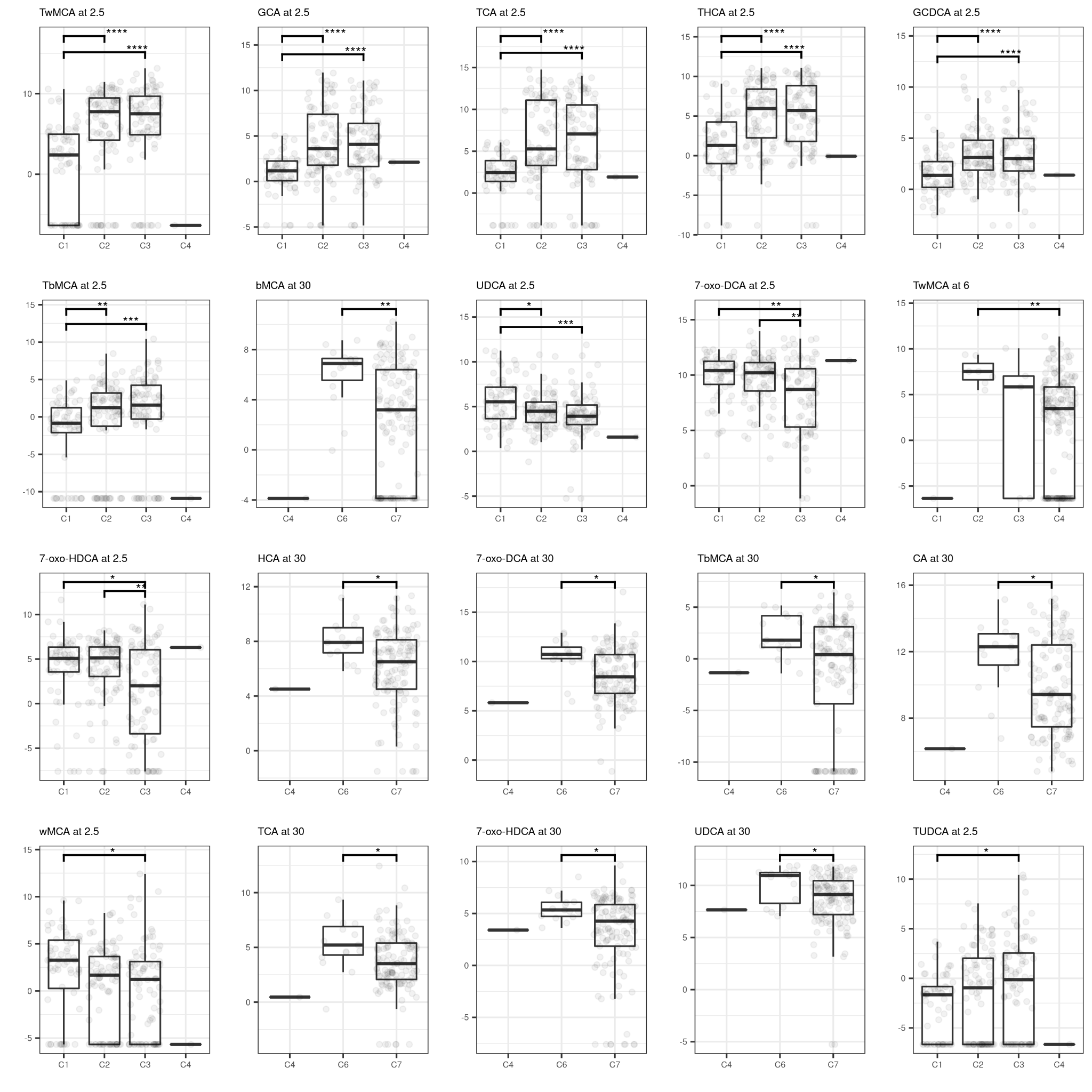

Figure 58. Differences in bile acid concentrations between clusters per timepoint. Only results with q < 0.05 and Dunn’s test adjusted p-value < 0.05 are shown.

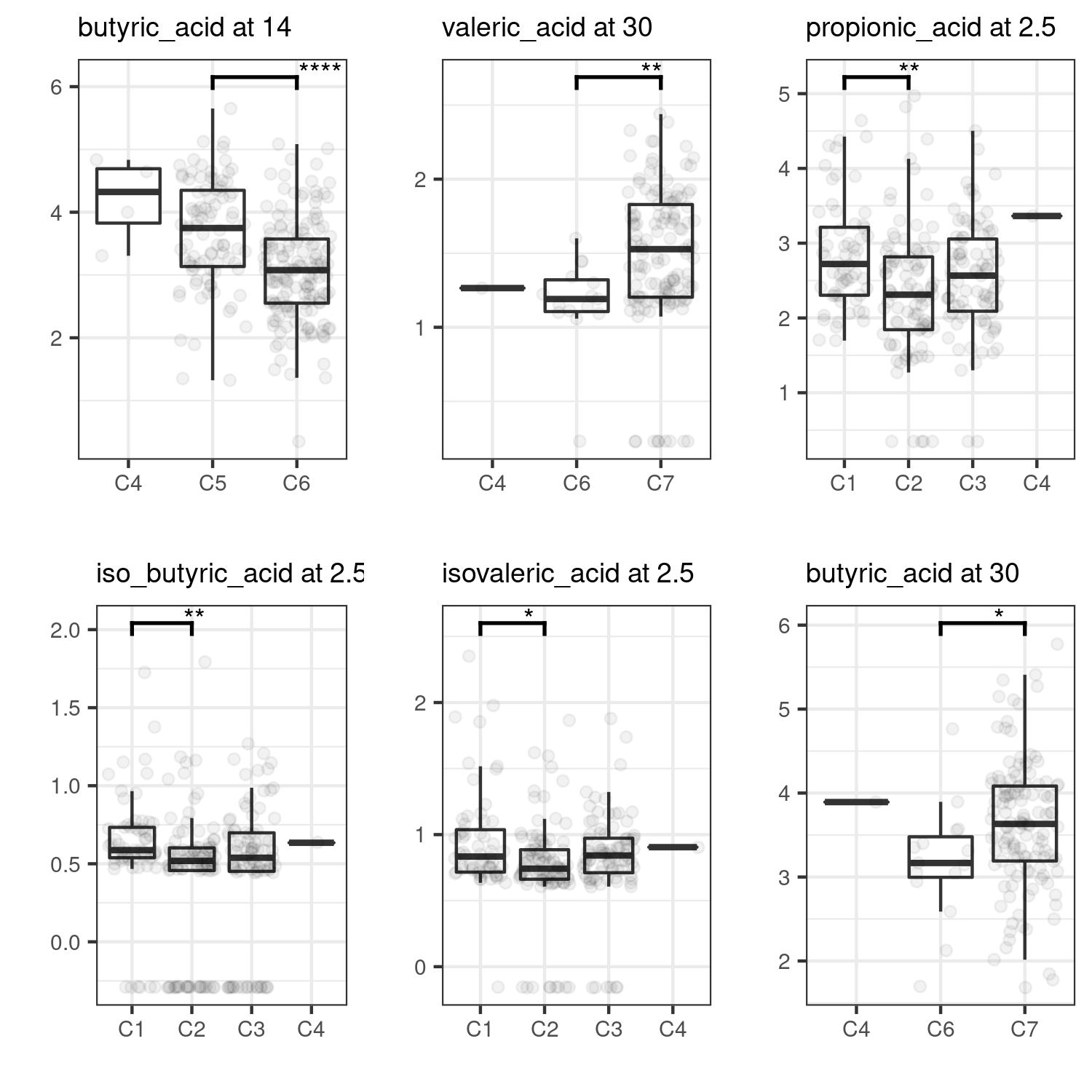

Figure 59. Differences in SCFA concentrations between clusters per timepoint. Only results with q < 0.05 and Dunn’s test adjusted p-value < 0.05 are shown.

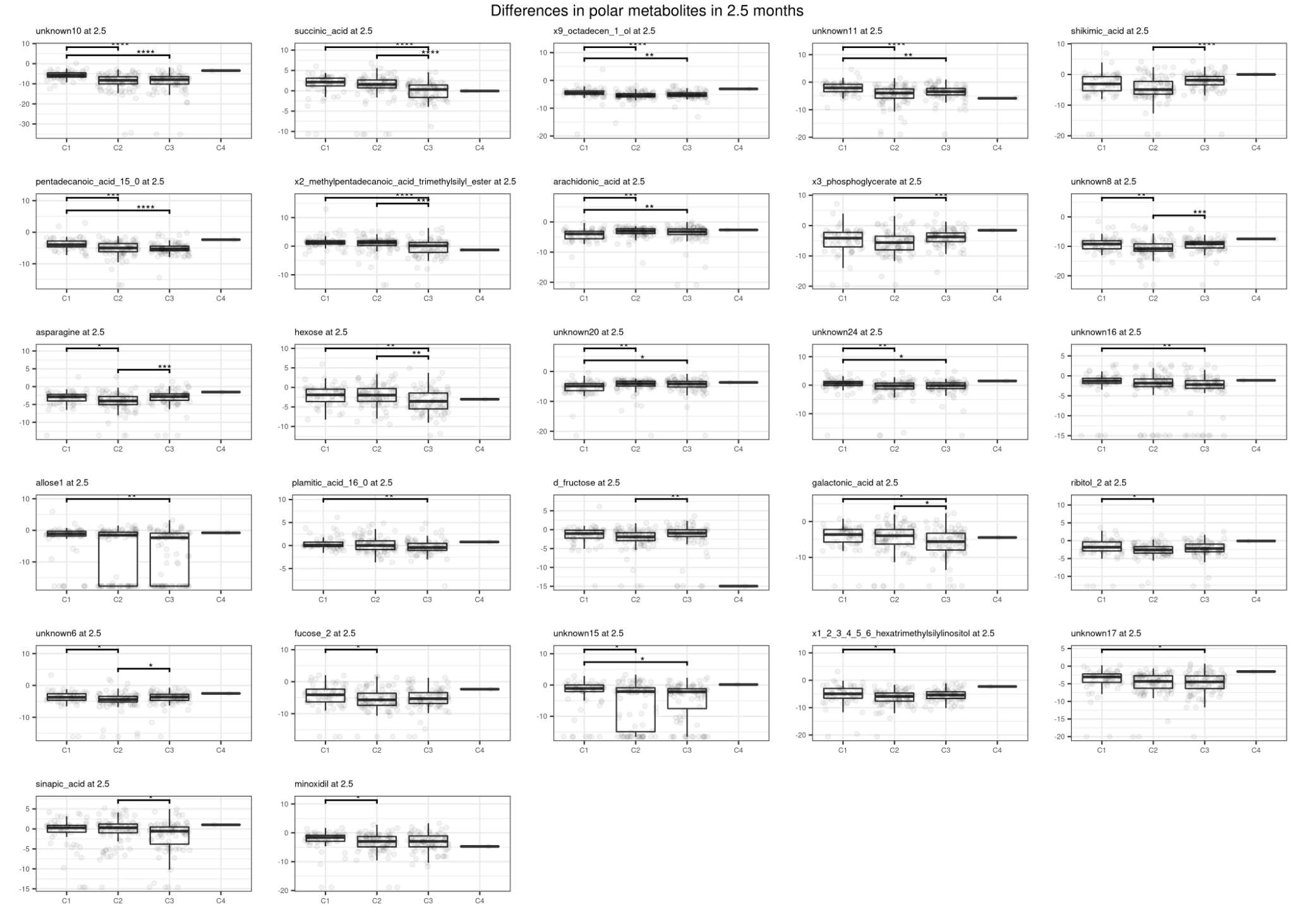

Figure 60. Differences in polar metabolites concentrations between clusters at 2.5 months. Only results with q < 0.05 and Dunn’s test adjusted p-value < 0.05 are shown.

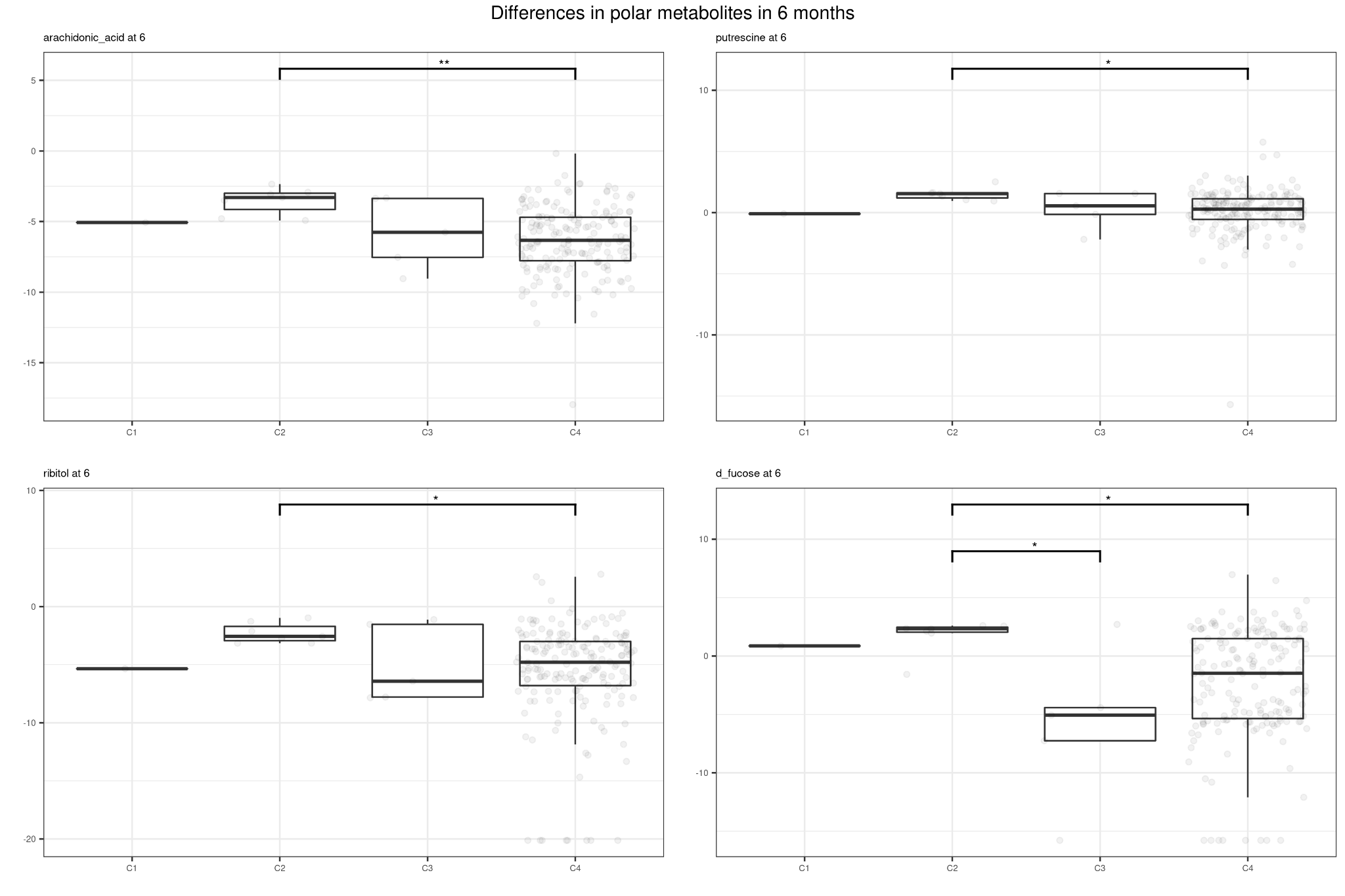

Figure 61. Differences in polar metabolites concentrations between clusters at 6 months. Only results with q < 0.05 and Dunn’s test adjusted p-value < 0.05 are shown.

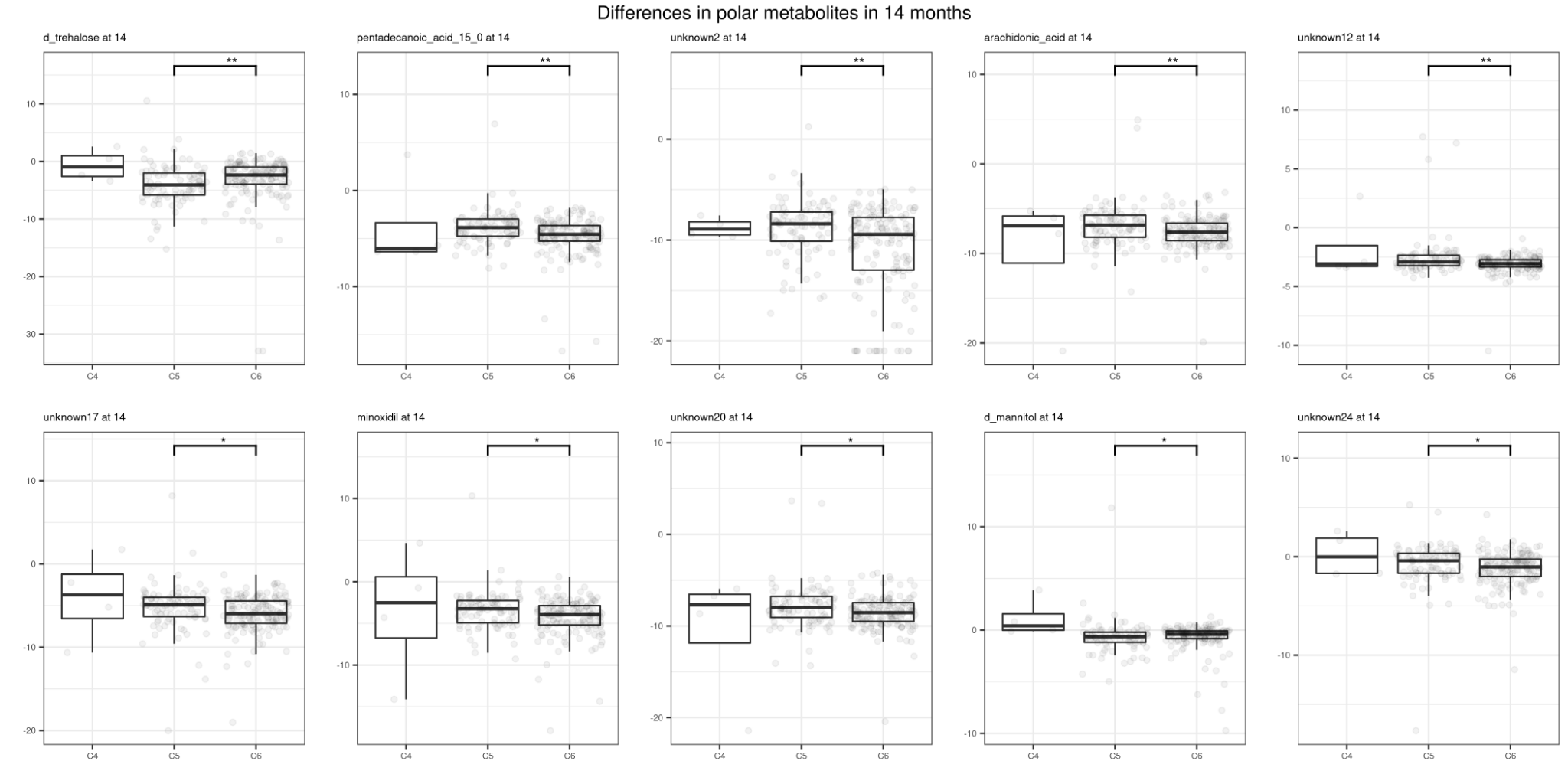

Figure 62. Differences in polar metabolites concentrations between clusters at 14 months. Only results with q < 0.05 and Dunn’s test adjusted p-value < 0.05 are shown.

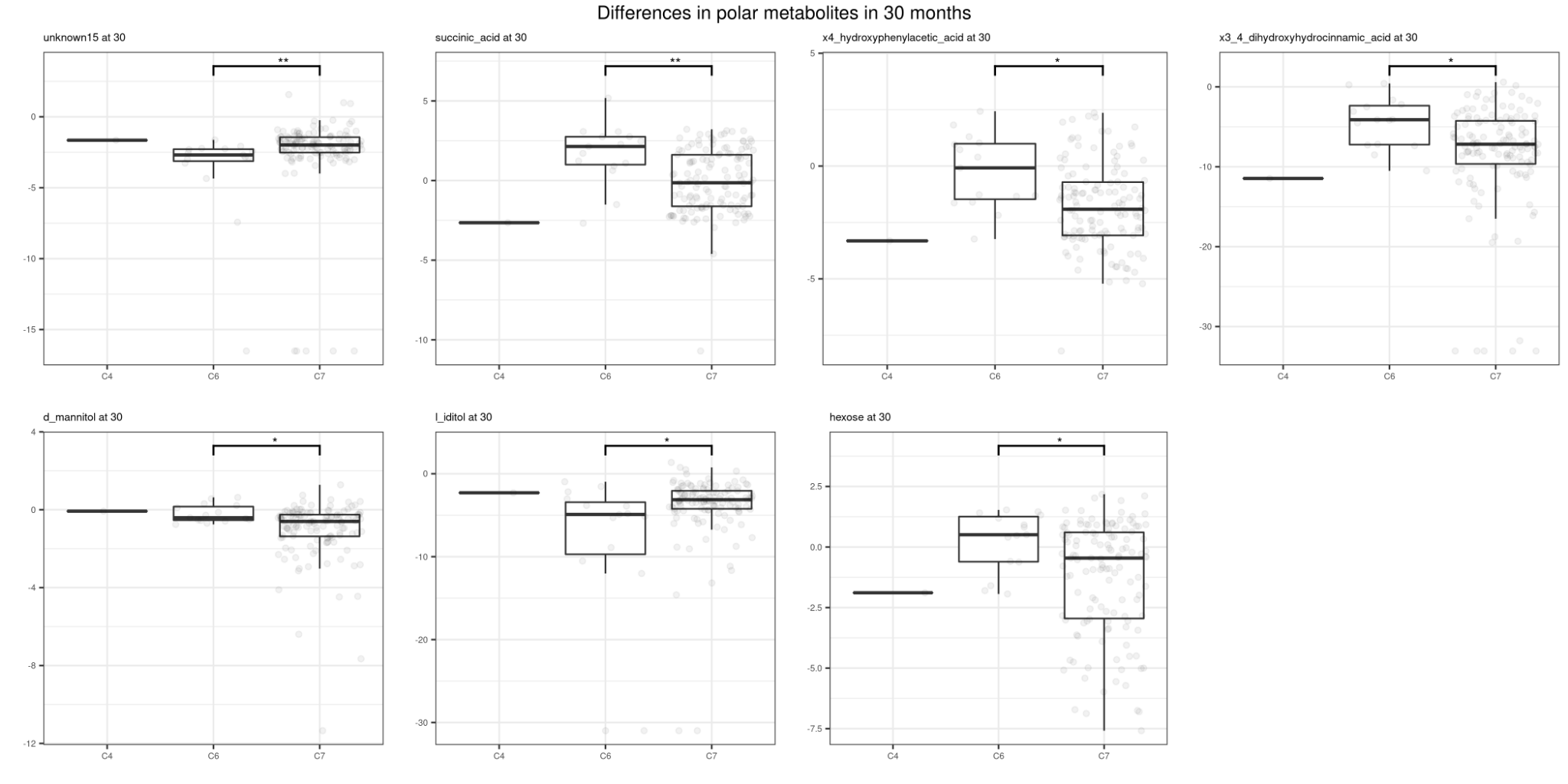

Figure 63. Differences in polar metabolites concentrations between clusters at 30 months. Only results with q < 0.05 and Dunn’s test adjusted p-value < 0.05 are shown.

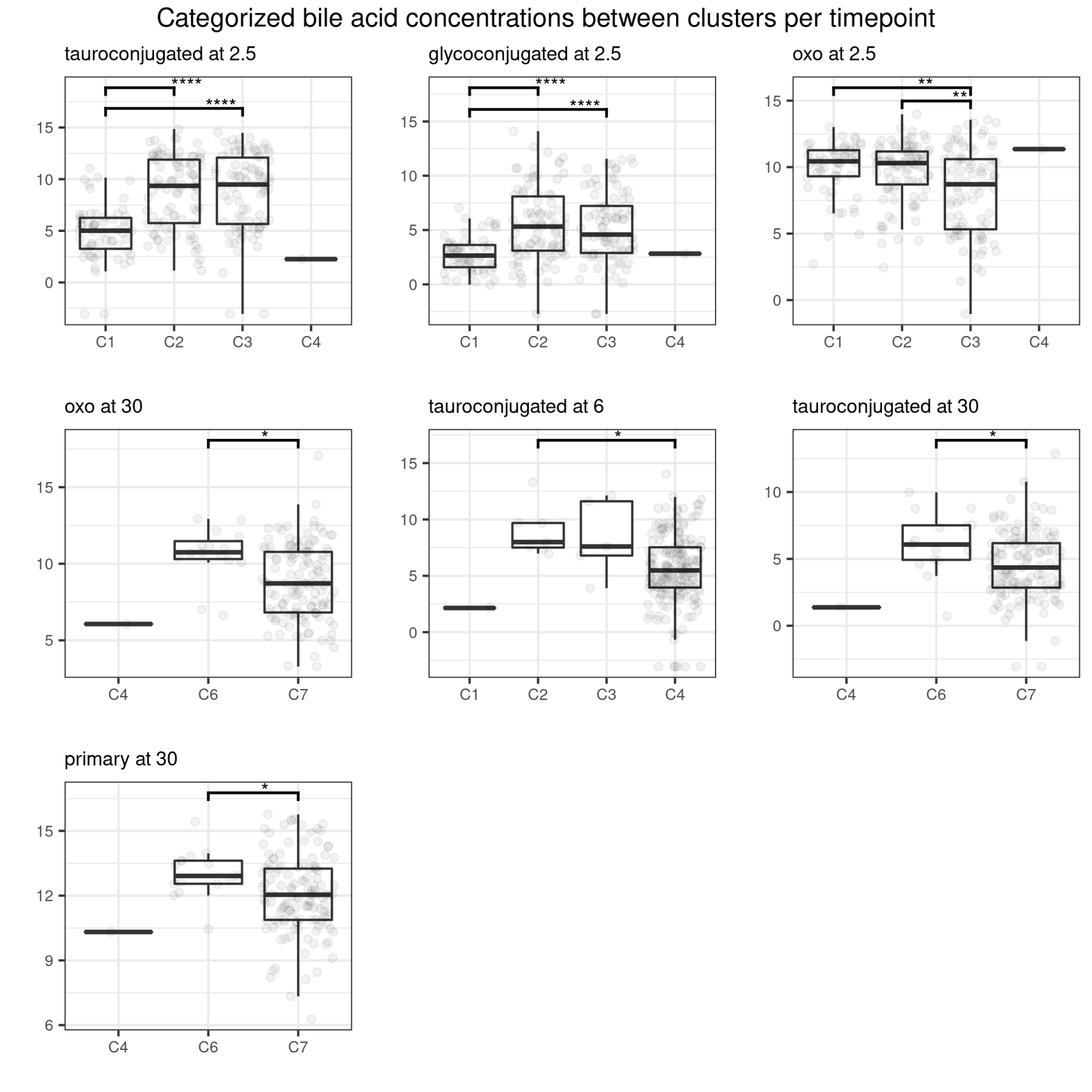

Figure 64. Differences in categorized bile acid concentrations between clusters. Only results with q < 0.05 and Dunn’s test adjusted p-value < 0.05 are shown.

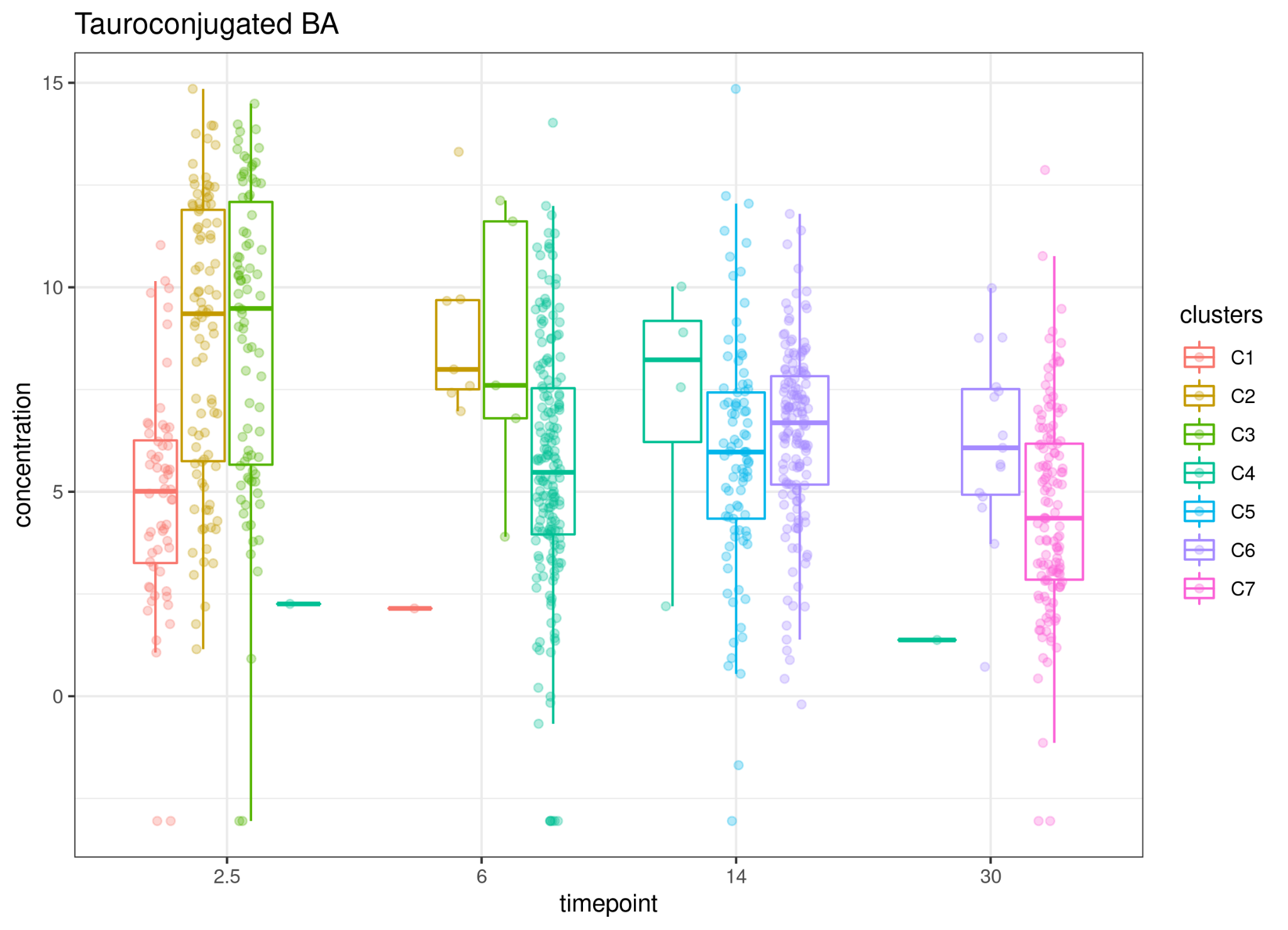

Figure 65. Tauroconjugated bile acids were lower in C1 compared with C2, C3 and C6 in mixed-models.

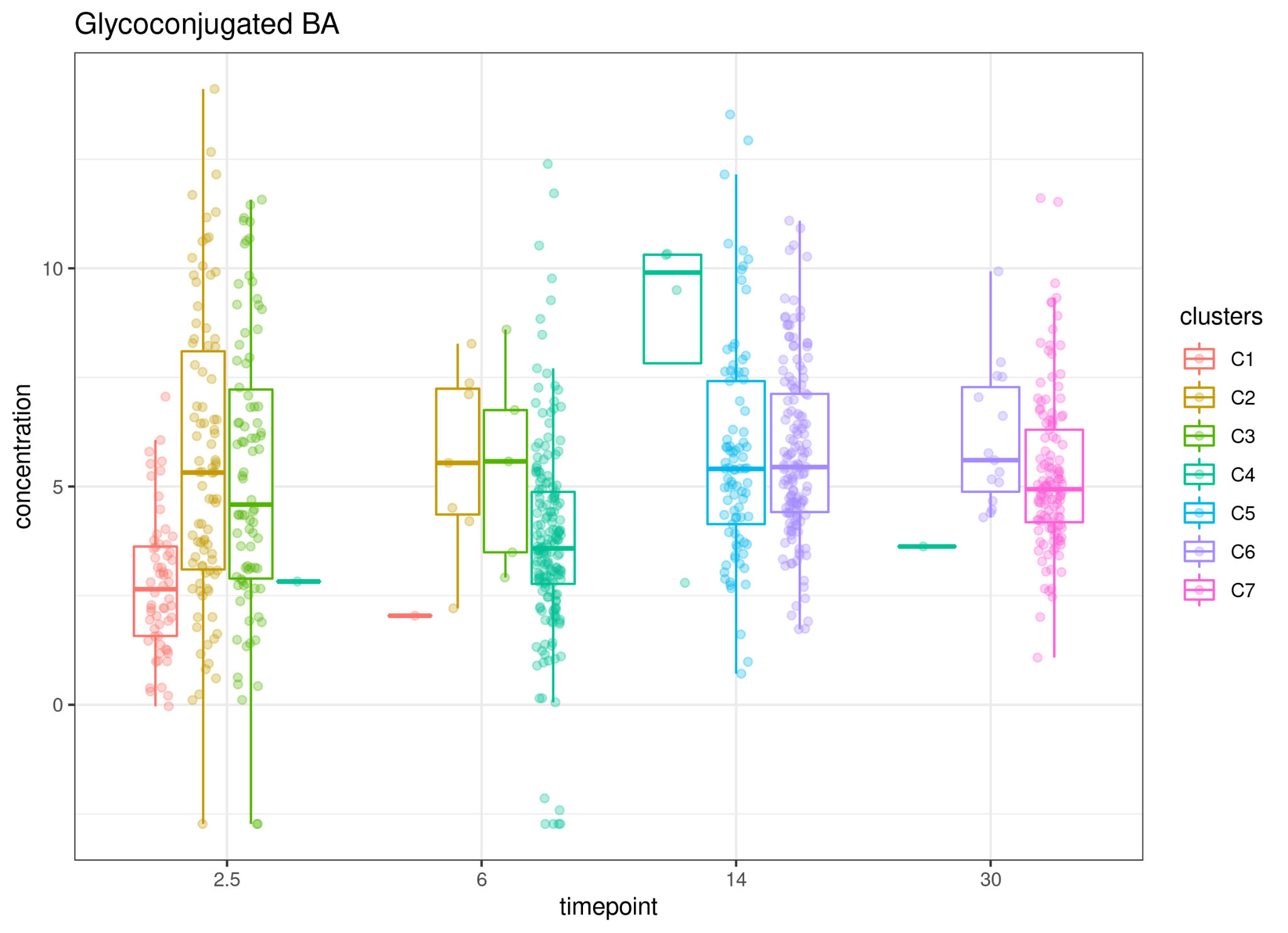

Figure 66. Glycoonjugated bile acids were lower in C1 compared with C2, C3, C4, C5 and C6 in mixed-models.

### Breastfeeding interaction with prevalent taxa abundances

Supplementary Table 9. Breastfeeding interaction with prevalent taxa rclr-transformed abundances in mixed model.

| taxa | rowname | statistic | p.value | lower | est. | upper | q | significant |
| --- | --- | --- | --- | --- | --- | --- | --- | --- |
| Bifidobacterium | GCA | -7.3 | 0.00 | -1.002 | -0.788 | -0.575 | 0.00000 | sig |
| Bifidobacterium | TCA | -5.8 | 0.00 | -1.024 | -0.763 | -0.502 | 0.00000 | sig |
| Bifidobacterium | tauroconjugated | -4.7 | 0.00 | -0.735 | -0.516 | -0.298 | 0.00060 | sig |
| Bifidobacterium | GUDCA | -4.1 | 0.00 | -0.936 | -0.634 | -0.331 | 0.00433 | sig |
| Bacteroides | pinitol | 3.8 | 0.00 | 0.268 | 0.554 | 0.840 | 0.00978 | sig |
| Escherichia | 7-oxo-HDCA | 3.8 | 0.00 | 0.224 | 0.463 | 0.702 | 0.00978 | sig |
| Bifidobacterium | aMCA | 3.3 | 0.00 | 0.114 | 0.280 | 0.447 | 0.04788 | sig |
| Bacteroides | secondary | 3.3 | 0.00 | 0.119 | 0.299 | 0.478 | 0.04788 | sig |
| Bacteroides | oxo | 3.3 | 0.00 | 0.115 | 0.290 | 0.464 | 0.04788 | sig |
| Bacteroides | unknown15 | 3.2 | 0.00 | 0.226 | 0.601 | 0.976 | 0.06074 | sig |
| Bacteroides | 7-oxo-DCA | 3.1 | 0.00 | 0.108 | 0.290 | 0.472 | 0.06074 | sig |
| Bifidobacterium | Acetic acid | 3.0 | 0.00 | 0.027 | 0.077 | 0.127 | 0.07961 | sig |
| Bifidobacterium | x3_indoleacetonitrile | -2.9 | 0.00 | -0.413 | -0.247 | -0.081 | 0.09156 | sig |
| Escherichia | oxo | 2.9 | 0.00 | 0.071 | 0.217 | 0.362 | 0.09156 | sig |
| Veillonella | x3_4_dihydroxyhydrocinnamic_acid | 2.8 | 0.01 | 0.200 | 0.669 | 1.138 | 0.11490 | sig |
| Clostridiaceae_Clostridium | succinic_acid | -2.8 | 0.01 | -0.569 | -0.334 | -0.100 | 0.11490 | sig |
| Bacteroides | tauroconjugated | -2.6 | 0.01 | -0.555 | -0.318 | -0.081 | 0.17749 | sig |
| Bifidobacterium | bMCA | 2.6 | 0.01 | 0.078 | 0.336 | 0.593 | 0.19484 | sig |
| Bifidobacterium | HCA | 2.6 | 0.01 | 0.047 | 0.199 | 0.352 | 0.19484 | sig |
| Veillonella | x5_hydroxyindoleactate_1 | 2.5 | 0.01 | 0.089 | 0.408 | 0.728 | 0.20353 | sig |
| Escherichia | GUDCA | 2.5 | 0.01 | 0.074 | 0.338 | 0.602 | 0.20353 | sig |
| Bacteroides | TwMCA | -2.5 | 0.01 | -0.961 | -0.536 | -0.110 | 0.20797 | sig |
| Bacteroides | 7-oxo-HDCA | 2.5 | 0.01 | 0.076 | 0.370 | 0.664 | 0.20797 | sig |
| Bifidobacterium | wMCA | 2.5 | 0.01 | 0.085 | 0.432 | 0.779 | 0.21162 | sig |
| Bacteroides | THCA | -2.4 | 0.02 | -0.706 | -0.390 | -0.074 | 0.21544 | sig |
| Escherichia | 7-oxo-DCA | 2.4 | 0.02 | 0.035 | 0.186 | 0.338 | 0.21544 | sig |
| Bacteroides | bMCA | 2.4 | 0.02 | 0.055 | 0.322 | 0.588 | 0.23194 | sig |
| Bacteroides | c16 | 2.4 | 0.02 | 0.068 | 0.411 | 0.753 | 0.23224 | sig |
| Bifidobacterium | x1_2_3_4_5_6_hexatrimethylsilylinositol | 2.3 | 0.02 | 0.035 | 0.224 | 0.413 | 0.23617 | sig |
| Bifidobacterium | unknown20 | -2.3 | 0.02 | -0.408 | -0.221 | -0.034 | 0.23617 | sig |
| Bifidobacterium | galactonic_acid | -2.2 | 0.03 | -0.579 | -0.307 | -0.035 | 0.29701 | sig |
| Bifidobacterium | THCA | -2.2 | 0.03 | -0.637 | -0.337 | -0.037 | 0.29701 | sig |
| Bifidobacterium | unknown3 | 2.2 | 0.03 | 0.015 | 0.145 | 0.276 | 0.29818 | sig |
| Bacteroides | x3_indoleacetonitrile | 2.2 | 0.03 | 0.019 | 0.192 | 0.366 | 0.29818 | sig |
| Clostridiaceae_Clostridium | putrescine | 2.2 | 0.03 | 0.024 | 0.251 | 0.478 | 0.29818 | sig |
| Bifidobacterium | palmitoleic_acid | -2.1 | 0.03 | -0.325 | -0.169 | -0.014 | 0.31259 | sig |
| Veillonella | x7_hexadecenyl_acetate | -2.1 | 0.04 | -0.483 | -0.249 | -0.015 | 0.34506 | sig |
| Bacteroides | x4_hydroxyphenylacetic_acid | 2.1 | 0.04 | 0.014 | 0.259 | 0.504 | 0.34769 | sig |
| Clostridiaceae_Clostridium | x1_trimethylsilylmethyl_3_cyclohexenyl_5_methyl_4_hexen_1_ol | -2.1 | 0.04 | -0.338 | -0.173 | -0.007 | 0.35999 | sig |
| Bacteroides | pantothenic_acid | 2.0 | 0.04 | 0.007 | 0.212 | 0.417 | 0.36431 | sig |
| Bifidobacterium | x7_hexadecenyl_acetate | 2.0 | 0.04 | 0.007 | 0.232 | 0.457 | 0.36720 | sig |
| Bacteroides | x1_2_3_4_5_6_hexatrimethylsilylinositol | 2.0 | 0.04 | 0.005 | 0.201 | 0.398 | 0.36818 | sig |
| Bifidobacterium | d_ribose | 1.9 | 0.06 | -0.010 | 0.174 | 0.359 | 0.38730 | non.sig |
| Bifidobacterium | unknown10 | -1.9 | 0.05 | -0.725 | -0.359 | 0.007 | 0.38730 | non.sig |
| Bifidobacterium | pantothenic_acid | -1.9 | 0.05 | -0.394 | -0.195 | 0.003 | 0.38730 | non.sig |
| Veillonella | galactonic_acid | -1.9 | 0.06 | -0.545 | -0.266 | 0.013 | 0.38730 | non.sig |
| Bacteroides | palmitoleic_acid | -1.8 | 0.07 | -0.311 | -0.150 | 0.011 | 0.38730 | non.sig |
| Bacteroides | n_acetyl_d_glucosamine | 1.9 | 0.06 | -0.009 | 0.206 | 0.421 | 0.38730 | non.sig |
| Bacteroides | unknown20 | -1.9 | 0.06 | -0.380 | -0.186 | 0.007 | 0.38730 | non.sig |
| Clostridiaceae_Clostridium | x4_hydroxyphenylacetic_acid | 2.0 | 0.05 | -0.002 | 0.271 | 0.544 | 0.38730 | non.sig |
| Clostridiaceae_Clostridium | x3_indoleacetonitrile | 2.0 | 0.05 | 0.001 | 0.192 | 0.384 | 0.38730 | sig |
| Clostridiaceae_Clostridium | x1_2_3_4_5_6_hexatrimethylsilylinositol | 1.9 | 0.05 | -0.003 | 0.215 | 0.432 | 0.38730 | non.sig |
| Clostridiaceae_Clostridium | x5_hydroxyindoleactate_1 | 1.9 | 0.06 | -0.017 | 0.338 | 0.693 | 0.38730 | non.sig |
| Bifidobacterium | TwMCA | -1.9 | 0.06 | -0.802 | -0.390 | 0.022 | 0.38730 | non.sig |
| Bifidobacterium | primary | 1.9 | 0.07 | -0.007 | 0.113 | 0.232 | 0.38730 | non.sig |
| Bacteroides | TCA | -1.9 | 0.06 | -0.569 | -0.280 | 0.008 | 0.38730 | non.sig |
| Bacteroides | aMCA | 1.8 | 0.07 | -0.012 | 0.163 | 0.337 | 0.38730 | non.sig |
| Clostridiaceae_Clostridium | Propionic acid | 2.0 | 0.05 | 0.000 | 0.068 | 0.136 | 0.38730 | non.sig |
| Clostridiaceae_Clostridium | secondary | -1.8 | 0.07 | -0.390 | -0.189 | 0.013 | 0.38730 | non.sig |
| Escherichia | TCA | 1.8 | 0.07 | -0.016 | 0.223 | 0.462 | 0.38730 | non.sig |
| Bifidobacterium | x9_octadecen_1_ol | 1.8 | 0.07 | -0.009 | 0.104 | 0.217 | 0.38898 | non.sig |
| Veillonella | TUDCA | 1.8 | 0.07 | -0.025 | 0.289 | 0.604 | 0.38898 | non.sig |
| Bacteroides | wMCA | 1.8 | 0.07 | -0.029 | 0.331 | 0.690 | 0.38898 | non.sig |
| Bacteroides | unknown5 | 1.8 | 0.07 | -0.015 | 0.152 | 0.319 | 0.38924 | non.sig |
| Clostridiaceae_Clostridium | unknown10 | -1.8 | 0.07 | -0.801 | -0.383 | 0.036 | 0.38924 | non.sig |
| Bifidobacterium | oxo | 1.8 | 0.08 | -0.016 | 0.156 | 0.328 | 0.39555 | non.sig |
| Bifidobacterium | unknown13 | 1.8 | 0.08 | -0.016 | 0.130 | 0.277 | 0.40776 | non.sig |
| Escherichia | GCA | 1.8 | 0.08 | -0.021 | 0.180 | 0.380 | 0.40776 | non.sig |
| Bifidobacterium | x1_pentadecanol | 1.7 | 0.08 | -0.029 | 0.226 | 0.481 | 0.40909 | non.sig |
| Bifidobacterium | unknown15 | 1.7 | 0.08 | -0.042 | 0.318 | 0.679 | 0.40909 | non.sig |
| Bifidobacterium | unknown5 | 1.7 | 0.08 | -0.019 | 0.140 | 0.299 | 0.41151 | non.sig |
| Veillonella | Propionic acid | 1.7 | 0.09 | -0.008 | 0.052 | 0.113 | 0.42554 | non.sig |
| Escherichia | x3_indoleacetonitrile | -1.7 | 0.09 | -0.264 | -0.122 | 0.020 | 0.43188 | non.sig |
| Bifidobacterium | unknown17 | 1.7 | 0.09 | -0.029 | 0.172 | 0.374 | 0.43341 | non.sig |
| Bifidobacterium | n_acetyl_d_glucosamine | 1.7 | 0.10 | -0.031 | 0.176 | 0.383 | 0.43626 | non.sig |
| Bifidobacterium | unknown18 | 1.7 | 0.10 | -0.029 | 0.161 | 0.351 | 0.43809 | non.sig |
| Veillonella | unknown19 | 1.7 | 0.10 | -0.023 | 0.117 | 0.257 | 0.44001 | non.sig |
| Clostridiaceae_Clostridium | THCA | 1.7 | 0.10 | -0.056 | 0.296 | 0.649 | 0.44001 | non.sig |
| Veillonella | p_hydroxylphenyllactic_acid | 1.6 | 0.11 | -0.031 | 0.144 | 0.320 | 0.45237 | non.sig |
| Bacteroides | x3_4_dihydroxyhydrocinnamic_acid | 1.6 | 0.11 | -0.087 | 0.384 | 0.854 | 0.45237 | non.sig |
| Clostridiaceae_Clostridium | unknown26 | -1.6 | 0.11 | -0.719 | -0.323 | 0.072 | 0.45237 | non.sig |
| Escherichia | unknown20 | -1.6 | 0.11 | -0.291 | -0.131 | 0.029 | 0.45237 | non.sig |
| Bifidobacterium | 7-oxo-DCA | 1.6 | 0.11 | -0.032 | 0.147 | 0.326 | 0.45237 | non.sig |
| Bifidobacterium | secondary | 1.6 | 0.11 | -0.033 | 0.143 | 0.318 | 0.45237 | non.sig |
| Escherichia | oleic_acid_18_1n_9 | 1.6 | 0.11 | -0.034 | 0.147 | 0.328 | 0.45346 | non.sig |
| Veillonella | Valeric acid | -1.6 | 0.12 | -0.062 | -0.027 | 0.007 | 0.47567 | non.sig |
| Bifidobacterium | Valeric acid | -1.5 | 0.12 | -0.059 | -0.026 | 0.007 | 0.48862 | non.sig |
| Veillonella | lauric_acid | 1.5 | 0.13 | -0.043 | 0.150 | 0.343 | 0.49510 | non.sig |
| Bifidobacterium | TUDCA | -1.5 | 0.13 | -0.529 | -0.230 | 0.069 | 0.49709 | non.sig |
| Bacteroides | Propionic acid | 1.5 | 0.13 | -0.014 | 0.047 | 0.108 | 0.49709 | non.sig |
| Escherichia | THCA | 1.5 | 0.13 | -0.060 | 0.202 | 0.465 | 0.49709 | non.sig |
| Escherichia | Iso-butyric acid | 1.5 | 0.13 | -0.006 | 0.018 | 0.043 | 0.49709 | non.sig |
| Bacteroides | galactonic_acid | 1.5 | 0.14 | -0.070 | 0.213 | 0.495 | 0.51598 | non.sig |
| Bifidobacterium | quinolinic_acid | -1.4 | 0.15 | -0.284 | -0.120 | 0.045 | 0.55617 | non.sig |
| Escherichia | unknown15 | -1.4 | 0.15 | -0.533 | -0.225 | 0.084 | 0.55617 | non.sig |
| Escherichia | pantothenic_acid | 1.4 | 0.16 | -0.047 | 0.122 | 0.291 | 0.56140 | non.sig |
| Veillonella | x4_hydroxyphenylacetic_acid | 1.4 | 0.16 | -0.071 | 0.174 | 0.419 | 0.57749 | non.sig |
| Veillonella | stearic_acid | -1.4 | 0.16 | -0.273 | -0.113 | 0.047 | 0.57749 | non.sig |
| Bacteroides | quinolinic_acid | 1.4 | 0.17 | -0.051 | 0.120 | 0.291 | 0.57749 | non.sig |
| Bacteroides | x5_hydroxyindoleactate_1 | 1.4 | 0.17 | -0.096 | 0.226 | 0.547 | 0.57749 | non.sig |
| Escherichia | c16 | -1.4 | 0.17 | -0.477 | -0.196 | 0.085 | 0.57749 | non.sig |
| Clostridiaceae_Clostridium | TCA | 1.4 | 0.17 | -0.096 | 0.221 | 0.538 | 0.57749 | non.sig |
| Clostridiaceae_Clostridium | quinolinic_acid | 1.4 | 0.17 | -0.058 | 0.131 | 0.321 | 0.57917 | non.sig |
| Escherichia | x9_octadecen_1_ol | 1.4 | 0.18 | -0.030 | 0.067 | 0.163 | 0.58039 | non.sig |
| Clostridiaceae_Clostridium | glycoconjugated | 1.4 | 0.18 | -0.066 | 0.146 | 0.359 | 0.58039 | non.sig |
| Veillonella | unknown15 | -1.3 | 0.18 | -0.630 | -0.255 | 0.121 | 0.58137 | non.sig |
| Clostridiaceae_Clostridium | unknown15 | -1.3 | 0.18 | -0.702 | -0.285 | 0.133 | 0.58137 | non.sig |
| Escherichia | udp_glucuronic_acid | -1.3 | 0.18 | -0.235 | -0.095 | 0.045 | 0.58137 | non.sig |
| Bacteroides | Iso-butyric acid | 1.3 | 0.18 | -0.010 | 0.020 | 0.050 | 0.58137 | non.sig |
| Bifidobacterium | udp_glucuronic_acid | -1.3 | 0.19 | -0.274 | -0.110 | 0.054 | 0.58984 | non.sig |
| Escherichia | putrescine | -1.3 | 0.19 | -0.281 | -0.112 | 0.056 | 0.59354 | non.sig |
| Bifidobacterium | CA | 1.3 | 0.20 | -0.057 | 0.105 | 0.267 | 0.61996 | non.sig |
| Escherichia | CDCA | -1.3 | 0.20 | -0.283 | -0.111 | 0.061 | 0.61996 | non.sig |
| Veillonella | udp_glucuronic_acid | 1.3 | 0.21 | -0.061 | 0.110 | 0.280 | 0.62456 | non.sig |
| Bifidobacterium | 7-oxo-HDCA | 1.3 | 0.21 | -0.104 | 0.182 | 0.468 | 0.63272 | non.sig |
| Veillonella | allose1 | -1.2 | 0.22 | -0.777 | -0.297 | 0.182 | 0.63405 | non.sig |
| Veillonella | plamitic_acid_16_0 | -1.2 | 0.22 | -0.170 | -0.066 | 0.039 | 0.63405 | non.sig |
| Escherichia | x1_2_3_4_5_6_hexatrimethylsilylinositol | -1.2 | 0.22 | -0.262 | -0.101 | 0.060 | 0.63405 | non.sig |
| Veillonella | 7-oxo-DCA | -1.2 | 0.22 | -0.304 | -0.117 | 0.069 | 0.63405 | non.sig |
| Clostridiaceae_Clostridium | CDCA | 1.2 | 0.22 | -0.088 | 0.144 | 0.376 | 0.63405 | non.sig |
| Escherichia | Valeric acid | 1.2 | 0.22 | -0.011 | 0.018 | 0.046 | 0.63405 | non.sig |
| Veillonella | unknown3 | 1.2 | 0.23 | -0.053 | 0.084 | 0.220 | 0.63622 | non.sig |
| Veillonella | unknown4 | 1.2 | 0.23 | -0.101 | 0.162 | 0.425 | 0.63622 | non.sig |
| Bacteroides | GCA | -1.2 | 0.23 | -0.392 | -0.149 | 0.095 | 0.63725 | non.sig |
| Clostridiaceae_Clostridium | x9_octadecen_1_ol | 1.2 | 0.24 | -0.052 | 0.079 | 0.209 | 0.64596 | non.sig |
| Clostridiaceae_Clostridium | tauroconjugated | 1.2 | 0.24 | -0.105 | 0.160 | 0.425 | 0.64596 | non.sig |
| Bacteroides | x1_pentadecanol | -1.2 | 0.25 | -0.423 | -0.157 | 0.109 | 0.66680 | non.sig |
| Bacteroides | unknown4 | 1.1 | 0.25 | -0.110 | 0.154 | 0.418 | 0.67758 | non.sig |
| Bacteroides | oleic_acid_18_1n_9 | 1.1 | 0.26 | -0.094 | 0.128 | 0.350 | 0.68275 | non.sig |
| Escherichia | unknown21 | 1.1 | 0.26 | -0.080 | 0.108 | 0.296 | 0.68275 | non.sig |
| Escherichia | Isovaleric acid | 1.1 | 0.26 | -0.013 | 0.017 | 0.048 | 0.68275 | non.sig |
| Escherichia | TUDCA | 1.1 | 0.26 | -0.111 | 0.147 | 0.406 | 0.68445 | non.sig |
| Escherichia | tauroconjugated | 1.1 | 0.27 | -0.086 | 0.113 | 0.312 | 0.68604 | non.sig |
| Veillonella | c16 | 1.1 | 0.27 | -0.151 | 0.192 | 0.534 | 0.68633 | non.sig |
| Bacteroides | udp_glucuronic_acid | 1.1 | 0.27 | -0.075 | 0.095 | 0.266 | 0.68633 | non.sig |
| Clostridiaceae_Clostridium | unknown20 | -1.1 | 0.27 | -0.338 | -0.121 | 0.095 | 0.68633 | non.sig |
| Veillonella | bMCA | -1.1 | 0.27 | -0.418 | -0.151 | 0.117 | 0.68633 | non.sig |
| Clostridiaceae_Clostridium | l_ascorbic_acid | -1.1 | 0.28 | -0.409 | -0.146 | 0.118 | 0.68845 | non.sig |
| Clostridiaceae_Clostridium | GCA | 1.1 | 0.28 | -0.119 | 0.147 | 0.413 | 0.68845 | non.sig |
| Clostridiaceae_Clostridium | unknown3 | 1.1 | 0.28 | -0.068 | 0.083 | 0.235 | 0.68945 | non.sig |
| Escherichia | primary | -1.1 | 0.28 | -0.157 | -0.055 | 0.046 | 0.69457 | non.sig |
| Clostridiaceae_Clostridium | Valeric acid | -1.1 | 0.29 | -0.059 | -0.020 | 0.018 | 0.71347 | non.sig |
| Escherichia | x5_hydroxyindoleactate_1 | 1.0 | 0.30 | -0.125 | 0.138 | 0.401 | 0.73000 | non.sig |
| Veillonella | oxo | -1.0 | 0.31 | -0.272 | -0.093 | 0.086 | 0.73000 | non.sig |
| Escherichia | unknown18 | -1.0 | 0.31 | -0.246 | -0.084 | 0.078 | 0.73171 | non.sig |
| Veillonella | x1_2_3_4_5_6_hexatrimethylsilylinositol | 1.0 | 0.32 | -0.097 | 0.100 | 0.297 | 0.73720 | non.sig |
| Bacteroides | unknown17 | 1.0 | 0.32 | -0.104 | 0.109 | 0.322 | 0.73720 | non.sig |
| Clostridiaceae_Clostridium | wMCA | -1.0 | 0.31 | -0.602 | -0.204 | 0.195 | 0.73720 | non.sig |
| Bacteroides | d_ribose | 1.0 | 0.32 | -0.095 | 0.097 | 0.289 | 0.74225 | non.sig |
| Bifidobacterium | x3_phosphoglycerate | 1.0 | 0.33 | -0.122 | 0.122 | 0.367 | 0.74744 | non.sig |
| Veillonella | quinolinic_acid | 1.0 | 0.33 | -0.086 | 0.085 | 0.256 | 0.75244 | non.sig |
| Bifidobacterium | pentadecanoic_acid_15_0 | 1.0 | 0.33 | -0.075 | 0.073 | 0.221 | 0.75519 | non.sig |
| Escherichia | l_arginine | 1.0 | 0.34 | -0.124 | 0.117 | 0.357 | 0.75906 | non.sig |
| Escherichia | bMCA | 1.0 | 0.34 | -0.113 | 0.106 | 0.326 | 0.75906 | non.sig |
| Bifidobacterium | plamitic_acid_16_0 | 0.9 | 0.34 | -0.052 | 0.048 | 0.147 | 0.76364 | non.sig |
| Bacteroides | l_arginine | 0.9 | 0.35 | -0.153 | 0.141 | 0.434 | 0.76524 | non.sig |
| Bacteroides | Valeric acid | -0.9 | 0.35 | -0.051 | -0.016 | 0.018 | 0.76524 | non.sig |
| Bifidobacterium | p_hydroxylphenyllactic_acid | 0.9 | 0.35 | -0.087 | 0.078 | 0.243 | 0.76593 | non.sig |
| Bacteroides | x3_phosphoglycerate | 0.9 | 0.35 | -0.134 | 0.120 | 0.373 | 0.76593 | non.sig |
| Bacteroides | allose1 | 0.9 | 0.36 | -0.255 | 0.220 | 0.695 | 0.77878 | non.sig |
| Bacteroides | l_iditol | 0.9 | 0.37 | -0.257 | 0.218 | 0.694 | 0.77878 | non.sig |
| Bacteroides | stearic_acid | 0.9 | 0.36 | -0.086 | 0.074 | 0.234 | 0.77878 | non.sig |
| Clostridiaceae_Clostridium | x3_4_dihydroxyhydrocinnamic_acid | 0.9 | 0.37 | -0.287 | 0.239 | 0.764 | 0.78483 | non.sig |
| Clostridiaceae_Clostridium | GUDCA | -0.9 | 0.37 | -0.518 | -0.161 | 0.196 | 0.78483 | non.sig |
| Veillonella | x3_phosphoglycerate | 0.9 | 0.38 | -0.142 | 0.112 | 0.366 | 0.79661 | non.sig |
| Clostridiaceae_Clostridium | oleic_acid_18_1n_9 | -0.9 | 0.38 | -0.354 | -0.109 | 0.137 | 0.79661 | non.sig |
| Escherichia | unknown26 | 0.9 | 0.39 | -0.164 | 0.129 | 0.422 | 0.79859 | non.sig |
| Bifidobacterium | unknown26 | 0.9 | 0.39 | -0.194 | 0.150 | 0.494 | 0.80027 | non.sig |
| Bifidobacterium | l_iditol | 0.8 | 0.41 | -0.264 | 0.193 | 0.649 | 0.80432 | non.sig |
| Veillonella | n_acetyl_d_glucosamine | -0.8 | 0.40 | -0.305 | -0.091 | 0.124 | 0.80432 | non.sig |
| Bacteroides | x1_trimethylsilylmethyl_3_cyclohexenyl_5_methyl_4_hexen_1_ol | 0.8 | 0.40 | -0.086 | 0.064 | 0.214 | 0.80432 | non.sig |
| Veillonella | TwMCA | 0.8 | 0.40 | -0.246 | 0.187 | 0.619 | 0.80432 | non.sig |
| Clostridiaceae_Clostridium | CA | 0.8 | 0.41 | -0.107 | 0.078 | 0.263 | 0.80432 | non.sig |
| Escherichia | Butyric acid | 0.8 | 0.41 | -0.030 | 0.022 | 0.073 | 0.80432 | non.sig |
| Clostridiaceae_Clostridium | unknown13 | 0.8 | 0.41 | -0.098 | 0.071 | 0.240 | 0.80435 | non.sig |
| Bacteroides | Acetic acid | -0.8 | 0.42 | -0.074 | -0.022 | 0.031 | 0.81234 | non.sig |
| Escherichia | secondary | 0.8 | 0.42 | -0.088 | 0.062 | 0.211 | 0.81234 | non.sig |
| Escherichia | TbMCA | 0.8 | 0.42 | -0.192 | 0.133 | 0.458 | 0.81389 | non.sig |
| Veillonella | THCA | 0.8 | 0.42 | -0.191 | 0.130 | 0.451 | 0.81647 | non.sig |
| Veillonella | CDCA | 0.8 | 0.43 | -0.126 | 0.084 | 0.294 | 0.82411 | non.sig |
| Bacteroides | p_hydroxylphenyllactic_acid | 0.8 | 0.43 | -0.106 | 0.070 | 0.246 | 0.82508 | non.sig |
| Bifidobacterium | x4_hydroxyphenylacetic_acid | 0.8 | 0.45 | -0.146 | 0.091 | 0.328 | 0.83300 | non.sig |
| Bifidobacterium | c16 | 0.8 | 0.45 | -0.201 | 0.128 | 0.456 | 0.83300 | non.sig |
| Bacteroides | x9_octadecen_1_ol | 0.8 | 0.45 | -0.073 | 0.045 | 0.164 | 0.83300 | non.sig |
| Clostridiaceae_Clostridium | lauric_acid | 0.8 | 0.45 | -0.132 | 0.082 | 0.297 | 0.83300 | non.sig |
| Veillonella | secondary | -0.8 | 0.45 | -0.253 | -0.071 | 0.112 | 0.83300 | non.sig |
| Veillonella | unknown22 | -0.8 | 0.45 | -0.234 | -0.065 | 0.105 | 0.83473 | non.sig |
| Bifidobacterium | lauric_acid | -0.7 | 0.47 | -0.255 | -0.069 | 0.118 | 0.83959 | non.sig |
| Bacteroides | succinic_acid | 0.7 | 0.47 | -0.134 | 0.079 | 0.292 | 0.83959 | non.sig |
| Clostridiaceae_Clostridium | x1_pentadecanol | 0.7 | 0.48 | -0.187 | 0.107 | 0.401 | 0.83959 | non.sig |
| Clostridiaceae_Clostridium | unknown19 | 0.7 | 0.47 | -0.099 | 0.057 | 0.213 | 0.83959 | non.sig |
| Clostridiaceae_Clostridium | unknown21 | -0.7 | 0.49 | -0.345 | -0.090 | 0.164 | 0.83959 | non.sig |
| Escherichia | stearic_acid | -0.7 | 0.48 | -0.179 | -0.047 | 0.084 | 0.83959 | non.sig |
| Veillonella | TCA | 0.7 | 0.48 | -0.188 | 0.104 | 0.397 | 0.83959 | non.sig |
| Veillonella | aMCA | 0.7 | 0.48 | -0.111 | 0.063 | 0.238 | 0.83959 | non.sig |
| Veillonella | GCA | 0.7 | 0.48 | -0.157 | 0.089 | 0.335 | 0.83959 | non.sig |
| Clostridiaceae_Clostridium | TbMCA | 0.7 | 0.48 | -0.281 | 0.158 | 0.598 | 0.83959 | non.sig |
| Escherichia | wMCA | 0.7 | 0.48 | -0.188 | 0.107 | 0.402 | 0.83959 | non.sig |
| Escherichia | CA | -0.7 | 0.48 | -0.186 | -0.049 | 0.088 | 0.83959 | non.sig |
| Veillonella | unknown13 | 0.7 | 0.49 | -0.099 | 0.053 | 0.206 | 0.84054 | non.sig |
| Clostridiaceae_Clostridium | p_hydroxylphenyllactic_acid | -0.7 | 0.49 | -0.263 | -0.068 | 0.126 | 0.84054 | non.sig |
| Bifidobacterium | unknown21 | -0.7 | 0.50 | -0.296 | -0.075 | 0.146 | 0.84484 | non.sig |
| Veillonella | l_ascorbic_acid | -0.7 | 0.50 | -0.320 | -0.082 | 0.156 | 0.84484 | non.sig |
| Veillonella | l_arginine | 0.7 | 0.51 | -0.194 | 0.099 | 0.392 | 0.84484 | non.sig |
| Escherichia | unknown3 | 0.7 | 0.50 | -0.074 | 0.038 | 0.150 | 0.84484 | non.sig |
| Escherichia | x7_hexadecenyl_acetate | -0.7 | 0.50 | -0.259 | -0.067 | 0.126 | 0.84484 | non.sig |
| Bifidobacterium | oleic_acid_18_1n_9 | 0.6 | 0.52 | -0.143 | 0.070 | 0.284 | 0.84592 | non.sig |
| Bifidobacterium | stearic_acid | 0.6 | 0.53 | -0.105 | 0.049 | 0.203 | 0.84592 | non.sig |
| Veillonella | unknown20 | 0.6 | 0.54 | -0.135 | 0.061 | 0.256 | 0.84592 | non.sig |
| Veillonella | melibiose | -0.6 | 0.53 | -0.584 | -0.141 | 0.303 | 0.84592 | non.sig |
| Clostridiaceae_Clostridium | l_arginine | 0.6 | 0.54 | -0.224 | 0.100 | 0.425 | 0.84592 | non.sig |
| Escherichia | unknown4 | -0.6 | 0.54 | -0.283 | -0.068 | 0.148 | 0.84592 | non.sig |
| Escherichia | palmitoleic_acid | 0.7 | 0.51 | -0.089 | 0.044 | 0.177 | 0.84592 | non.sig |
| Veillonella | wMCA | -0.6 | 0.53 | -0.474 | -0.114 | 0.247 | 0.84592 | non.sig |
| Bacteroides | TbMCA | 0.6 | 0.54 | -0.275 | 0.123 | 0.521 | 0.84592 | non.sig |
| Bacteroides | GUDCA | 0.7 | 0.51 | -0.216 | 0.109 | 0.433 | 0.84592 | non.sig |
| Bacteroides | CDCA | 0.6 | 0.53 | -0.143 | 0.066 | 0.276 | 0.84592 | non.sig |
| Bacteroides | Isovaleric acid | 0.6 | 0.54 | -0.026 | 0.011 | 0.048 | 0.84592 | non.sig |
| Clostridiaceae_Clostridium | TwMCA | 0.6 | 0.53 | -0.324 | 0.152 | 0.629 | 0.84592 | non.sig |
| Clostridiaceae_Clostridium | Butyric acid | -0.6 | 0.54 | -0.089 | -0.021 | 0.047 | 0.84592 | non.sig |
| Escherichia | aMCA | -0.6 | 0.53 | -0.189 | -0.045 | 0.098 | 0.84592 | non.sig |
| Bacteroides | unknown21 | -0.6 | 0.56 | -0.299 | -0.069 | 0.161 | 0.85434 | non.sig |
| Bacteroides | unknown26 | -0.6 | 0.55 | -0.465 | -0.108 | 0.250 | 0.85434 | non.sig |
| Escherichia | p_hydroxylphenyllactic_acid | -0.6 | 0.55 | -0.188 | -0.043 | 0.101 | 0.85434 | non.sig |
| Bifidobacterium | succinic_acid | 0.6 | 0.57 | -0.146 | 0.059 | 0.264 | 0.85452 | non.sig |
| Bifidobacterium | putrescine | -0.6 | 0.57 | -0.255 | -0.057 | 0.141 | 0.85452 | non.sig |
| Bifidobacterium | melibiose | 0.6 | 0.57 | -0.303 | 0.125 | 0.554 | 0.85452 | non.sig |
| Bacteroides | unknown18 | 0.6 | 0.57 | -0.141 | 0.057 | 0.256 | 0.85452 | non.sig |
| Escherichia | unknown10 | 0.6 | 0.57 | -0.222 | 0.090 | 0.402 | 0.85452 | non.sig |
| Clostridiaceae_Clostridium | TUDCA | 0.6 | 0.56 | -0.244 | 0.102 | 0.448 | 0.85452 | non.sig |
| Clostridiaceae_Clostridium | palmitoleic_acid | -0.5 | 0.59 | -0.228 | -0.049 | 0.130 | 0.87295 | non.sig |
| Escherichia | pentadecanoic_acid_15_0 | -0.5 | 0.59 | -0.162 | -0.035 | 0.092 | 0.87295 | non.sig |
| Bifidobacterium | x3_4_dihydroxyhydrocinnamic_acid | 0.5 | 0.62 | -0.340 | 0.114 | 0.569 | 0.87837 | non.sig |
| Veillonella | pentadecanoic_acid_15_0 | -0.5 | 0.61 | -0.196 | -0.040 | 0.115 | 0.87837 | non.sig |
| Veillonella | x1_trimethylsilylmethyl_3_cyclohexenyl_5_methyl_4_hexen_1_ol | 0.5 | 0.60 | -0.110 | 0.040 | 0.189 | 0.87837 | non.sig |
| Bacteroides | plamitic_acid_16_0 | 0.5 | 0.62 | -0.078 | 0.027 | 0.131 | 0.87837 | non.sig |
| Bacteroides | x7_hexadecenyl_acetate | 0.5 | 0.60 | -0.172 | 0.063 | 0.299 | 0.87837 | non.sig |
| Clostridiaceae_Clostridium | unknown4 | 0.5 | 0.62 | -0.219 | 0.073 | 0.366 | 0.87837 | non.sig |
| Clostridiaceae_Clostridium | plamitic_acid_16_0 | -0.5 | 0.60 | -0.146 | -0.031 | 0.084 | 0.87837 | non.sig |
| Veillonella | HCA | -0.5 | 0.61 | -0.200 | -0.042 | 0.116 | 0.87837 | non.sig |
| Veillonella | Butyric acid | 0.5 | 0.62 | -0.047 | 0.016 | 0.079 | 0.87837 | non.sig |
| Clostridiaceae_Clostridium | 7-oxo-DCA | -0.5 | 0.61 | -0.260 | -0.054 | 0.153 | 0.87837 | non.sig |
| Clostridiaceae_Clostridium | Isovaleric acid | -0.5 | 0.61 | -0.052 | -0.011 | 0.030 | 0.87837 | non.sig |
| Clostridiaceae_Clostridium | oxo | -0.5 | 0.62 | -0.247 | -0.049 | 0.148 | 0.87837 | non.sig |
| Bacteroides | l_ascorbic_acid | -0.5 | 0.64 | -0.296 | -0.058 | 0.181 | 0.89241 | non.sig |
| Veillonella | unknown26 | 0.5 | 0.64 | -0.273 | 0.084 | 0.442 | 0.89848 | non.sig |
| Escherichia | unknown17 | -0.5 | 0.65 | -0.215 | -0.041 | 0.133 | 0.89848 | non.sig |
| Bifidobacterium | l_ascorbic_acid | 0.5 | 0.65 | -0.176 | 0.052 | 0.281 | 0.90123 | non.sig |
| Clostridiaceae_Clostridium | unknown17 | -0.5 | 0.65 | -0.287 | -0.054 | 0.180 | 0.90123 | non.sig |
| Bifidobacterium | unknown8 | 0.4 | 0.72 | -0.146 | 0.033 | 0.212 | 0.90500 | non.sig |
| Bifidobacterium | unknown19 | -0.4 | 0.72 | -0.161 | -0.025 | 0.111 | 0.90500 | non.sig |
| Veillonella | putrescine | 0.4 | 0.70 | -0.165 | 0.040 | 0.245 | 0.90500 | non.sig |
| Veillonella | unknown10 | -0.4 | 0.70 | -0.456 | -0.075 | 0.306 | 0.90500 | non.sig |
| Veillonella | l_iditol | 0.4 | 0.72 | -0.388 | 0.087 | 0.562 | 0.90500 | non.sig |
| Bacteroides | putrescine | 0.4 | 0.67 | -0.162 | 0.045 | 0.251 | 0.90500 | non.sig |
| Bacteroides | pentadecanoic_acid_15_0 | 0.4 | 0.69 | -0.123 | 0.031 | 0.185 | 0.90500 | non.sig |
| Bacteroides | unknown13 | 0.4 | 0.69 | -0.121 | 0.032 | 0.184 | 0.90500 | non.sig |
| Clostridiaceae_Clostridium | udp_glucuronic_acid | 0.4 | 0.70 | -0.153 | 0.037 | 0.226 | 0.90500 | non.sig |
| Clostridiaceae_Clostridium | allose1 | 0.4 | 0.70 | -0.426 | 0.106 | 0.638 | 0.90500 | non.sig |
| Clostridiaceae_Clostridium | x7_hexadecenyl_acetate | 0.4 | 0.68 | -0.206 | 0.055 | 0.315 | 0.90500 | non.sig |
| Clostridiaceae_Clostridium | unknown18 | 0.4 | 0.68 | -0.174 | 0.045 | 0.265 | 0.90500 | non.sig |
| Clostridiaceae_Clostridium | unknown22 | -0.4 | 0.69 | -0.227 | -0.039 | 0.150 | 0.90500 | non.sig |
| Escherichia | d_ribose | 0.4 | 0.66 | -0.123 | 0.035 | 0.194 | 0.90500 | non.sig |
| Escherichia | plamitic_acid_16_0 | -0.4 | 0.71 | -0.102 | -0.016 | 0.069 | 0.90500 | non.sig |
| Bifidobacterium | Propionic acid | -0.4 | 0.70 | -0.072 | -0.012 | 0.048 | 0.90500 | non.sig |
| Veillonella | 7-oxo-HDCA | 0.4 | 0.68 | -0.234 | 0.062 | 0.359 | 0.90500 | non.sig |
| Veillonella | Acetic acid | -0.4 | 0.71 | -0.062 | -0.010 | 0.043 | 0.90500 | non.sig |
| Veillonella | tauroconjugated | 0.4 | 0.67 | -0.190 | 0.053 | 0.296 | 0.90500 | non.sig |
| Bacteroides | HCA | 0.4 | 0.71 | -0.128 | 0.031 | 0.190 | 0.90500 | non.sig |
| Bacteroides | CA | 0.4 | 0.72 | -0.138 | 0.031 | 0.199 | 0.90500 | non.sig |
| Bacteroides | primary | 0.4 | 0.72 | -0.102 | 0.023 | 0.148 | 0.90500 | non.sig |
| Clostridiaceae_Clostridium | bMCA | 0.4 | 0.67 | -0.232 | 0.065 | 0.361 | 0.90500 | non.sig |
| Escherichia | TwMCA | -0.4 | 0.69 | -0.428 | -0.073 | 0.282 | 0.90500 | non.sig |
| Escherichia | x1_pentadecanol | 0.4 | 0.72 | -0.179 | 0.040 | 0.258 | 0.90576 | non.sig |
| Bifidobacterium | allose1 | -0.3 | 0.74 | -0.537 | -0.077 | 0.383 | 0.90660 | non.sig |
| Bifidobacterium | x1_trimethylsilylmethyl_3_cyclohexenyl_5_methyl_4_hexen_1_ol | -0.3 | 0.77 | -0.165 | -0.021 | 0.123 | 0.90660 | non.sig |
| Bifidobacterium | x5_hydroxyindoleactate_1 | 0.3 | 0.76 | -0.260 | 0.049 | 0.358 | 0.90660 | non.sig |
| Veillonella | palmitoleic_acid | 0.3 | 0.73 | -0.133 | 0.028 | 0.190 | 0.90660 | non.sig |
| Veillonella | unknown17 | 0.3 | 0.77 | -0.181 | 0.032 | 0.244 | 0.90660 | non.sig |
| Veillonella | unknown21 | -0.3 | 0.75 | -0.266 | -0.037 | 0.193 | 0.90660 | non.sig |
| Bacteroides | unknown10 | 0.3 | 0.76 | -0.320 | 0.059 | 0.437 | 0.90660 | non.sig |
| Clostridiaceae_Clostridium | c16 | -0.3 | 0.77 | -0.438 | -0.058 | 0.323 | 0.90660 | non.sig |
| Clostridiaceae_Clostridium | n_acetyl_d_glucosamine | -0.3 | 0.75 | -0.276 | -0.039 | 0.198 | 0.90660 | non.sig |
| Escherichia | x4_hydroxyphenylacetic_acid | 0.3 | 0.75 | -0.169 | 0.033 | 0.235 | 0.90660 | non.sig |
| Escherichia | x3_phosphoglycerate | -0.3 | 0.77 | -0.239 | -0.031 | 0.177 | 0.90660 | non.sig |
| Escherichia | unknown13 | -0.3 | 0.76 | -0.145 | -0.019 | 0.106 | 0.90660 | non.sig |
| Escherichia | melibiose | -0.3 | 0.73 | -0.428 | -0.063 | 0.301 | 0.90660 | non.sig |
| Bifidobacterium | CDCA | -0.3 | 0.76 | -0.233 | -0.031 | 0.171 | 0.90660 | non.sig |
| Bifidobacterium | Butyric acid | -0.3 | 0.75 | -0.070 | -0.010 | 0.051 | 0.90660 | non.sig |
| Veillonella | Isovaleric acid | -0.3 | 0.77 | -0.042 | -0.006 | 0.031 | 0.90660 | non.sig |
| Bacteroides | Butyric acid | 0.3 | 0.73 | -0.052 | 0.011 | 0.074 | 0.90660 | non.sig |
| Clostridiaceae_Clostridium | aMCA | -0.3 | 0.76 | -0.223 | -0.030 | 0.163 | 0.90660 | non.sig |
| Clostridiaceae_Clostridium | primary | 0.3 | 0.76 | -0.116 | 0.021 | 0.158 | 0.90660 | non.sig |
| Bacteroides | melibiose | -0.3 | 0.77 | -0.513 | -0.065 | 0.383 | 0.90670 | non.sig |
| Escherichia | l_iditol | 0.3 | 0.78 | -0.335 | 0.054 | 0.443 | 0.91523 | non.sig |
| Veillonella | primary | -0.3 | 0.79 | -0.141 | -0.017 | 0.107 | 0.91523 | non.sig |
| Bifidobacterium | TbMCA | 0.3 | 0.80 | -0.333 | 0.048 | 0.430 | 0.92958 | non.sig |
| Veillonella | x9_octadecen_1_ol | 0.2 | 0.81 | -0.104 | 0.015 | 0.133 | 0.93393 | non.sig |
| Escherichia | lauric_acid | 0.2 | 0.81 | -0.139 | 0.019 | 0.177 | 0.93496 | non.sig |
| Clostridiaceae_Clostridium | HCA | 0.2 | 0.82 | -0.153 | 0.021 | 0.195 | 0.93496 | non.sig |
| Clostridiaceae_Clostridium | pentadecanoic_acid_15_0 | -0.2 | 0.82 | -0.191 | -0.020 | 0.151 | 0.93701 | non.sig |
| Clostridiaceae_Clostridium | stearic_acid | -0.2 | 0.82 | -0.198 | -0.020 | 0.158 | 0.93936 | non.sig |
| Bifidobacterium | unknown4 | -0.2 | 0.86 | -0.277 | -0.023 | 0.230 | 0.94296 | non.sig |
| Bifidobacterium | l_arginine | -0.2 | 0.86 | -0.308 | -0.025 | 0.257 | 0.94296 | non.sig |
| Veillonella | x3_indoleacetonitrile | 0.2 | 0.86 | -0.158 | 0.016 | 0.189 | 0.94296 | non.sig |
| Veillonella | oleic_acid_18_1n_9 | -0.2 | 0.84 | -0.244 | -0.022 | 0.199 | 0.94296 | non.sig |
| Bacteroides | lauric_acid | -0.2 | 0.84 | -0.213 | -0.019 | 0.174 | 0.94296 | non.sig |
| Bacteroides | unknown3 | 0.2 | 0.84 | -0.124 | 0.014 | 0.151 | 0.94296 | non.sig |
| Bacteroides | unknown19 | 0.2 | 0.85 | -0.128 | 0.013 | 0.154 | 0.94296 | non.sig |
| Escherichia | l_ascorbic_acid | 0.2 | 0.86 | -0.177 | 0.018 | 0.213 | 0.94296 | non.sig |
| Escherichia | galactonic_acid | 0.2 | 0.85 | -0.210 | 0.022 | 0.254 | 0.94296 | non.sig |
| Escherichia | n_acetyl_d_glucosamine | 0.2 | 0.86 | -0.161 | 0.016 | 0.192 | 0.94296 | non.sig |
| Veillonella | TbMCA | 0.2 | 0.84 | -0.357 | 0.040 | 0.437 | 0.94296 | non.sig |
| Bacteroides | TUDCA | 0.2 | 0.86 | -0.287 | 0.028 | 0.344 | 0.94296 | non.sig |
| Escherichia | Propionic acid | 0.2 | 0.86 | -0.046 | 0.004 | 0.055 | 0.94296 | non.sig |
| Bacteroides | unknown22 | -0.2 | 0.88 | -0.184 | -0.013 | 0.157 | 0.95267 | non.sig |
| Escherichia | unknown19 | -0.2 | 0.88 | -0.124 | -0.009 | 0.106 | 0.95267 | non.sig |
| Bifidobacterium | Iso-butyric acid | 0.1 | 0.88 | -0.026 | 0.002 | 0.031 | 0.95267 | non.sig |
| Clostridiaceae_Clostridium | Acetic acid | -0.1 | 0.88 | -0.063 | -0.004 | 0.054 | 0.95267 | non.sig |
| Veillonella | x1_pentadecanol | -0.1 | 0.89 | -0.284 | -0.018 | 0.248 | 0.96138 | non.sig |
| Veillonella | d_ribose | 0.1 | 0.90 | -0.181 | 0.012 | 0.205 | 0.96258 | non.sig |
| Clostridiaceae_Clostridium | pantothenic_acid | -0.1 | 0.90 | -0.244 | -0.015 | 0.215 | 0.96258 | non.sig |
| Escherichia | quinolinic_acid | -0.1 | 0.90 | -0.148 | -0.009 | 0.131 | 0.96258 | non.sig |
| Bifidobacterium | unknown22 | 0.1 | 0.91 | -0.154 | 0.009 | 0.173 | 0.96310 | non.sig |
| Escherichia | allose1 | -0.1 | 0.92 | -0.414 | -0.021 | 0.373 | 0.96310 | non.sig |
| Escherichia | x1_trimethylsilylmethyl_3_cyclohexenyl_5_methyl_4_hexen_1_ol | 0.1 | 0.91 | -0.116 | 0.007 | 0.130 | 0.96310 | non.sig |
| Veillonella | Iso-butyric acid | -0.1 | 0.92 | -0.031 | -0.002 | 0.028 | 0.96310 | non.sig |
| Escherichia | Acetic acid | 0.1 | 0.91 | -0.041 | 0.002 | 0.045 | 0.96310 | non.sig |
| Clostridiaceae_Clostridium | d_ribose | 0.1 | 0.92 | -0.204 | 0.010 | 0.224 | 0.96471 | non.sig |
| Clostridiaceae_Clostridium | melibiose | -0.1 | 0.94 | -0.511 | -0.018 | 0.475 | 0.96471 | non.sig |
| Escherichia | x3_4_dihydroxyhydrocinnamic_acid | 0.1 | 0.93 | -0.370 | 0.018 | 0.407 | 0.96471 | non.sig |
| Bifidobacterium | Isovaleric acid | 0.1 | 0.93 | -0.034 | 0.002 | 0.037 | 0.96471 | non.sig |
| Veillonella | GUDCA | 0.1 | 0.94 | -0.310 | 0.013 | 0.337 | 0.96471 | non.sig |
| Clostridiaceae_Clostridium | 7-oxo-HDCA | 0.1 | 0.93 | -0.314 | 0.014 | 0.342 | 0.96471 | non.sig |
| Clostridiaceae_Clostridium | Iso-butyric acid | 0.1 | 0.94 | -0.031 | 0.001 | 0.034 | 0.96471 | non.sig |
| Escherichia | HCA | 0.1 | 0.94 | -0.125 | 0.005 | 0.134 | 0.96471 | non.sig |
| Veillonella | pantothenic_acid | -0.1 | 0.95 | -0.214 | -0.007 | 0.200 | 0.96785 | non.sig |
| Escherichia | succinic_acid | -0.1 | 0.95 | -0.179 | -0.005 | 0.169 | 0.97022 | non.sig |
| Escherichia | unknown22 | 0.1 | 0.96 | -0.136 | 0.004 | 0.143 | 0.97270 | non.sig |
| Clostridiaceae_Clostridium | x3_phosphoglycerate | 0.0 | 0.96 | -0.288 | -0.007 | 0.274 | 0.97429 | non.sig |
| Clostridiaceae_Clostridium | l_iditol | 0.0 | 0.98 | -0.520 | 0.006 | 0.533 | 0.98828 | non.sig |
| Veillonella | CA | 0.0 | 0.98 | -0.169 | -0.002 | 0.165 | 0.98828 | non.sig |
| Clostridiaceae_Clostridium | galactonic_acid | 0.0 | 0.99 | -0.312 | 0.002 | 0.317 | 0.99250 | non.sig |
| Veillonella | succinic_acid | 0.0 | 0.99 | -0.212 | -0.001 | 0.210 | 0.99483 | non.sig |
